# MambaSleepCVD for prediction of long term cardiovascular outcomes from polysomnography

**DOI:** 10.64898/2026.09.08.26361626

**Authors:** Alessia Calzoni, Alvise Dei Rossi, Giuliana Monachino, Beatrice Zanchi, Luigi Fiorillo, Mattia Savardi, Francesca Dalia Faraci, Alberto Signoroni

## Abstract

Long-term cardiovascular disease (CVD) risk prediction remains a major global clinical priority. This study investigates how much prognostic information full-night polysomnography (PSG) carries after explicitly controlling demographic confounding, and whether combining specialized unimodal representations with continuous fusion mechanisms can effectively leverage it. We adopt a modular framework based on Coupled Mamba for simultaneous cross-modal and temporal fusion, capturing long-range dependencies and dynamic interactions between physiological modalities throughout sleep. With this capability we assess whether specialized supervised representations from task-specific unimodal encoders offer prognostic value comparable to task-agnostic multimodal self-supervised pre-training. Validated on the Sleep Heart Health Study (SHHS) dataset, our approach achieves performance comparable to or better than current state-of-the-art architectures in the fully supervised, data-abundant regime, although the cardiac channel alone accounts for most of the discriminative signal. More importantly, we identify a significant age-related bias in the reference dataset that, if left unaddressed, creates a shortcut leading to inflated risk predictions, accounting for a large part of the literature’s reported performance. Ultimately, our findings suggest that supervised unimodal experts integrated through a fusion engine provide a flexible pathway for prognostic risk ranking from sleep recordings, while reliable CVD screening will require independent external cohorts and explicit confounder control.

## Introduction

The relationship between sleep and cardiovascular disease (CVD) risk has been increasingly recognized. Several clinical and epidemiological studies have identified multiple physiological pathways through which sleep affects cardiovascular health [1–4]. Recognizing that sleep influences all other components of cardiovascular well-being, the American Heart Association has recently integrated optimal sleep duration into its “Life’s Essential 8” framework as a primary preventive health goal [2, 5]. Sleep health encompasses several elements, with sleep duration being central, as well as quality, efficiency, and regularity [1, 6]. Polysomnography (PSG), the gold standard for sleep evaluation, represents the most comprehensive approach for capturing the majority of these complex sleep dimensions within a single assessment. It records synchronous physiological signals throughout the whole night, such as electroencephalographic activity (electroencephalogram, EEG), eye movements (electrooculogram, EOG) and heart activity (electrocardiogram, ECG). Traditional quantification of cardiovascular risk through PSG typically relies on aggregate summary metrics, such as total sleep time, sleep latency, slow wave sleep, etc. [2, 7, 8]. However, these static indices often simplify the complexity of sleep physiology embedded within raw PSG signals. To overcome these limitations, deep learning architectures can be used to process high-dimensional raw data directly, yielding more holistic representations [9].

Despite this potential, the direct processing of full-night PSG recordings, typically spanning eight hours of multi-channel data, exposes a fundamental architectural challenge. Latest existing approaches, that aim to extract information from full-night PSG data, are based on a large-scale self-supervised learning (SSL) pre-training that tries to align different physiological modalities into a shared representation space. Due to the quadratic complexity of self-attention, current Transformer-based backbones are computationally unable to process such extensive sequences as a single, continuous input. Consequently, existing approaches often rely on frameworks that perform multi-modal fusion only within short segments or patches of the recording, while the long-range full-night temporal relationships are subsequently aggregated using recurrent or temporal modules in the finetuning phase to capture the overnight progression. During pre-training, such models aim to align information from different modalities within short-term segments: this fragmented processing inherently limits the model’s ability to capture the continuous, fine-grained cross-modal synergies that evolve across the entire sleep cycle [10–13]. To capture these potential long-range cross-modal interactions, we adopt a different paradigm involving an architecture capable of simultaneously processing the entire eight-hour sequence while performing continuous cross-modal fusion. In this direction, State Space Models (SSMs), and in particular the Mamba architecture [14], have gained significant attention as a promising alternative for sequence modeling. Mamba offers linear-time complexity while maintaining the ability to capture long-range dependencies, effectively matching the performance of state-of-the-art Transformers without their computational constraints [14, 15]. This architecture has recently demonstrated high efficacy across several biomedical domains by efficiently capturing fine-grained relationships between heterogeneous units of information (e.g., imaging, genomics, clinical notes) [15]. In the context of PSG analysis, to address its intrinsic multimodal complexity, Coupled Mamba [16] emerges as a fitting solution. It is an extension of Mamba specifically designed to facilitate the dynamic fusion of distinct signals within a unified multimodal framework. Unlike hierarchical approaches that first align modalities within short patches and subsequently aggregate them over time, Coupled Mamba enables a simultaneous temporal and cross-modal fusion. This theoretical concept better serves and investigates complex physiological systems evolution in time, like the interplay of cardiovascular system and sleep dynamics.

The central question of this work is how much prognostic information full-night PSG actually carries for long-term CVD risk once demographic confounding is explicitly controlled. To address it, we investigate the prognostic value of specialized unimodal representations, coupled with modular fusion architectures, as an alternative to the multimodal SSL pre-training paradigm currently dominating state-of-the-art CVD risk prediction from PSG. The strength of the multimodal SSL approach lies in its ability to theoretically extract complementary information across modalities during the pre-training phase, while avoiding task-specific labels from clinicians. Recent foundation models achieve this cross-modal alignment with unlabeled multivariate PSG and differ mainly in the pre-training objective: masked modeling, as in PFTSleep [11], He et al. [13], contrastive learning, as in SleepFM [10], or multi-pretext strategies combining time-frequency contrastive learning with masked autoencoding, as in SleepGPT [12]. Although these approaches differ in their specific SSL objectives and modality configurations, they generally establish cross-modal relationships during pretraining. These relationships are typically learned within restricted temporal windows, so modality-specific representations cannot interact dynamically while their trajectories are modeled across the complete recording. Although the current trend for downstream prognostic tasks heavily favors these SSL paradigms, it remains debatable whether SSL-derived embeddings offer superior prognostic value compared to specialized representations extracted from traditional supervised tasks, particularly when the downstream task is known to be associated with the supervised pre-training task, as is the case of CVD and sleep architecture.

High-quality representations extracted from unimodal models could already encapsulate the critical physiological information necessary for long-term CVD risk prediction. Traditionally, much of the deep-learning literature in PSG analysis has centered on sleep staging, the automatic determination of Wake, N1, N2, N3, and REM stages. Because sleep staging depends on both local waveform characteristics and temporal context, it provides a rich supervisory signal for learning physiological representations that generalize beyond sleep-stage classification. Supervised pre-training leverages the large availability of expert-annotated sleep-stage labels to learn specialized physiological representations [17]. Although human annotations are inherently noisy due to inter-rater variability [18], large-scale, heterogeneous datasets from repositories like the National Sleep Research Resource (NSRR) [19] still provide the diversity needed to train highly robust models. Architectures such as U-Sleep [20], and the supervised encoders benchmarked in SLEEPYLAND [21], typically operate on single channels of a single modality and combine additional channels only by voting or ad-hoc fusion [22], reaching expert-level sleep-staging performance, including on unseen external cohorts. Since sleep architecture is intrinsically linked to cardiovascular vulnerability [1–3, 23], embeddings from these task-specific encoders could provide highly competitive prognostic value compared to multimodal foundation models, which currently lag behind supervised alternatives for external cohorts [10]. Complementing this neurophysiological perspective, general-purpose ECG foundation models, such as the self-supervised contrastive framework Self-DANA [24], act as specialized cardiac experts capable of extracting robust features from the single-lead ECG setups typical of PSG. Therefore, provided they are integrated through a sufficiently powerful fusion engine, these unimodal representations could already provide information needed for long-term CVD risk prediction, without complex multimodal pre-training.

Rather than viewing integrated multimodal pre-training as a mandatory prerequisite, we shift the focus toward developing flexible fusion mechanisms capable of synthesizing informative unimodal representations across the entire eight-hour recording. This modular perspective offers a significant advantage in terms of flexibility: while multimodal foundation models are often rigidly constrained by the specific modalities included during their pre-training phase, a modular framework allows for the seamless integration or swapping of modality-specific experts, together with directly using encoders trained end-to-end on raw PSG waveforms. For this aim, we introduce MambaSleepCVD, a modular framework that enables the simultaneous and continuous fusion of full-night PSG signals. This approach allows the model to maintain a global context of the entire recording without the unsustainable memory requirements of full-sequence attention. Coupled Mamba is adopted here as an existing long-sequence fusion mechanism to enable continuous full-night multimodal interaction, and we use this modular framework for confounder-controlled evaluation of overnight PSG physiology in long-term CVD risk prediction. We validate our framework on the Sleep Heart Health Study (SHHS) dataset [25, 26], a large-scale, multi-center prospective cohort that serves as the gold standard for sleep-related cardiovascular research. SHHS2, the follow-up visit of the same cohort, is used for within-cohort temporal evaluation, as it comprises recordings of SHHS1 participants obtained several years later, thereby enabling assessment of model performance under temporal distribution shift. We model risks through a multi-label survival analysis task, aiming to predict the time-to-event of different CVD outcomes. Beyond performance evaluation, we place at the centre of this work the necessity of a rigorous examination of the dataset’s characteristics. Large-scale clinical cohorts often hold inherent biases and population-specific distributions that, if not properly accounted for, can lead to misleading interpretations and artificially inflated results, lacking clinical validity. In particular, we address the risk of “shortcut learning” [27], a phenomenon where models achieve high predictive performance by exploiting strong statistical correlations rather than capturing the underlying physiological mechanisms of disease. In a context where simple variables can act as strong proxies and become the primary drivers of performance, seemingly straightforward design choices can result in misleading conclusions. Underestimating architectural decisions may inadvertently obscure the evaluation and hinder the extraction of meaningful information from physiological signals. Therefore, we perform a controlled analysis of demographics to decouple the contributions of the learned sleep representations from simple demographic proxies, ensuring that our findings reflect cardiovascular-sleep synergies rather than chronological age.

In summary, our contributions are four-fold:

- we identify and quantify a critical demographic bias in the reference sleep dataset for CVD risk prediction, showing that chronological age acts as a dominant statistical shortcut that inflates survival metrics and must be controlled before any claim about the predictive value of physiological signals can be made;
- under this confounder-controlled setting, we quantify how much prognostic information full-night PSG actually carries, showing that the cardiac channel accounts for most of the discriminative signal while the neurophysiological channels contribute mainly to stability across recording periods;
- we show that supervised representations provide a robust alternative to complex self-supervised pre-training, challenging the assumption that task-agnostic PSG pre-training necessarily yields more generalizable representations for CVD risk prediction;
- we demonstrate the efficacy of Coupled Mamba for full-night multimodal fusion, providing a scalable and modular testbed and a practical pathway for future work on deep-learning-based CVD risk stratification in sleep medicine.

## Results

We describe cohort construction, preprocessing, architecture, and training procedures in detail in Methods. Briefly, we used full-night PSG from the SHHS and adopted SleepFM as the primary SSL benchmark. This choice is motivated by the model’s significant impact on the field, the comprehensiveness of the SHHS dataset employed in its validation, and the public availability of its pre-trained weights and architectures, thereby enabling results replication as well as facilitating comparative evaluations across different dataset settings [10]. On SHHS1 (*n* = 5774), we therefore used the test partition proposed by SleepFM as our internal test set (*n* = 1991) and split the remaining participants into training and validation. SHHS2 (*n* = 2618), a follow-up visit of the same parent cohort several years later, was reserved for within-cohort temporal evaluation. Because some SHHS2 participants also appear in the SHHS1 training set, SHHS2 is reported both as a complete cohort and after splitting into SHHS1-training-overlap and SHHS1-training-disjoint subsets. The montage used comprises EEG, EOG, and ECG. Long-term CVD risk was formulated as a multi-label survival task over six endpoints: Angina, CVD Death, Congestive Heart Failure (CHF), Coronary Heart Disease (CHD) Death, Myocardial Infarction (MI), and Stroke. For each endpoint the model assigns a prognostic risk score used to rank participants by time-to-event. Event times with undocumented onset, concentrated in Angina, were masked for the affected labels rather than imputed (Figure 1). Event counts for the datasets splits are summarized in Table 1.

**Fig. 1:**
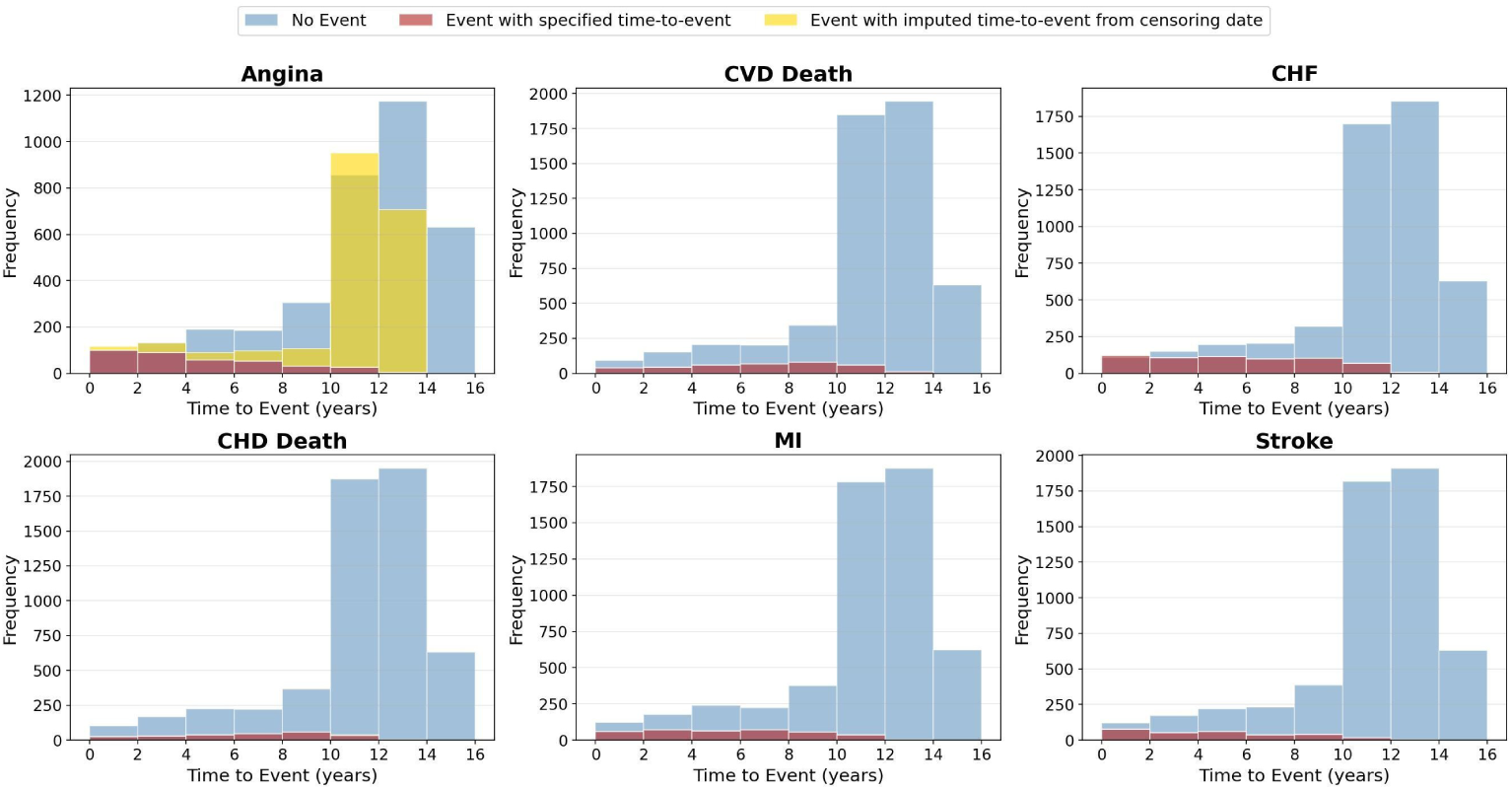
Frequency distribution of time-to-event (in years) across CVD labels. Blue indicates subjects with no events; red and yellow together mark recorded events. Within the event group, confirmed onset dates (red) are distinguished from missing onset times shown at the censoring date for illustration only (yellow). In all analyses, labels with undocumented onset were masked rather than imputed.

**Table 1:**
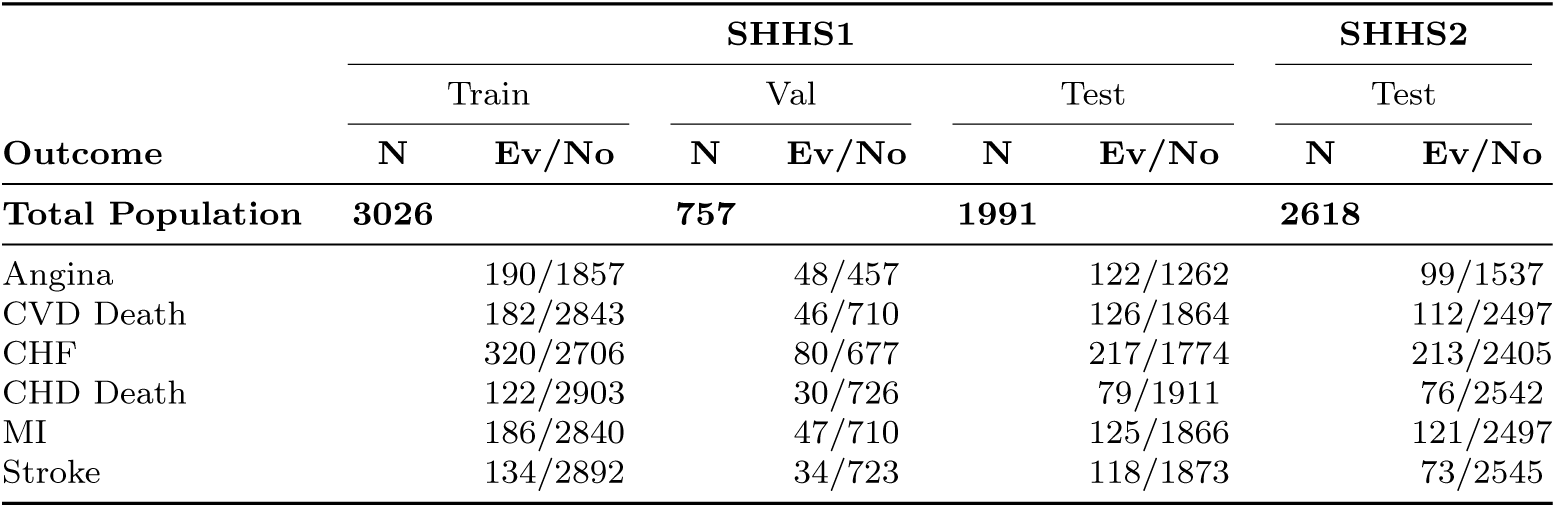
Distribution of CVD outcomes in SHHS1 test and SHHS2. For each split the total number of subjects (N) is reported, together with the number of events (Ev) and the number of non-events (No). These counts account for the masking protocol applied to subjects with “uncertain” clinical status.

MambaSleepCVD comprises modality-specific encoders, a Coupled Mamba fusion stage that integrates the synchronized full-night sequences, and a multi-label survival head (Figure 2). Encoder embeddings were taken from lightweight convolutional networks trained end-to-end, from frozen U-Sleep models pretrained for sleep staging, or from a hybrid setting in which Self-DANA replaces the ECG branch. These configurations were compared with a reimplementation of SleepFM that uses its original LSTM temporal head (hereafter SleepFM + LSTM) [10]. Holding encoder embeddings fixed, Coupled Mamba was also substituted with attention-based LSTM fusion and with late unimodal state-space fusion. Unless otherwise specified, models received only physiological waveforms; age, sex, BMI, and medications were added only in dedicated sensitivity experiments. Discrimination was quantified primarily by Harrell’s C-index, with 95% confidence intervals from 100 bootstrap iterations.

**Fig. 2:**
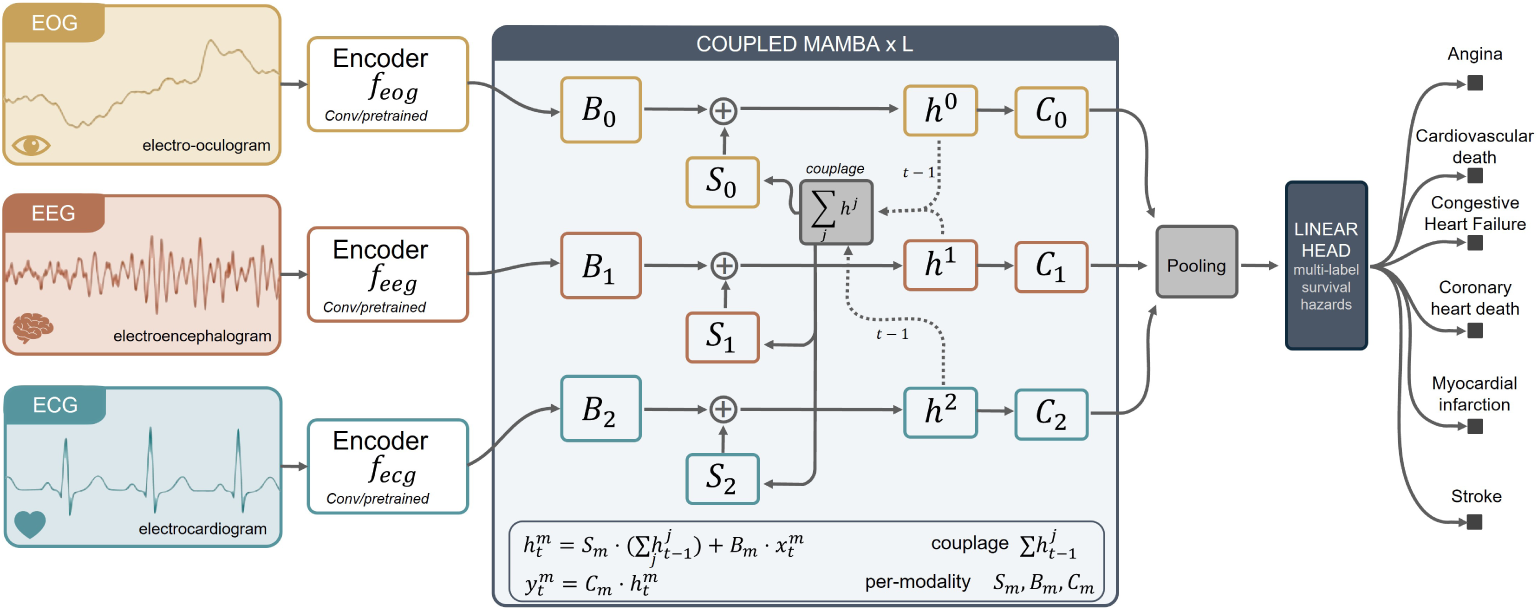
Schematic representation of the proposed modular framework. The architecture consists of: the modality-specific encoders, utilizing either end-to-end convolutional layers or frozen pre-trained unimodal models to extract synchronized feature tokens; the Coupled Mamba layer, which processes these features through a stack of *L* blocks with shared state-space recurrence, enabling continuous cross-modal interaction in a latent space; and the survival prediction head, where temporal sequences are aggregated to estimate the hazards for six cardiovascular outcomes.

### Impact of Age

Our preliminary analysis of the SHHS1 cohort revealed a significant age-related distribution shift. As shown in Supplementary Figure S1, all CVD outcomes exhibit a clear bias towards older age groups. While the visual distribution highlights the clinical trend, Table 2 provides quantitative confirmation, showing that these differences are statistically significant with moderate-to-strong effect sizes. Corresponding analyses for the SHHS2 temporal evaluation cohort and for the SHHS1 training, validation, and test partitions are reported in Supplementary Tables S1, S2, S3, S4, respectively. A deeper investigation into the cohort’s temporal structure reveals an extensive confounding mechanism between age and survival outcomes. As illustrated in Figure 3, we observe a systematic inverse correlation between age and time-to-event across all considered CVD labels. This coupling has profound mathematical implications for model optimization and evaluation. Both the CoxPH loss and Harrell’s C-index are fundamentally driven by the relative ordering of survival times. They both evaluate the model based on its ability to distinguish an individual experiencing an event from a set of subjects who survive for a longer duration (see Equations 3, 5). In a dataset where age and survival time are inversely correlated, these mathematical comparisons systematically contrast older individuals (who experience events earlier) with younger counterparts (who remain at risk). As a result, the optimization objective is heavily biased toward biological aging, as correctly ranking these “older/shorter survival” subjects against “younger/longer survival” subjects becomes the most efficient path to minimizing the loss and maximizing the concordance score.

**Fig. 3:**
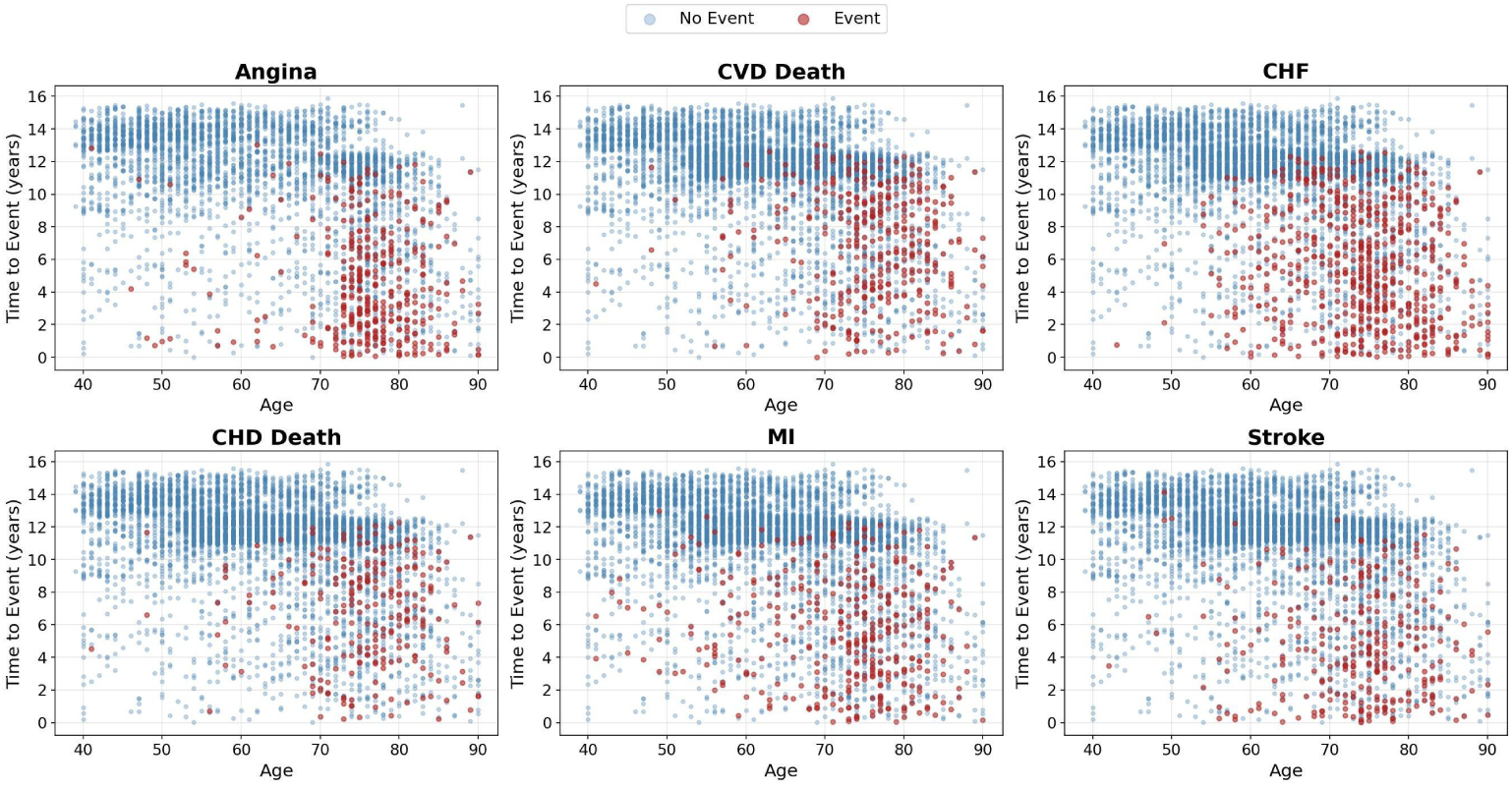
Distribution of baseline age versus time-to-event across CVD outcomes in SHHS1. Scatter plots illustrate the relationship between participant age at baseline and survival time (either event date or censoring date) for the six CVD outcomes. Red points represent subjects who experienced an event, while blue points indicate censored observations (no event).

**Table 2:**
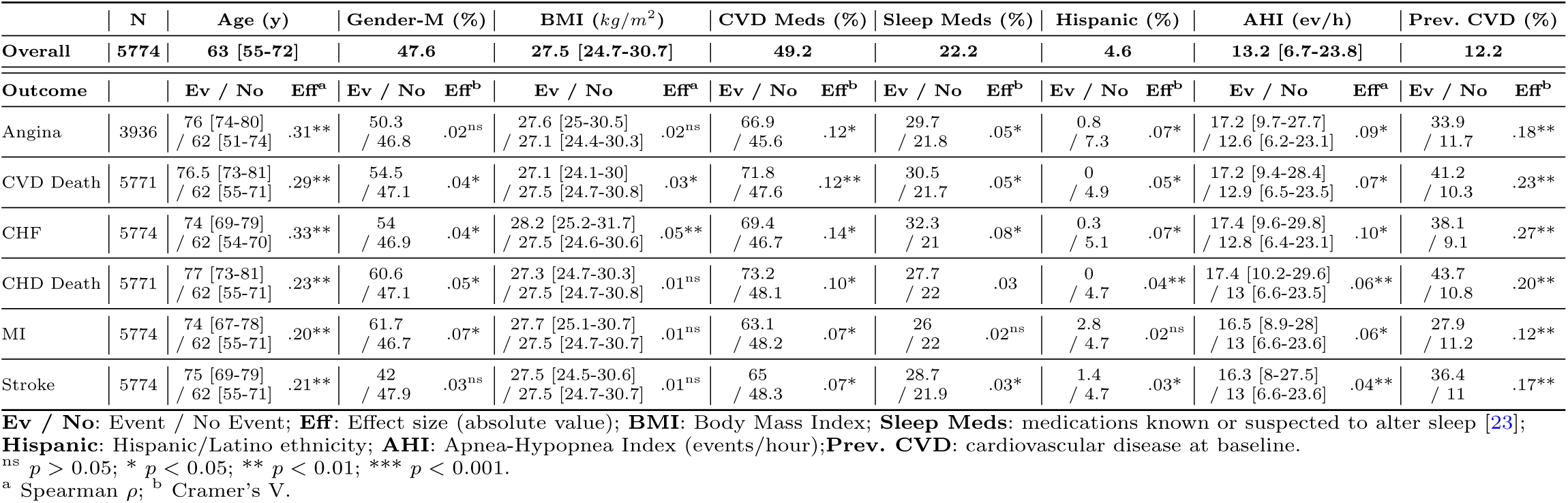
Baseline characteristics of the SHHS1 study population. Continuous variables are presented as median [interquartile range], while categorical variables are reported as percentages. Statistical significance was assessed using the Mann-Whitney test for continuous variables and the *χ*^2^-test for categorical data. The strength of associations (effect size) was quantified using Spearman’s rank correlation for continuous variables and Cramer’s V for categorical variables.

To further investigate how integrating demographics near the decision layer may encourage the gradient to flow through the age shortcut, we conducted an analysis using SleepFM [10] as a reference framework. We compared a re-implementation of the full multimodal SleepFM architecture against a demographic-only Multi-Layer Perceptron (MLP) baseline, trained exclusively on age and gender. These experiments demonstrate that SleepFM’s global performance metrics can be largely replicated using only two demographic variables, effectively bypassing the need for complex physiological feature extraction (see Supplementary Table S5). Furthermore, the direct impact of the age-survival inverse correlation on the C-index is clearly illustrated by the significant performance drop observed when evaluating the models within narrower, age-homogeneous subgroups (Supplementary Figure S2). By separating these factors, we provide a more direct assessment of the model’s capacity to identify independent CVD signatures from physiological signals.

### Age Estimation

Because excluding age as an explicit input does not preclude age-related information from being present in overnight physiology, we quantified how well chronological age could be recovered from no-demographics embeddings (Supplementary Figures S3 and S4). On SHHS1, age was moderately recoverable from both U-Sleep and SleepFM representations (*R*^2^=0.52 and 0.53, respectively). On SHHS2, recoverability remained moderate for SleepFM and lower for U-Sleep. These results indicate that age-related information is partially embedded in pretrained overnight representations and may contribute to no-demographics prognostic performance, while a substantial fraction of chronological age remains unexplained. Accordingly, the no-demographics setting should be interpreted as reducing explicit demographic confounding at the decision layer, not as establishing fully age-independent prognostic value.

### Incremental Prognostic Value of PSG-derived Risk Scores

To determine whether the learned representations provide clinical value beyond established risk factors, we evaluated PSG-derived risk scores from our no-demographics, all-modalities configurations (U-Sleep, Self-DANA, and Raw + Coupled Mamba) in multivariable Cox proportional hazards models adjusted for age, sex, BMI, baseline cardiovascular history, cardiovascular medication use, systolic blood pressure, hypertension, and diabetes. After adjustment, higher PSG risk scores remained associated with increased hazard of CVD Death, CHD death, and CHF across configurations (HR per 1 SD approximately 1.4–1.6 for fatal outcomes and 1.25–1.43 for CHF; Supplementary Table S6), whereas associations for Angina, MI, and Stroke were weaker or not significant. This result supports the endpoint-dependent incremental prognostic value of overnight PSG information beyond the included clinical risk factors.

### Sensitivity to Endpoint Formulation

We compared the primary joint multi-label formulation with independent single-label survival models trained under matched conditions. Macro-average discrimination was comparable between the two formulations on both SHHS1 and SHHS2 (Supplementary Tables S7 and S8). Because Angina remains the most uncertain endpoint, we additionally performed a specific analysis excluding Angina (Supplementary Tables S9 and S10). Macro-average differences relative to the six-endpoint model were negligible. Altogether, these sensitivity analyses support retaining the SleepFM-aligned multi-label, six-endpoint formulation as the primary experimental setting.

### Comparison with State-of-the-Art

We benchmark the performance of our proposed frameworks against the state-of-the-art SleepFM + LSTM architecture. Crucially, in every experimental configuration, all models utilize the full set of physiological signals (ECG, EEG, and EOG). To evaluate the robustness and generalizability of the models, we report results for both the SHHS1 internal test set and the SHHS2 within-cohort temporal validation set, with 95% confidence intervals derived from 100 bootstrap iterations. Furthermore, to assess whether these findings were affected by participant overlap, SHHS2 was stratified into SHHS1-training-overlap and SHHS1-training-disjoint subsets. The age-shortcut pattern observed across demographic configurations was preserved in both subsets (Figure 4 and Supplementary Figure S5), indicating that it was not confined to participants whose baseline recordings had been used for model training. For our proposed models, we explored a comprehensive demographic integration (“All Demo”) and an ablation group specifically excluding chronological age (“Without Age”). To facilitate a direct comparison with the methodology established by SleepFM [10], we evaluated both our architectures and the SleepFM + LSTM baseline on configurations combining the physiological signals with “Only Age + Gender” and “Only Gender”. Notably, considering a comprehensive set of demographics (i.e., age, gender, BMI, and both cardiovascular and sleep-related medications), alongside the physiological data, yields performance levels comparable to those achieved when aggregating only age and gender information (the choice followed by SleepFM [10]). However, the isolated removal of chronological age from these configurations (“Without Age” and “Only Gender” groups) leads to a significant and sharp drop in results, confirming that age is the primary predictive feature in this dataset.

**Fig. 4:**
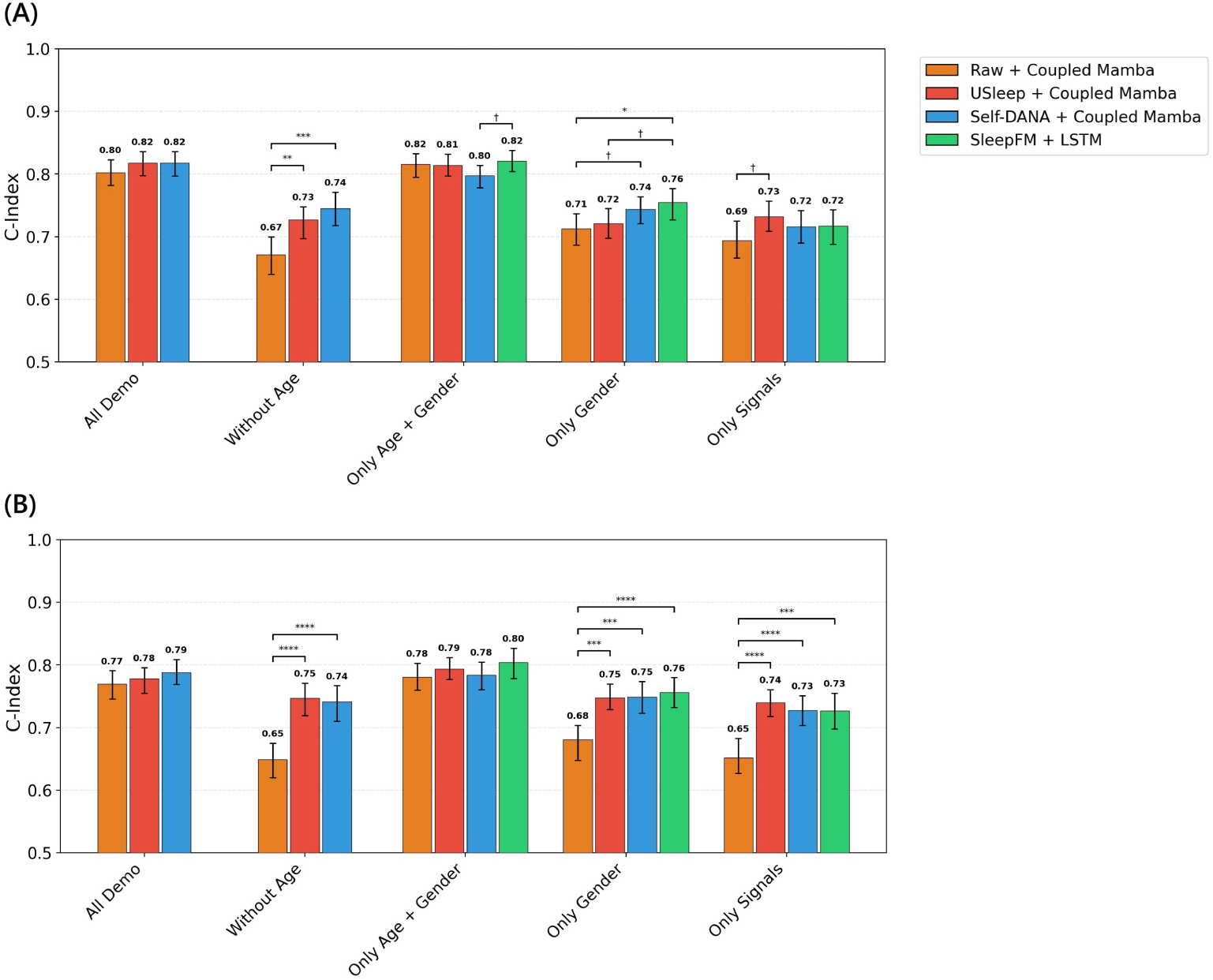
Macro-average C-index performance across demographic configurations. Comparative analysis of the proposed Coupled Mamba framework (utilizing Raw, U-Sleep, and Self-DANA encoders) against the SleepFM + LSTM baseline. In each configuration, the models utilize the full set of physiological signals (ECG, EEG, and EOG) combined with varying subsets of demographics. Results are reported for both **(A)** the SHHS1 internal test set and **(B)** the SHHS2 within-cohort temporal validation set. The “age shortcut effect” is demonstrated by the significant performance drop when age is isolated and removed from the demographic inputs. Error bars represent 95% confidence intervals derived from 100 bootstrap iterations. Statistical significance between model configurations: † p *≤* 0.1; * p *≤* 0.05; ** p *≤* 0.01; *** p *≤* 0.001; **** p *≤* 0.0001

Focusing on the no-demographics scenario, our approach, based on U-Sleep representations fused through Coupled Mamba, slightly but consistently outperforms the SleepFM + LSTM baseline. Furthermore, the substantial performance gap between the pre-trained encoders and the Raw + Coupled Mamba model underscores the necessity of pre-training. This suggests that the raw signals alone, without a structured representational foundation, are insufficient for the model to extract deep prognostic insights from the overnight waveforms.

Finally, while we report macro-average metrics here for brevity, a detailed breakdown of performance for each of the six specific CVD labels is provided in Supplementary Tables S11 and S12. Five-year macro-average AUROC analyses yielded discrimination patterns concordant with the C-index across the same experimental configurations and are reported, for each endpoint, in Supplementary Tables S13 and S14; absolute-risk calibration at the same horizon is reported in Supplementary Table S15.

### Downstream Sample-Efficiency Analysis

Under the reduced-training protocol, U-Sleep + Coupled Mamba remained competitive or superior to SleepFM + LSTM across all SHHS1 training fractions (Supplementary Figures S6 and S7). At 10%, U-Sleep showed higher point estimates and lower variability across training subsets, with overlapping confidence intervals. At 20%, the two approaches were broadly comparable, and at 50% U-Sleep remained competitive. Endpoint-specific curves followed the same pattern, with the clearest supervised advantage in the lowest-data regime. In this specific scenario, these findings indicate greater stability of the U-Sleep pipeline compared to SleepFM.

### Modality Ablation Study

To evaluate the contribution of each physiological stream, we performed a systematic ablation study on input modalities. All experiments in this section were conducted in the no-demographics configuration to ensure that the observed performance is driven by the physiological waveforms. The results for SHHS1 test set and the complete SHHS2 are illustrated in Figure 5, while Supplementary Figure S8 details the SHHS2 evaluation stratified by SHHS1 training-set overlap.

**Fig. 5:**
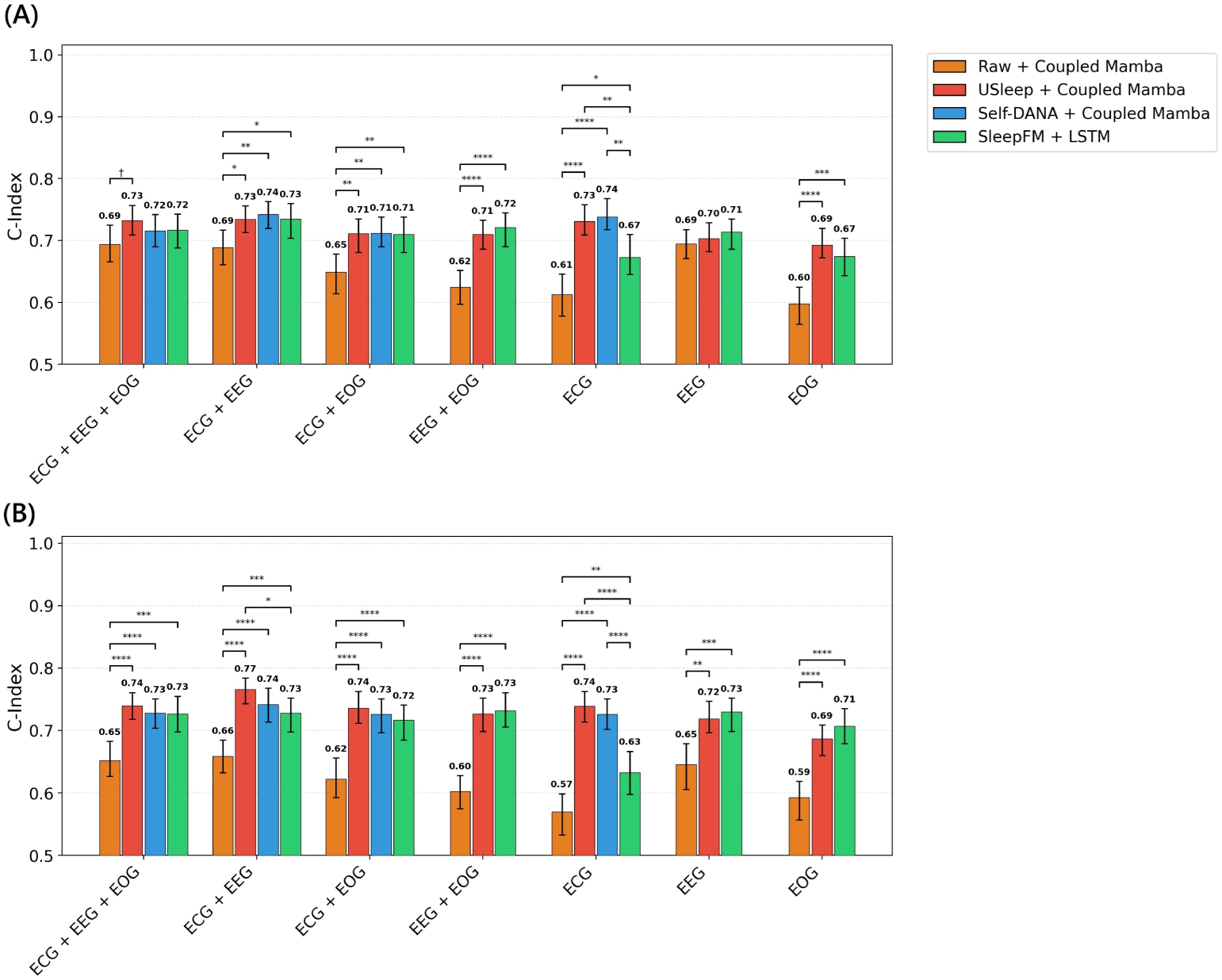
Macro-average C-index performance across modality ablation configurations. Comparative analysis of the proposed framework and the SleepFM + LSTM baseline using different combinations of physiological signals (ECG, EEG, and EOG). Notably, the Self-DANA + Coupled Mamba configuration is omitted in neurophysiological-only experiments (EEG, EOG, and EEG+EOG) as the Self-DANA encoder is specifically designed for specialized cardiac feature extraction. To ensure results are driven by overnight waveforms, all experiments are conducted in the no-demographics configuration. Results are reported for **(A)** the SHHS1 internal test set and **(B)** the SHHS2 within-cohort temporal validation set. Error bars represent 95% confidence intervals derived from 100 bootstrap iterations. Statistical significance between model configurations: † p *≤* 0.1; * p *≤* 0.05; ** p *≤* 0.01; *** p *≤* 0.001; **** p *≤* 0.0001

In the internal SHHS1 test set, our U-Sleep + Coupled Mamba framework demonstrates performance parity with the SleepFM + LSTM baseline (C-index 0.73 vs 0.72, respectively). In this cohort, the predictive power is largely driven by the cardiac signal. Specifically, when examining the ECG-only configuration, specialized representations from both Self-DANA and U-Sleep exhibit comparable prognostic value, matching or even exceeding the performance of the full multimodal models. In contrast, SleepFM exhibits a more pronounced degradation in its cardiac-based predictions, with its performance dropping significantly compared to the specialized encoders. Furthermore, the contribution of ocular dynamics appears limited: combining EEG and EOG does not yield significant gains over the EEG-only baseline (0.70-0.71), suggesting that EOG provides redundant information within the internal test distribution.

A more direct assessment of incremental neurophysiological information was obtained by comparing U-Sleep ECG+EEG with ECG-only baselines, U-Sleep ECG and Self-DANA ECG (Supplementary Table S16). On SHHS1, macro-average discrimination was largely comparable between ECG+EEG and unimodal configurations. A different pattern emerged under temporal distribution shift on SHHS2 where U-Sleep ECG+EEG showed higher macro-average discrimination than both U-Sleep ECG (ΔC-index = +0.027) and Self-DANA ECG (ΔC-index = +0.039). Point estimates were comparable in the subject-disjoint SHHS2 subset (Supplementary Table S17), with wider intervals reflecting the reduced sample size, indicating that this multimodal advantage was not driven solely by participants included in model development.

A key finding emerges when comparing the two datasets: while SleepFM exhibits a more pronounced decline in performance on the SHHS2 longitudinal set, our proposed framework remains significantly more robust. The overlap-stratified analysis further clarified this pattern. The relative advantage of the U-Sleep-based configurations over SleepFM was retained in the SHHS1-training-disjoint subset, indicating that it was not confined to participants encountered during model training. Across nearly all modality combinations in SHHS2, our approach maintains a stable performance, suggesting that it captures physiological features that are more invariant to the temporal shift between the two study cohorts.

Notably, the consistent underperformance of the Raw + Coupled Mamba model confirms that pre-trained representations remain essential for navigating the complex cross-modal dynamics of overnight recordings.

Comprehensive performance metrics for each cardiovascular outcome are detailed in Supplementary Tables S11 and S12 for C-index, and Supplementary Tables S13 and S14 for five-year AUROC. Absolute-risk calibration at the same horizon is imperfect and configuration-dependent (Supplementary Table S15). Accordingly, model outputs are interpreted primarily as prognostic risk scores for ranking and stratification rather than as calibrated individualized event probabilities.

### Impact of Cross-Modal Integration Approaches

In this final analysis, we investigate the underlying drivers of the observed robustness and performance by isolating the contribution of the Coupled Mamba block. We performed a controlled comparison by substituting this fusion engine with two controlled alternatives while keeping the pretrained embeddings fixed: attention-based modality aggregation followed by a bidirectional LSTM, and late unimodal state-space fusion (Mamba Fusion). Results for the SHHS1 test set and the complete SHHS2 cohort are presented in Figure 6, whereas the corresponding SHHS2 results stratified by SHHS1 training-set overlap are reported in Supplementary Figure S9. Note that Mamba Fusion was evaluated only in multimodal configurations, since with a single input modality it is architecturally equivalent to the corresponding single-branch Coupled Mamba model.

**Fig. 6:**
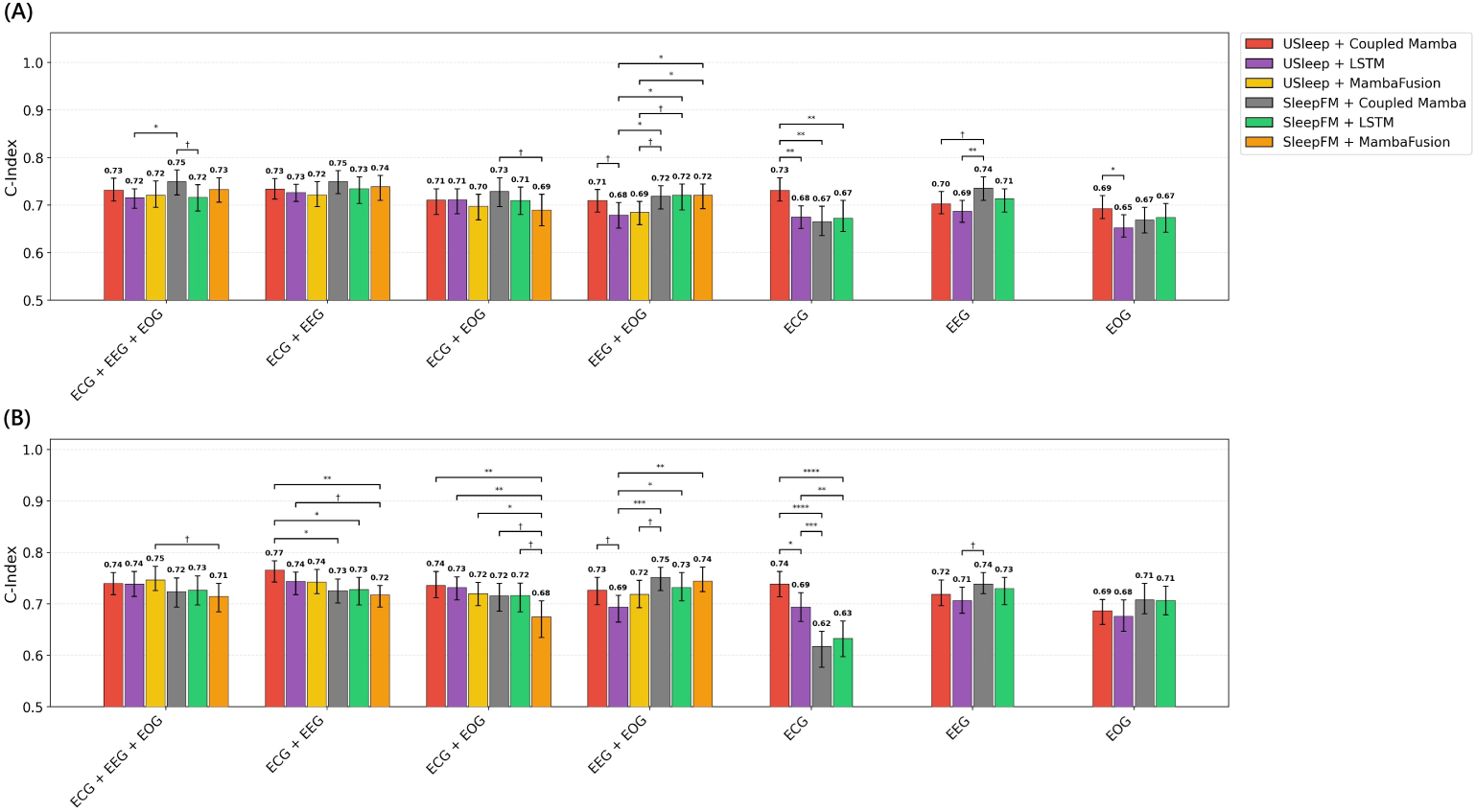
Macro-average C-index performance comparison between fusion architectures. Controlled comparison of Coupled Mamba, attention-based fusion followed by a bidirectional LSTM, and late unimodal state-space fusion (Mamba Fusion), while keeping the feature embeddings (U-Sleep and SleepFM) fixed. To evaluate the engine’s intrinsic capacity to model long-range and cross-modal dependencies, all experiments are conducted across various signal combinations without demographics. Results are reported for **(A)** the SHHS1 internal test set and **(B)** the SHHS2 within-cohort temporal validation set. Error bars represent 95% confidence intervals derived from 100 bootstrap iterations. Statistical significance between model configurations: † p *≤* 0.1; * p *≤* 0.05; ** p *≤* 0.01; *** p *≤* 0.001; **** p *≤* 0.0001

The experimental evidence shows that Coupled Mamba consistently outperforms or matches the LSTM equivalent, regardless of the input embeddings used. In SHHS1, replacing the LSTM with Coupled Mamba using the SleepFM backbone leads to a significant performance boost in the full multimodal setting. Under SHHS2 temporal evaluation, architecture choice remained non-neutral: the U-Sleep + LSTM configuration exhibits a more visible decline compared to the Coupled Mamba setting.

Relative to Mamba Fusion, Coupled Mamba showed a consistent advantage on the SHHS1 test set across multimodal configurations. Under SHHS2 temporal-shift evaluation, the most consistent Coupled Mamba advantage emerged in the ECG+EEG setting: this mirrors the modality-ablation results, where ECG+EEG provided incremental value under temporal shift relative to ECG-only baselines.

Importantly, macro-average performance in the SHHS1-training-disjoint subset remained comparable to that observed in the complete SHHS2 cohort across modality and fusion configurations, with no systematic performance degradation. The relative patterns among encoder and fusion strategies were also generally preserved, indicating that the overall SHHS2 results were not confined to participants encountered during model training. This suggests that while specialized supervised encoders provide the primary foundation for stability, the synchronized cross-modal fusion logic further assists in preserving these features across the longitudinal follow-up. Even under an identical end-to-end raw-encoder protocol, Coupled Mamba improved macro-average discrimination over LSTM on both SHHS1 and SHHS2 (Supplementary Tables S18 and S19).

### Primary and Secondary Prevention Analysis

To further evaluate the clinical utility of our framework, we conducted a stratified analysis distinguishing between primary prevention and secondary prevention cohorts. As detailed in Supplementary Figures S10, S11, S12, discriminative performance was consistently higher in the primary prevention group across all experimental configurations. A similar trend was observed in the within-cohort SHHS2 temporal validation set, where the framework demonstrated greater robustness in the primary prevention subset (Supplementary Figures S13, S14, S15). The superior performance in primary prevention suggests that overnight physiological signatures, such as sleep macrostructure and autonomic regulation, are highly effective markers for identifying early-stage cardiovascular vulnerability before major events occur. In the context of secondary prevention, the overall decline in C-index across all models indicates that predicting recurrent acute events is inherently more complex. This performance gap is likely attributable to the increased physiological noise and the burden of confounding comorbidities typically associated with established cardiovascular disease.

## Discussion

In this study, we investigated whether long-term CVD risk can be predicted from full-night PSG recordings through a modular framework that combines modality-specific representations with continuous cross-modal fusion. We adopt Coupled Mamba as an existing long-sequence fusion module that makes continuous overnight cross-modal modeling computationally tractable. Our results support four main findings. First, we identify strong demographic confounding in the SHHS dataset, which represents a widely used resource for investigating the influence of sleep on cardiovascular health [10, 13], although this issue is seldom addressed. This confounding, specifically related to age, can substantially inflate survival-prediction performance when demographic variables are introduced into the model without accounting for their role as statistical shortcuts that obscure physiological signals’ contributions. Second, under this confounder-controlled setting, we quantify how much prognostic information full-night PSG carries. The cardiac channel accounts for most of the discriminative signal on the internal test set, whereas neurophysiological information, particularly EEG, contributes mainly to preserving performance under temporal distribution shift across recording periods. Third, we demonstrate that Coupled Mamba represents an effective fusion mechanism for full-night PSG analysis, providing a scalable and modular pathway for multimodal CVD risk assessment. Lastly, we show that supervised unimodal representations, particularly those derived from sleep-staging encoders, provide a competitive and robust alternative to complex SSL multimodal pre-training.

A central motivation of this work was to evaluate whether the inherent generalizability attributed to SSL-derived representations truly outperforms specialized features from supervised models when applied to the prediction of long-term cardiovascular outcomes under data-abundant supervision. Recent foundation models for sleep analysis have demonstrated the value of aligning heterogeneous physiological signals through SSL objectives. However, these approaches generally learn cross-modal interactions over short temporal windows and then rely on downstream recurrent or temporal heads to aggregate overnight information. Our findings demonstrate that a modular temporal fusion of specialized unimodal encoders constitutes a highly effective alternative to the multimodal pre-training paradigm. Downstream sample-efficiency analyses further indicated that the supervised pipeline remained competitive under reduced CVD-label availability, although zero-shot and cross-task transfer, where SSL is expected to have its main advantage, were not evaluated here. This indicates that cross-modal interactions can be captured at a later stage effectively and that representations learned from a supervised sleep-staging task can retain clinically relevant information beyond sleep-stage classification itself. This result is physiologically plausible. Sleep staging requires the encoder to capture macrostructural organization, transitions between vigilance states, cortical oscillatory patterns, and sleep continuity. These features are closely linked to autonomic regulation, metabolic stress, inflammation, blood pressure dynamics, and cardiovascular vulnerability [1–3]. Therefore, although sleep-staging encoders are not explicitly trained to predict CVD outcomes, they may learn intermediate representations that summarize clinically meaningful properties of sleep architecture. Crucially, the robustness of these encoders derives from a synergy between the staging task, linking sleep macrostructure to cardiovascular health, and exposure to large-scale, heterogeneous populations. This is particularly effective for PSGs, as the abundance of labeled staging data eliminates the constraints that typically motivate the use of SSL. The present results suggest that such task-specific supervised encoders can serve as effective physiological experts when coupled with a sufficiently expressive temporal-fusion module.

The comparison with the raw-signal configuration further emphasizes the importance of representation learning. Across experiments, the Raw + Coupled Mamba model underperformed the models based on pre-trained encoders. Nonetheless, when raw end-to-end encoders were held fixed, Coupled Mamba still outperformed LSTM, suggesting that fusion design contributes incremental value even without pretrained representations. However, although Coupled Mamba is well suited for long-range sequence modeling, the complexity and noise of full-night PSG signals make direct end-to-end learning difficult in the available sample size. In other words, the fusion engine alone is not sufficient: robust pretraining, based on large-scale, heterogeneous data, either supervised or self-supervised, remains essential.

The modality ablation experiments provide additional insight into the relative contributions of the physiological sources for CVD risk assessment. The relatively limited value of EOG in the internal test set suggests partial redundancy with EEG-derived sleep-architecture information. Since EOG contributes strongly to the identification of REM sleep and eye-movement dynamics, its prognostic role may already be partially captured by sleep-stage representations derived from EEG. Moreover, the ablation analysis shows that ECG-only configuration achieved comparable or higher performance than when considering other modalities: this should not be interpreted as evidence that cardiac morphology alone, rather than sleep information, completely drives the prediction. In our primary setting, the ECG branch uses a U-Sleep encoder pretrained for sleep-stage classification, with embeddings extracted from the dense segmentation layer immediately preceding the final five-class sleep-stage projection. These representations are therefore task-aligned with sleep-state discrimination and capture overnight sleep-related dynamics from the ECG channel, rather than just cardiac morphology. The comparable performance of Self-DANA indicates that part of this signal is cardiac in nature; the distinction we draw is not that ECG carries no cardiac information, but that ECG-only performance cannot be used to exclude sleep-related contributions. The strong ECG-only performance on SHHS1 indicates that sleep-related cardiac dynamics contain a substantial portion of the prognostic information for long-term CVD risk. However, a different pattern emerged under temporal evaluation on SHHS2. There, integrating EEG with ECG remained competitive with ECG-only baseline. This suggests that the principal value of multimodality in our setting may lie less in maximizing internal-test discrimination and more in preserving performance when the evaluation cohort shifts to a later follow-up visit. One possible interpretation of this asymmetry relates to how the two modalities access sleep state. Although U-Sleep ECG encodes sleep-related information, it does so through autonomic modulation of cardiac rhythm across sleep stages, whereas EEG records the cortical activity that defines sleep states more directly. This distinction concerns the transmission pathway rather than the nature of the encoded information. Between the SHHS1 baseline and the SHHS2 follow-up visit, participants aged, accumulated incident comorbidities, and may have initiated cardiovascular medications; such changes could remodel nocturnal autonomic–cardiac coupling and partially alter the mapping between ECG-derived sleep dynamics and cardiovascular risk that was learned at baseline [28, 29]. Consistent with this interpretation, sleep EEG spectral activity exhibits trait-like intra-individual stability and retains its fingerprint-like character into older adulthood [30]: this suggests that neurophysiological information may help preserve discrimination under temporal shift. However, we emphasize that this interpretation remains hypothetical and is not causally demonstrated.

The architectural comparison between Coupled Mamba, attention-based fusion combined with an LSTM (explored by SleepFM [10]), and late unimodal state-space fusion (Mamba Fusion) clarifies the contribution of the fusion engine under fixed pretrained embeddings. The LSTM comparison shows that an early attention-based fusion followed by classical recurrent overnight modeling is generally less effective than continuous state-space coupling for this full-night setting. Mamba Fusion then isolates a finer question within the state-space family: whether modalities should interact continuously or only after independent temporal modeling. Synchronized coupling is most coherent with settings in which complementary streams must interact over the night. In particular, the ECG+EEG advantage under temporal shift discussed above aligns well with Coupled Mamba, because cardiac-autonomic and cortical routes can inform one another throughout the recording rather than only after separate temporal summaries are formed. Late fusion may instead be more robust when a weaker or partly redundant channel is added, because independent temporal modeling limits how much unstable modality-specific variability enters the shared representation. The contribution of the fusion stage was also evident when SleepFM embeddings were combined with Coupled Mamba, showing that the gains are not restricted to U-Sleep features.

These findings suggest that the fusion mechanism itself contributes meaningfully to performance, even when short-term modality alignment is performed as part of the pre-training task. Coupled Mamba is designed to model long-range dependencies while enabling continuous interaction between modalities. This makes it especially appropriate for PSG, where clinically relevant patterns may unfold over hours.

A major contribution of this work is the detailed analysis of a demographic bias, which helps to better contextualize results reported by recent literature. The SHHS cohort exhibits a strong association between age and CVD outcomes: a demographic-only MLP replicated much of the performance of the full multimodal pipeline, and age is inversely correlated with time-to-event, so that ranking older-earlier against younger-later becomes the most efficient path to high concordance. This structure interacts directly with the CoxPH loss and Harrell’s C-index, both of which are based on ranking individuals according to survival time. In such a setting, a model can obtain high concordance by learning the age distribution rather than extracting independent physiological biomarkers from PSG signals. The demographic-only experiments reinforce this concern, showing that age acts as the dominant shortcut in the SHHS dataset when provided explicitly near the decision layer. This does not mean that age is clinically irrelevant; on the contrary, age is one of the most important CVD risk factors. Nevertheless, multivariable Cox models adjusted for age and other clinical covariates show that PSG scores retain independent associations with multiple CVD endpoints. Complementary analyses predicting chronological age from no-demographics embeddings showed that age was recoverable with moderate agreement, indicating that age-related information is partially encoded in the representations while substantial prognostic signal still resides in the physiological waveforms. Accordingly, since the scientific objective is to evaluate whether PSG waveforms contain independent prognostic information, direct integration of age near the decision layer can obscure the contribution of the physiological signals. For this reason, excluding demographics from the main architecture was a deliberate methodological choice. The objective was not to build the strongest possible clinical risk calculator, but to isolate the predictive value of full-night physiological dynamics. In a deployment-oriented system, demographic and clinical variables would likely need to be incorporated, yet their inclusion must be accompanied by the development of robust frameworks for bias mitigation and confounding control. A more clinically meaningful future strategy would be to move beyond simple covariate integration toward sophisticated architectural conditioning. For instance, adopting techniques such as Feature-wise Linear Modulation (FiLM) [31] would allow demographic priors to dynamically condition the physiological encoders, enabling the model to contextualize overnight signals without being dominated by them. Furthermore, implementing demographic masking or dropout during training could enhance model robustness, forcing the network to extract independent biomarkers from the raw waveforms even when clinical covariates are absent. These strategies, combined with residual-risk modeling (i.e., evaluating the marginal contribution of sleep-derived features over baseline risk), would provide a more rigorous framework to quantify the unique prognostic contribution of the sleep-heart axis.

Clinically, the present framework is best positioned for opportunistic risk stratification rather than standalone screening. Because diagnostic PSG is already acquired in sleep workflows, the same recording can yield a PSG-derived risk score to flag higher-versus lower-risk patients for closer cardiovascular follow-up. The proposed framework also offers distinct practical advantages. Its modular design allows modality-specific encoders to be swapped or updated without retraining a monolithic multimodal foundation model. A modular architecture can integrate EEG, EOG, or ECG experts depending on availability, together with the integration of unconventional modalities (e.g., audio or images), while the fusion module provides a unified mechanism for temporal and cross-modal integration. This flexibility may be especially useful as new foundation models emerge for individual physiological modalities.

From a computational perspective, the linear-time complexity of Coupled Mamba offers significant efficiency gains for full-night sequence modeling. As demonstrated in our computational benchmark, it avoids the severe memory bottlenecks of standard attention mechanisms, remaining scalable even at fine-grained temporal resolutions. However, the overall speed of our best-performing pipeline is currently bottlenecked by the U-Sleep encoders. U-Sleep is a heavyweight architecture that was integrated as a frozen feature extractor without extensive, pipeline-specific optimization or hyperparameter tuning. However, given the proven prognostic robustness of these supervised representations, a highly promising future extension involves knowledge distillation. By distilling the rich, high-dimensional feature maps of U-Sleep into a much smaller, lightweight convolutional network (similar to the compact 1D-CNNs utilized in our raw configuration) we might effectively transfer this physiological expertise. This approach would yield an end-to-end architecture that preserves the high predictive accuracy while operating with the extreme computational efficiency required for deployment on low-power or wearable monitoring devices. Despite these promising results, several limitations should be acknowledged. First, although SHHS is a large and well-characterized cohort, the study population is not fully representative of contemporary clinical sleep populations. The recordings were collected decades ago, and changes in cardiovascular prevention, medication use, diagnostic criteria, and population health may affect generalizability. Second, while SHHS2 enables a within-cohort temporal evaluation, it does not constitute a fully independent external validation. Rigorous evaluation in independently recruited cohorts across diverse populations, clinical sites, and acquisition systems remains necessary before clinical-generalization claims.

In addition, although Coupled Mamba’s input-dependent selection mechanism can selectively propagate or attenuate information over long sequences [14, 16], and may thereby reduce the impact of transient artifacts relative to less selective temporal aggregators, we did not perform dedicated stress tests under controlled noise or sensor-failure conditions. Robustness to arbitrary signal artifacts therefore remains to be established.

Even if the modular design offers flexibility in terms of modality types, the current implementation assumes complete ECG/EEG/EOG availability, as in SHHS, and therefore does not require missing-modality handling at inference. For real-world incomplete montages, further work should include modality dropout during training together with a missing-modality mask, so that absent streams do not contribute to the shared state update. Currently, our framework focuses on EEG, EOG, and ECG to emphasize the brain-heart axis and support low-burden monitoring. However, excluding respiratory effort, airflow, oxygen saturation, and EMG may omit important pathways linking sleep-disordered breathing, sympathetic activation, and cardiovascular outcomes. While the selected modalities are attractive for portable monitoring, future work should quantify how much incremental value is gained by adding respiratory and oximetry signals.

Absolute-risk calibration at 5 years was imperfect across several configurations, particularly without demographic inputs. This is expected in our setting where the models were trained to optimize prognostic ranking rather than calibrated event probabilities. We therefore limit clinical interpretation to risk stratification and ranking, rather than individualized absolute-risk prediction.

Finally, the observational nature of the SHHS cohort prevents causal interpretation. The model identifies associations between PSG-derived patterns and future CVD events, but these associations may reflect unmeasured comorbidities, treatment effects, socioeconomic factors, or other latent variables. Future studies should evaluate whether the learned representations are stable under stronger adjustment strategies and whether they can identify modifiable sleep-related risk factors.

In this work, we demonstrated that full-night PSG recordings contain rich prognostic signatures for long-term CVD risk that can be effectively extracted using unimodal supervised encoders, without relying on multimodal self-supervised pre-training. By coupling these encoders with a Coupled Mamba architecture, we modeled long-range temporal dependencies while enabling continuous interactions between physiological modalities throughout the night. Our findings challenge the recent prevailing SSL paradigm by showing that task-specific supervised encoders provide highly competitive representations for downstream clinical tasks when paired with a sufficiently powerful fusion engine. Learning from long temporal sequences allows the model to capture slow physiological drifts and sleep-cycle effects that windowed SSL objectives, operating on short segments, are structurally unable to access. Model outputs are best interpreted as prognostic risk scores for ranking and stratification rather than as calibrated absolute event probabilities for individualized screening. Furthermore, we emphasize the critical need for rigorous demographic analysis in sleep-related survival modeling. We showed that chronological age acts as a dominant statistical shortcut in the commonly employed SHHS dataset, necessitating a no-demographics evaluation approach to isolate the predictive value of raw physiological signals. The modularity of our framework offers a flexible pathway for future clinical integration, allowing for the seamless swapping of modality experts and adaptation to low-burden, portable monitoring devices. Future research should focus on refining bias mitigation strategies and exploring the incremental value of additional respiratory channels within this state-space modeling paradigm.

## Methods

### Dataset

The SHHS is a multi-center prospective cohort study from the NSRR designed to explore the association between sleep architecture and CVD outcomes. The dataset consists of two subsets: SHHS1 (baseline) and SHHS2 (follow-up). During the first visit, conducted between 1995 and 1998, 6441 participants were enrolled, and approximately half of this cohort (3295) underwent a second visit about 5-8 years later. Full-night PSG was performed during both visits. The recorded montage includes two bipolar EEG channels (C4-A1 and C3-A2), two EOG channels (left and right), one ECG bipolar lead, and two leg electromyographic channels. Additional physiological data were captured via pulse oximetry, airflow nasal-oral sensors, and respiratory excursions. Major CVD outcomes were monitored for over a decade longitudinally from the baseline assessment [25, 26].

In this study, the SHHS1 cohort is utilized for model development, while SHHS2 is reserved exclusively for within-cohort temporal validation. The initial dataset comprised 5793 and 2651 recordings for SHHS1 and SHHS2, respectively. After excluding participants with null follow-up durations or entirely missing demographics information, the final study population consists of 5774 subjects in SHHS1 and 2618 in SHHS2. To ensure a rigorous and fair comparison with current state-of-the-art methods, we retained the SleepFM [10] test partition for SHHS1, as that work serves as a primary benchmark for our task. The remaining SHHS1 participants were divided at the participant level into training and validation sets using an 80/20 split stratified using multilabel stratified sampling to preserve the joint distribution of the six cardiovascular endpoints, binned event times, and demographic variables (i.e., sex, age, BMI, medication use). Since SHHS2 represents a repeat visit from the same parent cohort, participant identifiers were used to quantify overlap with the SHHS1 training set. Performance was therefore evaluated in the complete SHHS2 cohort and separately in the SHHS1-training-overlap and SHHS1-training-disjoint subsets.

Participants with a history of CVD events prior to the baseline assessment were not excluded, so that the study population reflects both primary prevention (individuals without prior CVD events) and secondary prevention (individuals with established CVD) scenarios. This choice aligns with the methodology established by Thapa et al. [10] and is further motivated by the clinical observation that the considered outcomes are primarily acute events that may reoccur. To evaluate the clinical utility of our framework across different risk profiles, we conduct separate performance analyses for primary and secondary prevention. Specifically, the former cohort comprises participants who were CVD-free at baseline, while the latter includes those with a prior history of CVD. These subgroup evaluations are performed across both the SHHS1 test set and the entire SHHS2 cohort.

### CVD Outcomes and Problem Formulation

The prediction of long-term CVD risk is formalized as a multi-label survival analysis task, employing EEG, EOG, and ECG signals as input modalities. The exclusion of other traditional PSG modalities, such as respiratory belts and EMG, is motivated by the physiological centrality of the brain-heart axis in CVD risk [32, 33], together with the general clinical objective of enabling portable, low-burden longitudinal monitoring without affecting the model’s ability to capture systemic autonomic dysregulation.

Following the framework established by SleepFM [10], the model simultaneously estimates the risk of onset for six distinct CVD outcomes: Angina, CVD Death, CHF, CHD Death, MI, and Stroke. For each subject *i* and event *k*, the target is represented by a pair (*T_i,k_, δ_i,k_*), where *T_i,k_* denotes the time-to-event (or time-to-censoring if the event does not occur) and *δ_i,k_ ∈ {*0, 1*}* serves as the event indicator. In this context, the model aims to estimate a prognostic risk score *h_i,k_* for each event. This score represents the cumulative hazard, reflecting the subject’s instantaneous susceptibility to a specific CVD event onset given their physiological state during the sleep recording. Rather than a simple classification, this approach treats cardiovascular risk as a prognostic ranking problem: the model learns to assign higher risk scores to subjects who experience events earlier within the longitudinal follow-up window.

We opted for joint multi-endpoint optimization to maintain alignment with the current foundation-model benchmark (e.g., SleepFM [10]).

### Missing Data Handling

Analysis of the CVD outcomes revealed that several subjects were recorded as having experienced an event despite lacking a documented onset date. This discrepancy was primarily concentrated in the Angina label, with only rare instances identified among some of the other CVD outcomes. Specifically, in the SHHS1 cohort, 1838 out of 2198 recorded Angina events (83.6%) lack a documented time-to-event, and this proportion was even higher in SHHS2, reaching 90.8% (982 of 1081 events). Unfortunately, the mechanism underlying the missing event times could not be determined from the publicly available SHHS documentation. Figure 1 shows the frequency distribution of follow-up times across the six CVD labels, distinguishing participants with no event, events with a documented onset date, and events whose onset time is missing. For most endpoints, documented events are relatively few and spread over follow-up, whereas the no-event group concentrates near the end of the observation window. For Angina, missing onset times dominate the event group. While imputing these missing values with the patient’s censoring date (i.e., the time from baseline to the last known contact or death) represents a potential strategy, such an approach significantly distorts the temporal distribution for this specific outcome, as illustrated in Figure 1. For this reason, we adopt a masking strategy for these “uncertain” subjects. Instead of discarding these participants entirely, which would result in a substantial loss of informative data for other concurrently modeled CVD outcomes, we exclude them from both the loss function and performance metric calculations only for the specific labels where the onset time is missing. This selective masking allows the model to leverage the physiological information of these patients for documented outcomes while avoiding the introduction of bias through arbitrary imputation. Following the implementation of this masking strategy, the final distribution of cardiovascular events for the SHHS1 test and SHHS2 cohorts is detailed in Table 1. Supplementary Table S20 provides a more granular stratification of the SHHS1 test set and SHHS2 by primary and secondary prevention, with SHHS2 additionally divided according to participant overlap with the SHHS1 training set.

### Demographic Analysis

In the context of CVD risk prediction, variables such as age, gender, BMI (Body Mass Index), and medications known or suspected to alter sleep or CVD risk should be taken into account [23]. While we initially considered adopting the strategy used by SleepFM [10], where these variables are integrated near the decision layer, we argue that including such demographic priors would, in this way, likely encourage the model to exploit them as statistical ‘shortcuts’ rather than learning to isolate independent prognostic biomarkers from the raw physiological waveforms. To prevent this over-reliance and to rigorously assess the prognostic value of raw data, we made a strategic design choice to exclude all demographic variables from the final architecture. By restricting the input to raw signals, our objective is to evaluate how much prognostic information can be recovered from PSG signals when demographic variables are not provided explicitly near the decision layer. To support this decision, we performed an extensive preliminary investigation of the cohort’s demographic and temporal distributions to identify inherent biases that could act as statistical shortcuts. Nonetheless, to empirically quantify the impact of these demographic features, we conducted a series of supplementary experiments incorporating these variables into our framework. Specifically, we evaluated our model across four configurations: (i) all demographic integration (including age, gender, BMI, and medications); (ii) all demographics excluding age; (iii) age and gender only; and (iv) gender only. The latter two configurations were specifically designed to facilitate a direct and fair comparison with the original SleepFM framework [10], while the isolated removal of age allows us to quantify the contribution of chronological age when provided as an explicit model input, without excluding that age-related information could be indirectly present in the physiological waveforms.

### Age Estimation Analysis

To assess whether chronological age remains implicitly encoded in overnight physiological representations when demographic variables are not provided as model inputs, we performed a dedicated age-estimation analysis. We used the same full-modality encoder configurations as in the main survival experiments (U-Sleep and SleepFM), without demographic inputs, and the same Coupled Mamba temporal-fusion backbone and hyperparameters. Instead of predicting cardiovascular outcomes, the model was trained to regress chronological age from the fused overnight representations. Models were trained on the SHHS1 training cohort, and performance evaluated on the SHHS1 test set and subsequently on the complete SHHS2 cohort. Agreement between predicted and chronological age was summarized using the coefficient of determination (*R*^2^).

### Model Architecture and Theoretical Background

Our proposed framework follows a modular design consisting of three main stages: modality-specific feature extraction, continuous cross-modal fusion through Coupled Mamba, and a survival analysis head for multi-label risk prediction (Figure 2).

To evaluate the impact of different representation learning strategies, we explore two distinct approaches for the encoder stage, which processes the raw physiological signals (*x_EEG_, x_EOG_, x_ECG_*):

- we implement a lightweight architecture composed of modality-specific convolutional layers designed to extract informative patterns directly from the raw waveforms. These architectures are adapted from established literature [34–36] and refined to align with the temporal requirements of our framework. Specifically, the ECG encoder utilizes a 1D-CNN backbone to capture essential cardiac morphological information [34]. The EOG encoder employs a dual-branch structure designed to independently characterize rapid eye movements and slow oscillations [35], while the EEG encoder integrates both raw temporal sequences and FFT-derived spectral information to capture multi-scale cortical dynamics [36]. To ensure a synchronized multi-modal representation, all encoders were calibrated to produce one feature token every 5 seconds, following the temporal resolution adopted in SleepFM [10]. These encoders are trained from scratch alongside the rest of the network, allowing the convolutional filters to optimize specifically for the downstream survival analysis task;
- we utilize unimodal modality-specific encoders pre-trained on large-scale datasets. For each physiological stream, we employ a modality-specific instance of the U-Sleep architecture [20], with weights pre-trained to predict sleep stages from that specific single modality. To ensure rigorous evaluation and avoid data leakage, all SHHS records were excluded from the pre-training cohort: all the other data used is detailed in the original work [21]. Embeddings are extracted from the dense segmentation layer, located immediately before the final projection, leading to a 6-dimensional embedding at a resolution of one token per second [20]. To maintain temporal consistency with the 5-second framework, a concatenation layer is applied across five consecutive time steps. For modalities involving multiple channels (i.e., EEG and EOG), the embeddings extracted from each individual channel are aggregated via average pooling to produce a single representative vector. Additionally, we evaluate a specific hybrid configuration where the Self-DANA encoder [24] is used exclusively to replace the ECG branch, while the EEG and EOG branches remain consistent with the U-Sleep setup. Self-DANA produces a 768-dimensional embedding for every 5-second window.

Full-night PSG sequences contain structure at multiple timescales, from transient waveform events to dynamics that unfold over hours. Convolutional encoders extract local and hierarchical features efficiently, and recurrent networks can carry a hidden state across time, but recurrence remains sequential and therefore limits parallelization over long recordings [37, 38]. Transformers overcome this sequential bottleneck by modeling direct interactions between distant elements in parallel, yet their computational and memory cost scale quadratically with length [14, 15]. Sparse or factorized attention and latent bottlenecks reduce this cost, typically by restricting the pairwise interactions that make full attention effective [14, 39, 40]. While this bottleneck is manageable for short sequences of 30-second sleep epochs, it becomes computationally prohibitive for synthesizing an entire eight-hour PSG recording. We therefore implement fusion with Coupled Mamba [16], a multimodal extension of selective SSMs that processes the synchronized encoder sequences jointly over the full night at linear cost in sequence length.

An SSM summarizes the input history in a latent state *h_t_* whose discrete update is linear in sequence length,

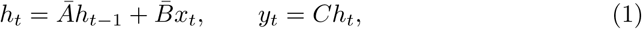

after discretization of a continuous-time linear system [14, 37]. Classical structured SSMs keep these maps time-invariant: they retain a compressed long history, but cannot decide from the content of the current token which information to keep or discard. Mamba replaces this with a selection mechanism that makes the discretization step and the input/output projections functions of *x_t_*, so that the state can propagate or suppress information dynamically [14]. Input dependence precludes the convolutional training used by time-invariant SSMs; a hardware-aware parallel scan restores efficient training while retaining Transformer-level sequence modeling with linear memory scaling [14]. This combination (i.e. content-dependent long-range memory without quadratic attention) is what makes continuous full-night fusion computationally tractable.

Coupled Mamba extends this framework to multimodal sequences by enabling interactions between modality-specific state chains [16]. Rather than integrating modalities only at the input or decision level, it introduces an inter-modal state-transition mechanism in which the state of modality *m* depends on its current input and on the preceding states of all *M* modality chains:

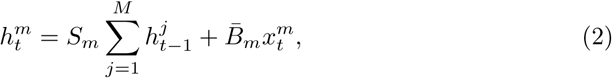

where *S_m_*governs the transition of the aggregated historical state for modality *m*. By combining modality-specific state evolution with continuous exchange of historical information, Coupled Mamba provides a unified mechanism for long-range temporal modeling and intermediate multimodal fusion. This formulation is particularly suited to our objective of integrating heterogeneous physiological representations continuously across a complete recording, while allowing each modality to retain its own evolving state.

The interaction layer is designed to process the synchronized multimodal sequences through a stack of multiple Coupled Mamba blocks. Since the encoders could produce embeddings of varying sizes, each modality is first projected into a shared latent space of dimension *d_model_* using modality-specific linear layers. Each block incorporates skip connections and a normalization layer to stabilize gradient flow and preserve modality information. Given the causal nature of the SSMs, each hidden state at time *t* encodes the previous physiological trajectory. Therefore, to aggregate the output sequences of each modality into fixed-size global vectors (*d_model_*) we can use either mean pooling or last token extraction. To empirically validate the efficiency of this formulation for full-night sequences, we conducted a computational benchmark against standard cross-attention (Supplementary Figure S16). Results confirmed that Coupled Mamba yields a substantially lower memory usage and scales linearly, preventing the out-of-memory failures typically encountered by attention mechanisms at high temporal resolutions.

The prediction head maps the integrated features to the risk of six CVD outcomes. The pooled vectors from the EEG, EOG, and ECG branches are concatenated into a single global descriptor, followed by a dropout layer to enhance generalization. Finally, a multi-label linear head projects the concatenated representation to K-label output neurons. Each value represents the predicted risk for a specific cardiovascular event.

### Data Preparation

In both SHHS1 and SHHS2, we used the same SHHS montage. EEG was analyzed using C4–A1 and C3–A2, EOG using ROC–A1 and LOC–A2, and ECG using a single bipolar chest lead. This ECG configuration is not a standard clinical 12-lead setup; electrodes were placed approximately 3–5 cm below the midpoint of the right and left collarbones, in the intercostal spaces, following the SHHS recording protocol. Because this montage was fixed across included recordings, all analyzed studies contained two EEG channels, two EOG channels, and one ECG channel. We therefore did not apply modality-specific missing-channel handling, channel imputation, or modality-dropout strategies. Preprocessing was implemented according to the input requirements of each encoder backbone, while preserving a common five-second tokenization protocol for full-night modeling. In SHHS, EEG was acquired at 125 Hz, EOG at 50 Hz and ECG at 125 Hz in SHHS1 and 250 Hz in SHHS2. For Self-DANA, ECG was upsampled to 500 Hz using cubic-spline interpolation, segmented into non-overlapping five-second windows, mean-centered, and filtered with a fifth-order moving-average filter and a fourth-order Butterworth band-pass filter (0.5–40 Hz). For U-Sleep, signals were resampled to 128 Hz using polyphase filtering and channel-wise per-subject rescaled to median 0 and IQR 1. For SleepFM and the raw end-to-end configuration, recordings were upsampled to 128 Hz using linear interpolation and standardized to zero mean and unit variance. This information, including the reproduced SleepFM comparator, is summarized in Supplementary Table S21.

### Training Details

Training was performed using the AdamW optimizer coupled with a One Cycle learning rate scheduler. The models were trained for a maximum of 100 epochs; early stopping was applied if the validation loss failed to show any further decrease for 15 consecutive epochs to prevent overfitting. To identify the optimal configuration for the Coupled Mamba framework, an extensive hyperparameter tuning phase was conducted. This search was performed exclusively on the hybrid pre-trained configuration (utilizing U-Sleep for EEG/EOG and Self-DANA for ECG) as it represents the most heterogeneous input scenario. Given the significant dimensional difference between USleep and Self-DANA embeddings, tuning the Coupled Mamba block on this setup ensured that the interaction layers were optimized to handle diverse representational scales. In this way, we guaranteed that the framework remained robust when applied to end-to-end or entirely U-Sleep-based setups, maintaining a fair evaluation environment where the only variable is the nature of the feature extraction. The specifications of the explored and selected parameters are detailed in Supplementary Table S22.

Following the methodology established in SleepFM [10], we utilize the Cox Proportional Hazards (CoxPH) loss function. It maximizes the partial likelihood and, for each single label *k*, is defined as:

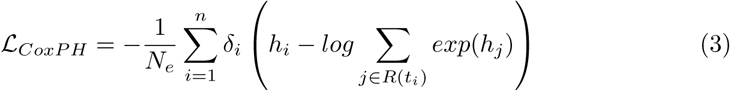

where *h_i_*is the predicted hazard for the *i*th patient, *δ_i_* is the event indicator (1 if occurs, 0 otherwise), *t_i_* represents either the time-to-event or the censoring time, *R*(*t_i_*) is the risk set of all patients with event times *≥ t_i_*, *n* the total number of patients, and *N_e_* the number of events. To accommodate the multi-label nature of the task, SleepFM extends this loss by computing it independently for each outcome and summing the results, obtaining the final loss as:

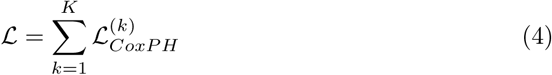

where *K* represents the total number of labels, i.e., six CVD outcomes. Moreover, to address the class imbalance of the datasets, we introduce a weighting factor for each label. These weights are calculated to be inversely proportional to the frequency of events for each outcome.

### Evaluation Metrics

To evaluate the discriminative performance of our models, we primarily utilize Harrell’s concordance index (C-index), the standard metric for survival analysis. The C-index quantifies the model’s ability to correctly rank subjects according to their risk of an event over the entire follow-up period. It is formally defined as:

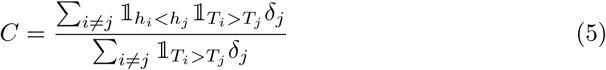

where *h* represents the predicted risk score, *T* denotes the observed time (either event time or censoring time), and *δ_j_* is the event indicator.

Because Harrell’s C-index evaluates ranking rather than absolute-risk accuracy, we additionally assessed discrimination and calibration at a clinically meaningful 5-year horizon. Time-dependent discrimination was quantified using cumulative dynamic AUROC computed from model-derived risk scores. To obtain horizon-specific event probabilities, risk scores were mapped to survival functions through a Cox proportional hazards formulation with a Breslow-estimated baseline hazard [41] fitted on the SHHS1 training set and applied unchanged to the SHHS1 test set and the SHHS2 cohort. Breslow’s approach uses a non-parametric maximum likelihood estimation of the cumulative baseline hazard function:

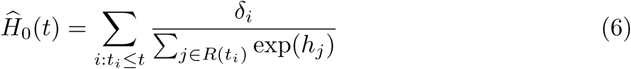

where *δ* represents the status indicator (1 if event, 0 otherwise), *h* the predicted risk score and *R*(*t_i_*) the risk set of all patients with event times *≥ t_i_*.

Using this estimation, subject-specific survival probabilities are computed as:

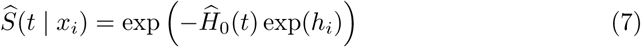

and horizon-specific event probabilities are, consequently, obtained as:

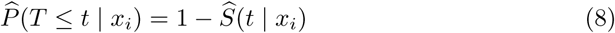

Calibration at 5 years was summarized by calibration slope and calibration-in-the-large (CITL) [42], together with the Index of Prediction Accuracy (IPA) [43] based on the censoring-adjusted Brier score. Ideal calibration corresponds to CITL near 0 and slope near 1; positive IPA indicates improved overall accuracy relative to a null model.

### Ablation Studies

In addition to the demographic sensitivity analysis previously described, we perform a series of ablation studies to systematically evaluate the influence of data inputs and architectural choices on the model’s performance. First, we analyze the input modalities’ contributions by training and testing the framework on individual signals (ECG, EEG, EOG) and their various combinations (e.g., ECG+EEG, EEG+EOG). These experiments were performed exclusively in a “no-demographics” configuration to ensure that the observed prognostic value is derived from the overnight waveforms only. This approach allows us to identify signal-specific contributions and potential cross-modal redundancies or synergies. To isolate the contribution of the fusion engine, we performed controlled architectural comparisons while keeping the pretrained feature embeddings fixed. Specifically, Coupled Mamba was compared against an attention-based modality aggregation followed by a bidirectional LSTM (as used in the original SleepFM pipeline [10]), and a late-fusion state-space baseline, in which each modality is processed independently by a modality-specific Mamba backbone and the resulting representations are combined only at the decision stage. By freezing the representational foundation, we ensure that any variance in discriminative performance is strictly attributable to the engine’s capacity to model long-range and cross-modal dependencies.

### Downstream Sample-Efficiency Analysis

To assess performance under reduced data availability, we conducted a controlled sample-efficiency analysis under matched downstream conditions. Starting from the primary no-demographics, full-modality setting, we retrained both the U-Sleep + Coupled Mamba and SleepFM + LSTM pipelines on reduced fractions of the SHHS1 training cohort (10%, 20%, and 50%) while keeping the SHHS1 test partition unchanged. For each training fraction, five independently sampled training subsets were drawn from the original SHHS1 training set, and models were trained separately on each subset. Hyperparameters, survival objective, endpoint definitions, and evaluation metrics were held fixed relative to the main experiments. This analysis evaluates downstream sample efficiency under supervised adaptation to the CVD survival task.

## Declarations

### Data availability

The Sleep Heart Health Study (SHHS) is publicly available and can be accessed at https://sleepdata.org/datasets/shhs.

### Code availability

The underlying code for this study is available in MambaSleepCVD repository and can be accessed at https://github.com/alessiacalzoni/MambaSleep-CVD

## Acknowledgments

A.C. was funded by the European Union-NextGeneration EU, Mission 4 Component C2 CUP D82B23004220008. The funder played no role in study design, data collection, analysis and interpretation of data, or the writing of this manuscript.

The Sleep Heart Health Study (SHHS) was supported by National Heart, Lung, and Blood Institute cooperative agreements U01HL53916 (University of California, Davis), U01HL53931 (New York University), U01HL53934 (University of Minnesota), U01HL53937 and U01HL64360 (Johns Hopkins University), U01HL53938 (University of Arizona), U01HL53940 (University of Washington), U01HL53941 (Boston University), and U01HL63463 (Case Western Reserve University). The National Sleep Research Resource was supported by the National Heart, Lung, and Blood Institute (R24 HL114473, 75N92019R002).

## Author contributions

Conceptualization: A.C.; Data curation: A.C., A.D.R., G.M., L.F.; Formal analysis: A.C., A.D.R., M.S., G.M., B.Z.; Funding acquisition: A.S.; Investigation: A.C., A.D.R., G.M., B.Z., M.S.; Methodology: A.C., A.D.R., G.M., B.Z., M.S.; Project administration: A.S., F.D.F.; Resources: M.S., A.S.; Software: A.C.; Supervision: A.S., F.D.F.; Validation: A.C.; Visualization: A.C., M.S.; Writing - original draft: A.C.; Writing - review & editing: A.C., A.D.R., G.M., B.Z., L.F., M.S., F.D.F., A.S.

