## supplementary material for "MambaSleepCVD for prediction of long term cardiovascular outcomes from polysomnography"

### Supplementary figures

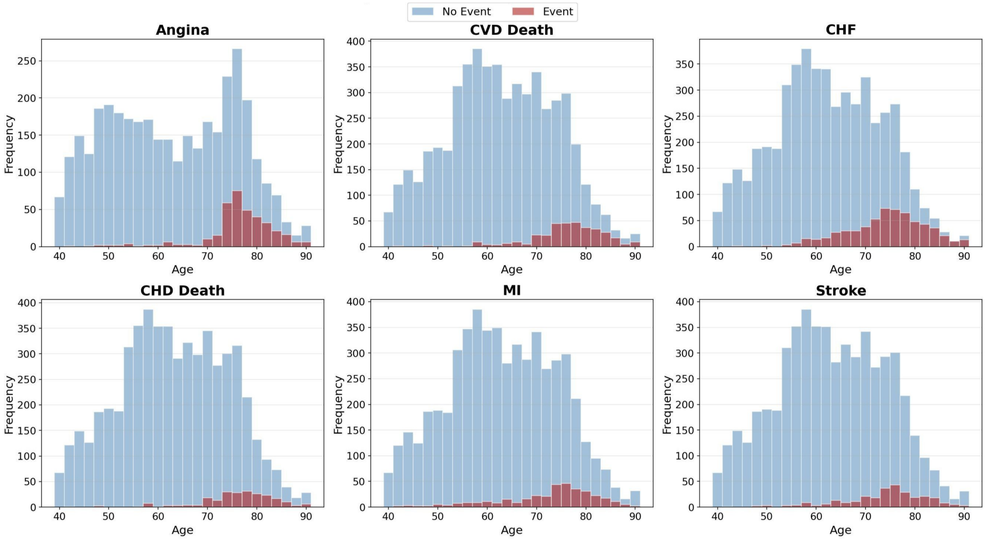


**Fig. S1**: Age distribution in SHHS1 across CVD outcomes. Histograms comparing the age frequency of subjects who did not experience an event (blue) with those who did (red) for each of the six evaluated labels. This plot highlights how CVD outcomes are heavily concentrated in the elderly population, providing visual evidence for the significant age-related distribution shift identified in the study population.

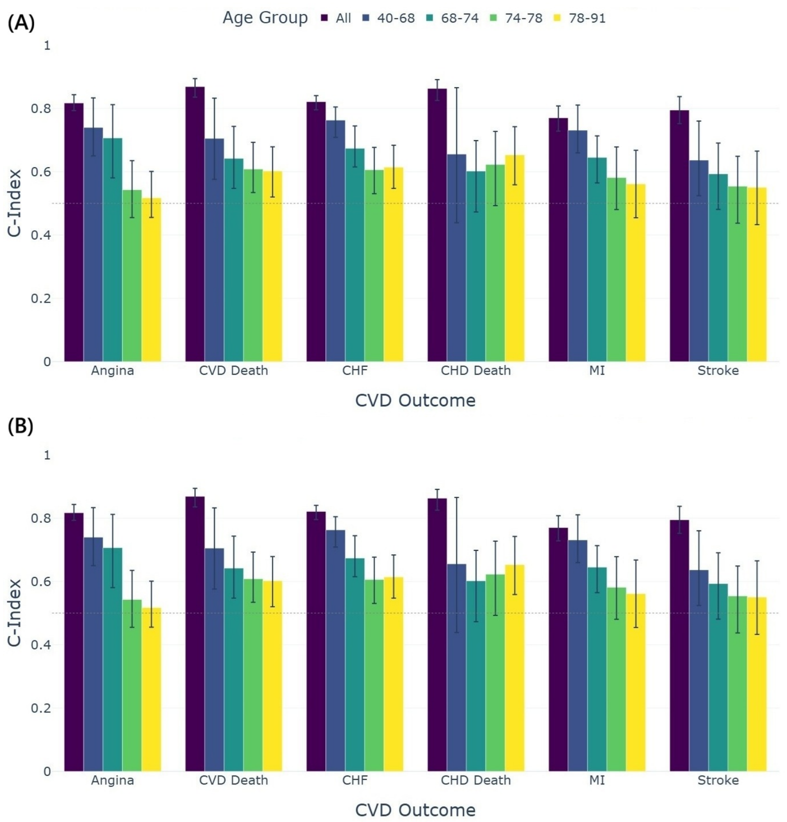


**Fig. S2**: C-index performance degradation across age-homogeneous subgroups. Analysis of the predictive capacity of **(A)** the SleepFM replica and **(B)** the demographic MLP baseline (i.e. age and gender) when evaluated within restricted age intervals. The systematic decline of the C-index toward the 0.5 baseline in narrower age groups mathematically confirms that biological age acts as the dominant statistical shortcut for risk ranking in the SHHS dataset. Error bars represent 95% confidence intervals derived from 100 bootstrap iterations.

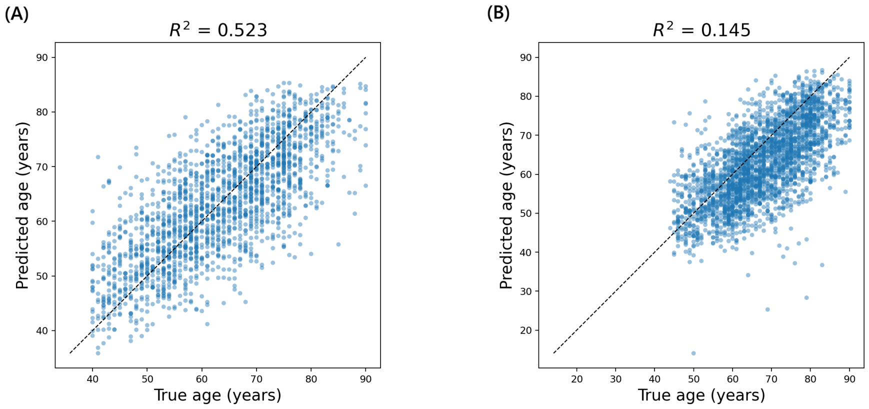


**Fig. S3**: Age prediction from U-Sleep representations. Predicted versus true chronological age obtained by regressing age from full-modality U-Sleep embeddings (ECG, EEG, and EOG) using the Coupled Mamba temporal backbone, without demographic inputs. **(A)**: SHHS1 test set ($R^2 = 0.523$). **(B)**: SHHS2 within-cohort temporal evaluation set ($R^2 = 0.145$). The dashed line indicates the identity line (perfect prediction)

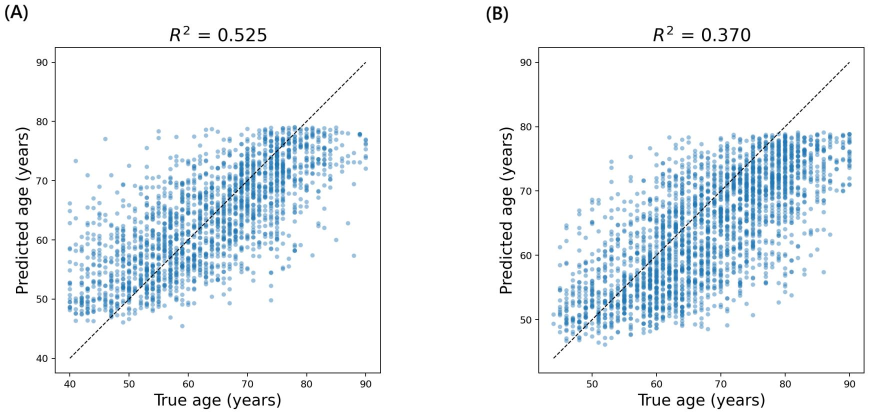


**Fig. S4**: Age prediction from SleepFM representations. Predicted versus true chronological age obtained by regressing age from full-modality SleepFM embeddings (ECG, EEG, and EOG) using the Coupled Mamba temporal backbone, without demographic inputs. **(A)**: SHHS1 test set ($R^2 = 0.525$). **(B)**: SHHS2 within-cohort temporal evaluation set ($R^2 = 0.370$). The dashed line indicates the identity line (perfect prediction)

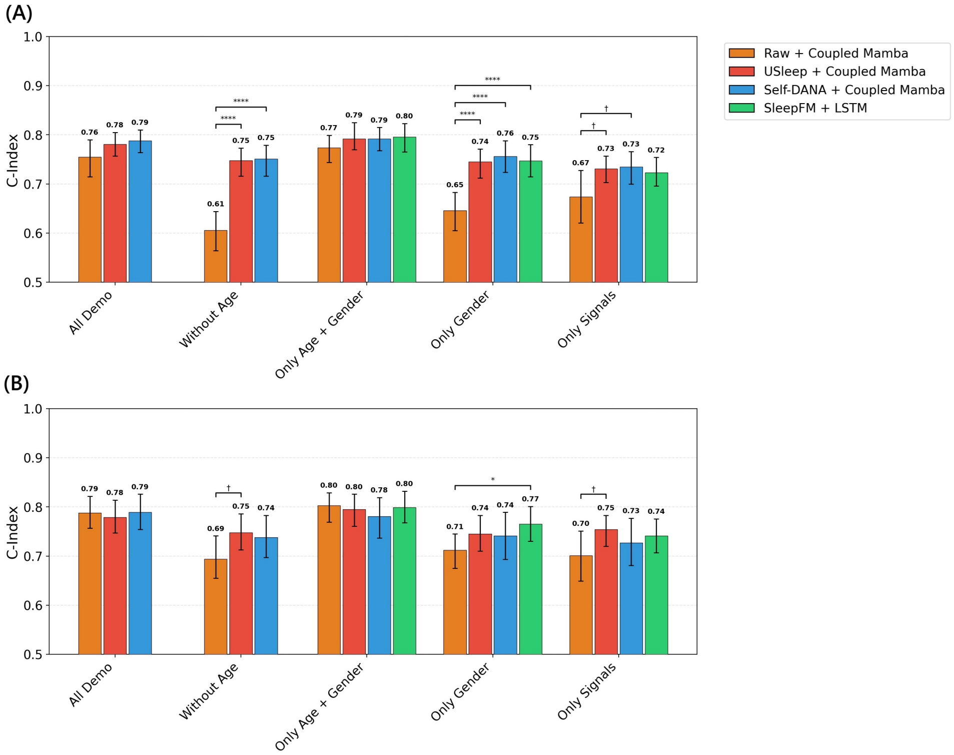


**Fig. S5**: C-index macro-average performance in SHHS2 stratified by SHHS1 training-set overlap across demographic configurations. Stratified performance on the SHHS2 follow-up cohort for participants overlapping with **(A)** the SHHS1 training set versus **(B)** the SHHS1-training-disjoint subset. Error bars represent 95% confidence intervals derived from 100 bootstrap iterations. Statistical significance between model configurations: † $p \leq 0.1$; * $p \leq 0.05$; ** $p \leq 0.01$; *** $p \leq 0.001$; **** $p \leq 0.0001$

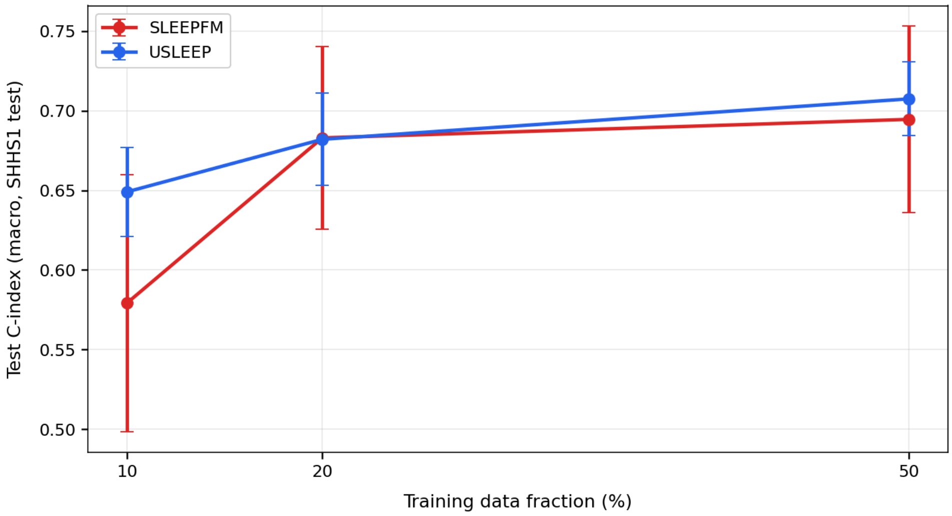


**Fig. S6**: Downstream sample efficiency on SHHS1 (macro-average). Macro-average C-index for U-Sleep + Coupled Mamba and SleepFM + LSTM trained on 10%, 20%, or 50% of the SHHS1 training set (no demographics, all modalities), keeping the test partition fixed. Error bars denote variability (± standard deviation) across five independently sampled training subsets.

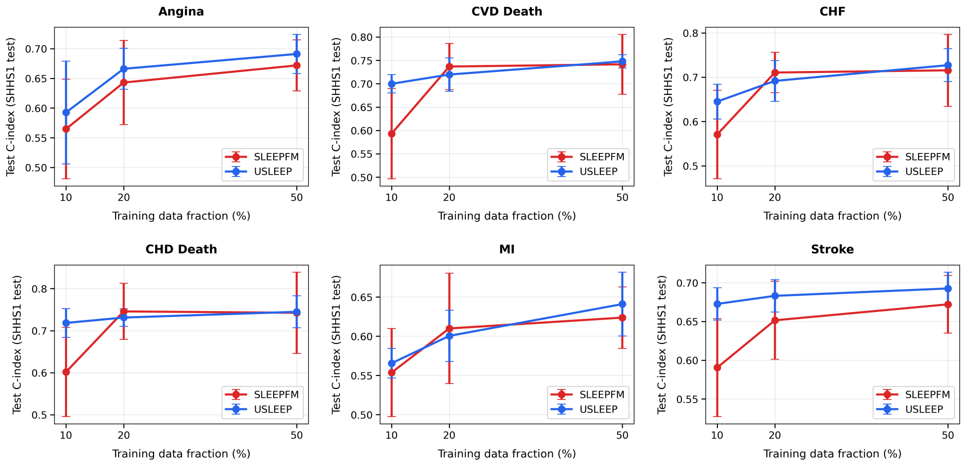


**Fig. S7**: Downstream sample efficiency on SHHS1 (endpoint-specific). Endpoint-specific C-index under the reduced-training protocol, keeping fixed the test set. Error bars denote variability (± standard deviation) across five independently sampled training subsets.

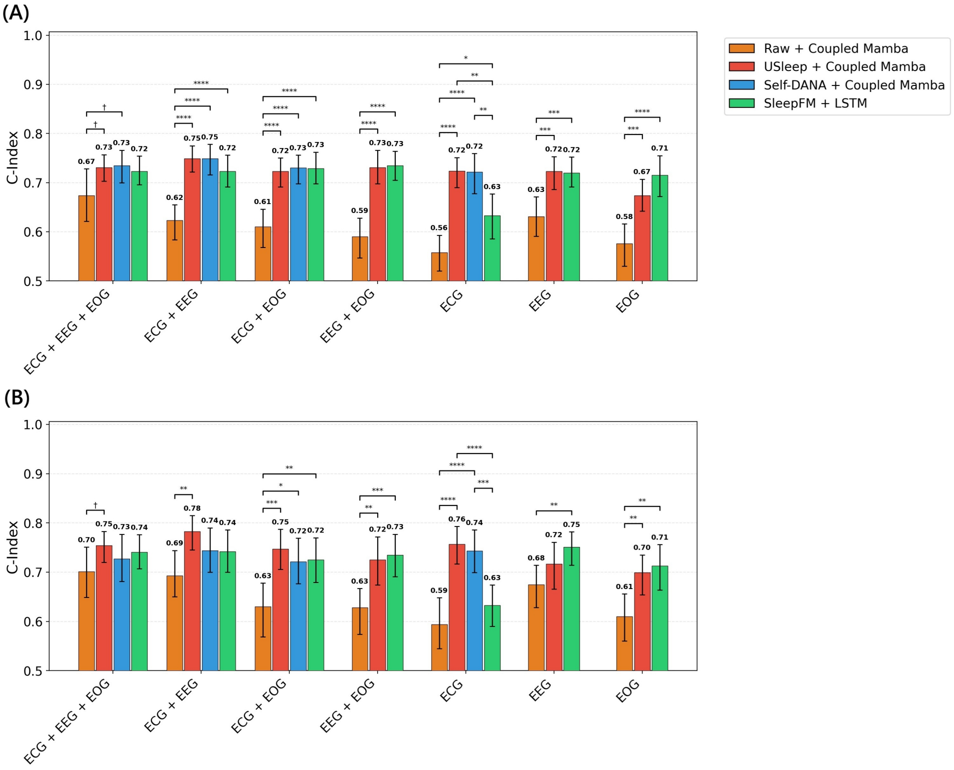


**Fig. S8**: C-index macro-average performance in SHHS2 stratified by SHHS1 training-set overlap across modality ablation configurations. Within-cohort temporal validation on SHHS2 for **(A)** the SHHS1-training-overlapping subset and **(B)** the SHHS1-training-disjoint subset, assessing the signal-specific impact (ECG, EEG, EOG) conducted exclusively in the “no-demographics" configuration. Error bars represent 95% confidence intervals derived from 100 bootstrap iterations. Statistical significance between model configurations: † $p \leq 0.1$; * $p \leq 0.05$; ** $p \leq 0.01$; *** $p \leq 0.001$; **** $p \leq 0.0001$

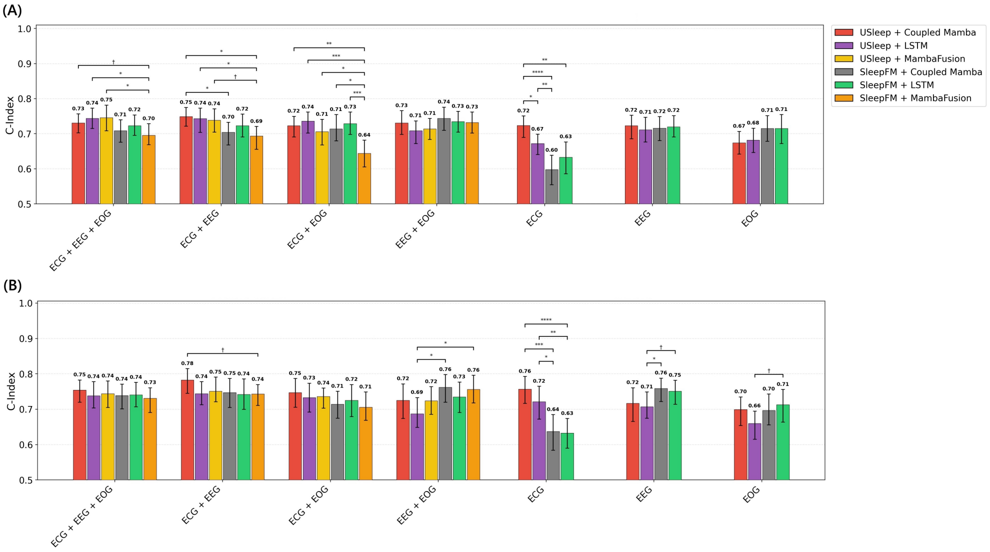


**Fig. S9**: C-index macro-average performance comparison between fusion engines (Coupled Mamba, LSTM and Mamba Fusion) in SHHS2 stratified by training-set overlap. Performance evaluation for **(A)** the SHHS1-training-overlapping subset and **(B)** the SHHS1-training-disjoint subset across different feature backbones (U-Sleep and SleepFM), highlighting changes in the fusion strategies. Error bars represent 95% confidence intervals derived from 100 bootstrap iterations. Statistical significance between model configurations: † $p \leq 0.1$; * $p \leq 0.05$; ** $p \leq 0.01$; *** $p \leq 0.001$; **** $p \leq 0.0001$

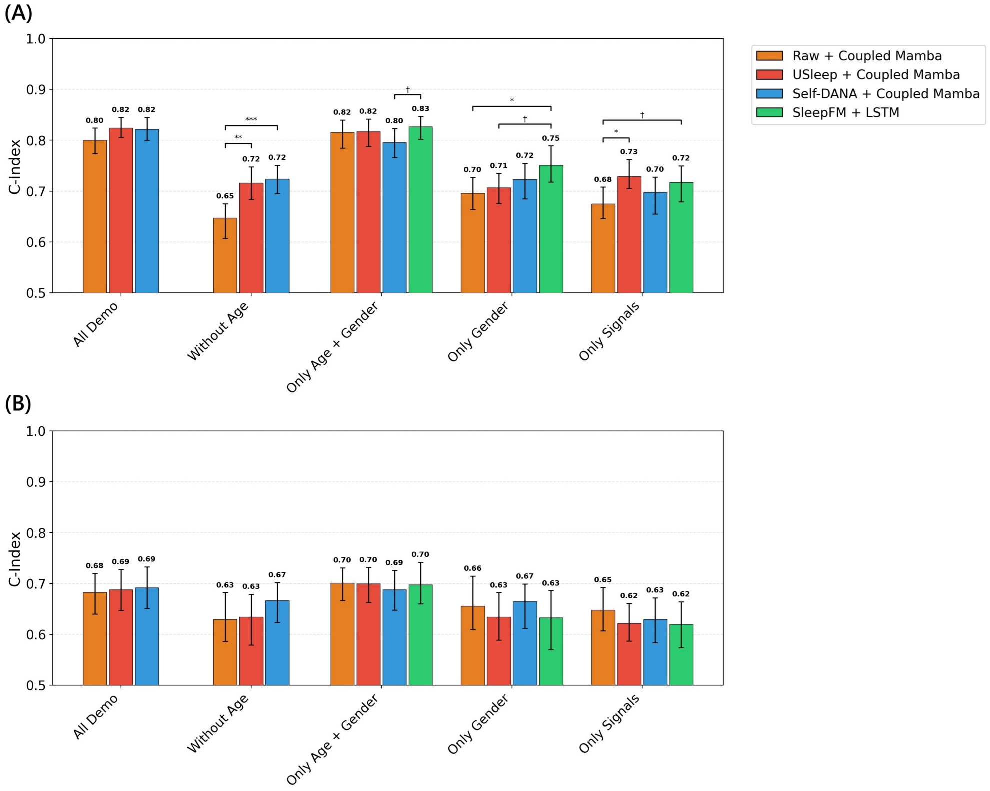


**Fig. S10**: C-index macro-average performance in SHHS1 for primary vs. secondary prevention across demographic configurations. Comparison of C-index results between the Coupled Mamba framework and SleepFM, stratified by participants in primary prevention**(A)** and those in secondary prevention **(B)**. Error bars represent 95% confidence intervals derived from 100 bootstrap iterations. Statistical significance between model configurations: † $p \leq 0.1$; * $p \leq 0.05$; ** $p \leq 0.01$; *** $p \leq 0.001$; **** $p \leq 0.0001$

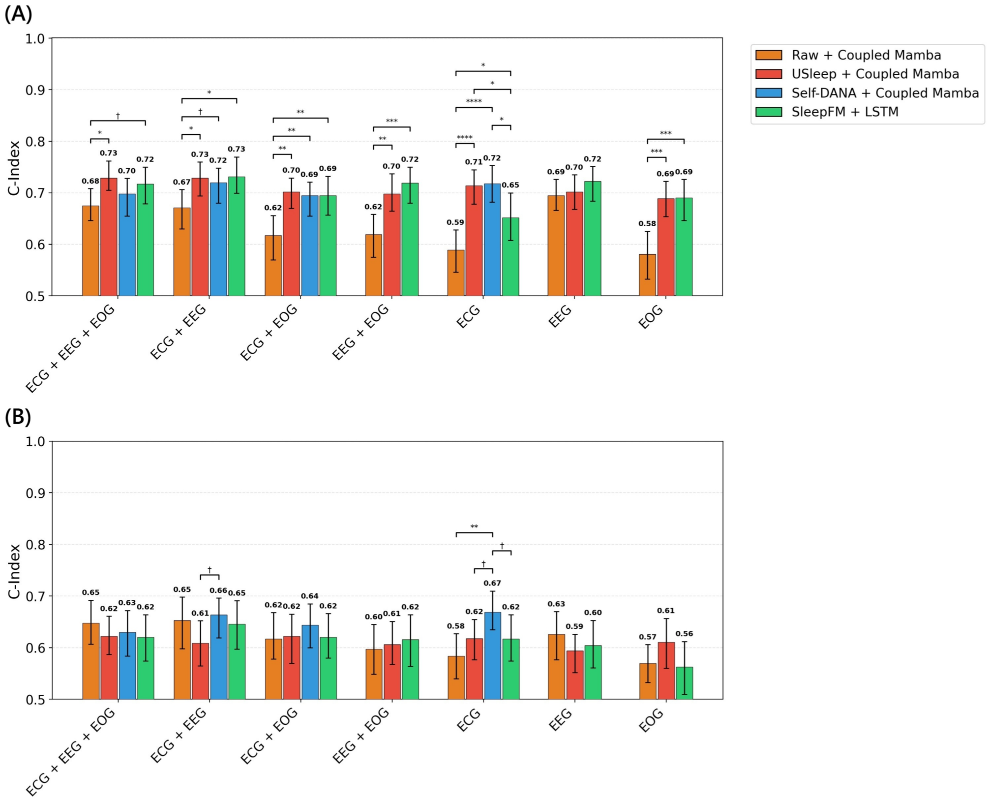


**Fig. S11**: C-index macro-average performance in SHHS1 for primary vs. secondary prevention across modality ablation configurations. Internal testing results for primary **(A)** and secondary **(B)** prevention, evaluating the signal-specific contributions (ECG, EEG, EOG) conducted exclusively in the "no-demographics" configuration. Error bars represent 95% confidence intervals derived from 100 bootstrap iterations. Statistical significance between model configurations: † p $\leq$ 0.1; * p $\leq$ 0.05; ** p $\leq$ 0.01; *** p $\leq$ 0.001; **** p $\leq$ 0.0001

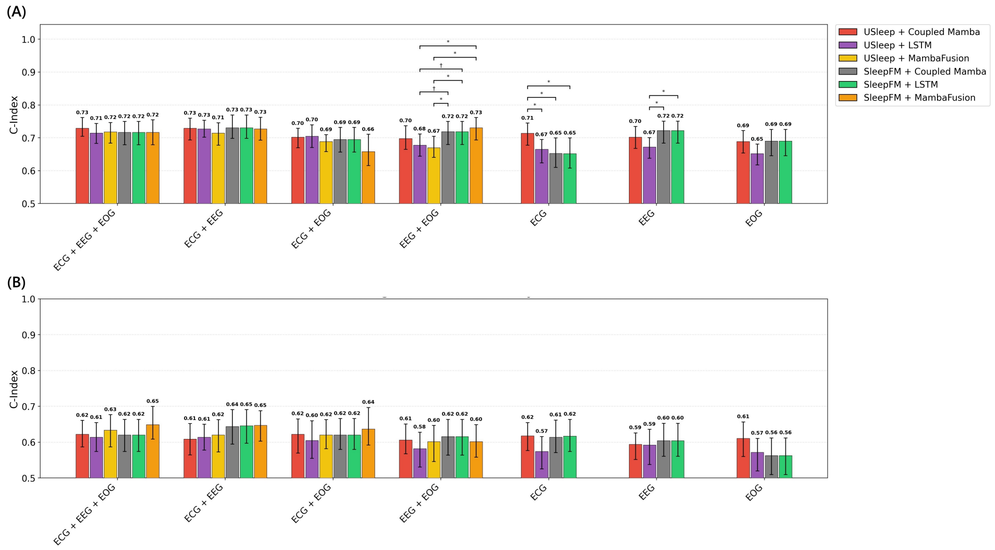


**Fig. S12**: C-index macro-average performance comparison between fusion engines (Coupled Mamba, LSTM and Mamba Fusion) in SHHS1 for primary and secondary prevention. Performance evaluation for primary **(A)** and secondary **(B)** prevention across different feature backbones (U-Sleep and SleepFM), changing the fusion strategy. Error bars represent 95% confidence intervals derived from 100 bootstrap iterations. Statistical significance between model configurations: † $p \leq 0.1$; * $p \leq 0.05$; ** $p \leq 0.01$; *** $p \leq 0.001$; **** $p \leq 0.0001$

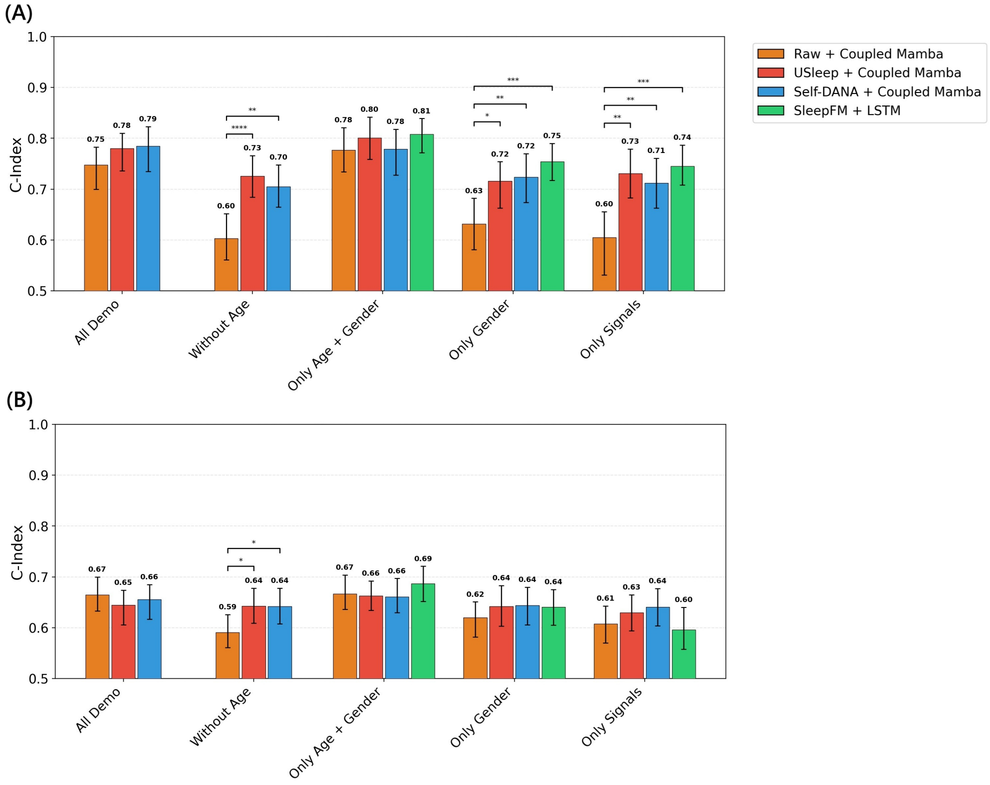


**Fig. S13**: C-index macro-average performance in SHHS2 for primary vs. secondary prevention across demographic configurations. Within-cohort temporal validation of stratified performance on the SHHS2 follow-up cohort for primary **(A)** and secondary **(B)** prevention. Error bars represent 95% confidence intervals derived from 100 bootstrap iterations. Statistical significance between model configurations: † $p \leq 0.1$; * $p \leq 0.05$; ** $p \leq 0.01$; *** $p \leq 0.001$; **** $p \leq 0.0001$

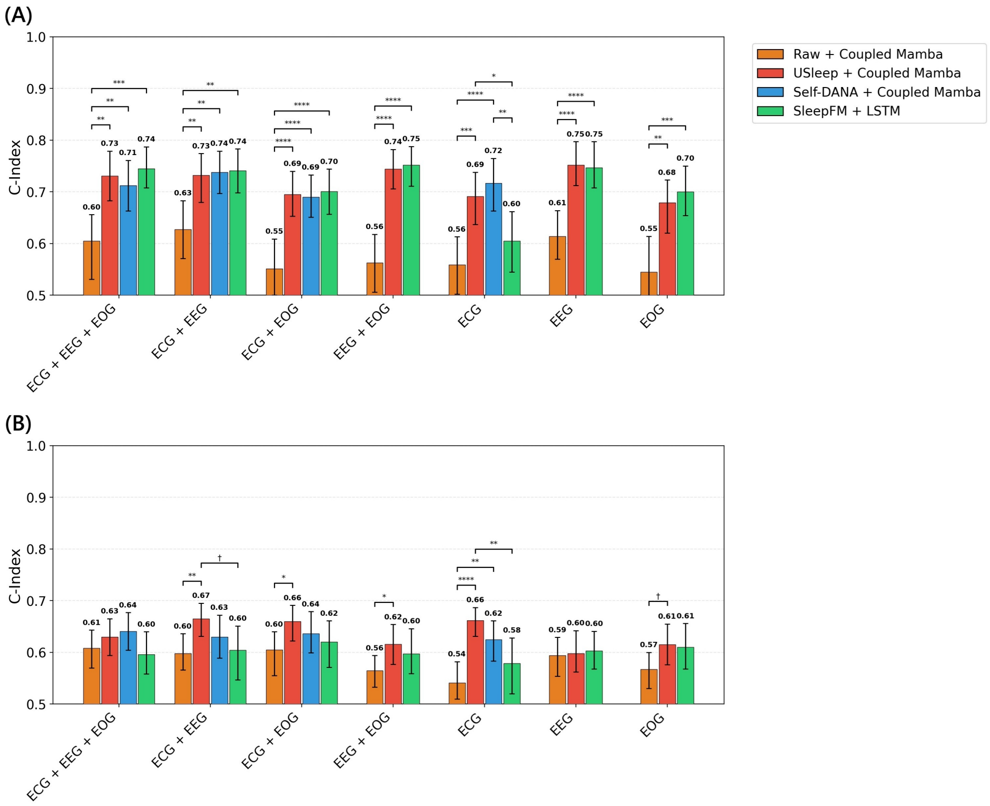


**Fig. S14**: C-index macro-average performance in SHHS2 for primary vs. secondary prevention across modality ablation configurations. Within-cohort temporal validation for primary **(A)** and secondary **(B)** prevention, assessing the longitudinal stability of modality-specific performance across different signal combinations in the "no-demographics configuration". Error bars represent 95% confidence intervals derived from 100 bootstrap iterations. Statistical significance between model configurations: † $p \leq 0.1$; * $p \leq 0.05$; ** $p \leq 0.01$; *** $p \leq 0.001$; **** $p \leq 0.0001$

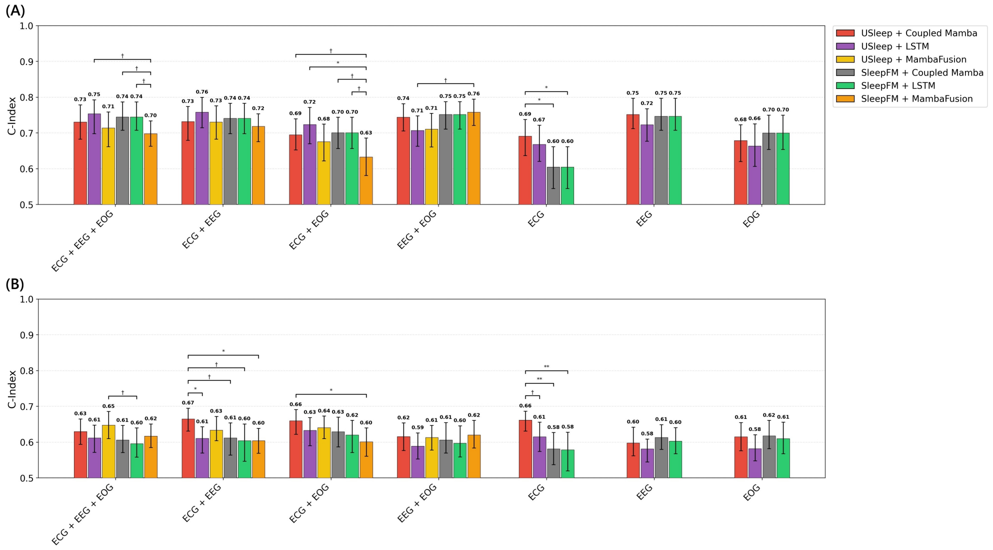


**Fig. S15**: C-index macro-average performance comparison between fusion engines (Coupled Mamba, LSTM and Mamba Fusion) in SHHS2 for primary and secondary prevention. Within-cohort temporal validation for primary **(A)** and secondary **(B)** prevention, highlighting the differences between fusion architectures across different feature backbones. Error bars represent 95% confidence intervals derived from 100 bootstrap iterations. Statistical significance between model configurations: † $p \leq 0.1$; * $p \leq 0.05$; ** $p \leq 0.01$; *** $p \leq 0.001$; **** $p \leq 0.0001$

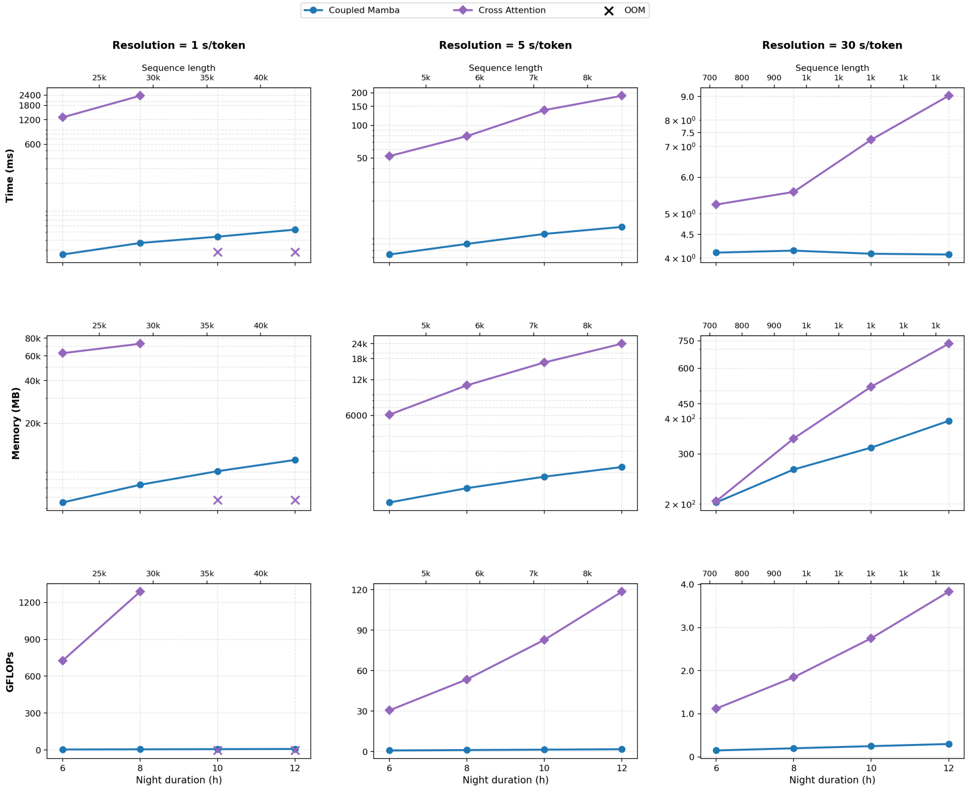


**Fig. S16**: Computational benchmarking of full-night cross-modal fusion architectures. Comparison of Coupled Mamba and Cross-Attention fusion on sequences of increasing duration (6–12 h) and token resolution (1, 5, 30 s). Metrics include inference runtime (ms), peak GPU memory (MB), and GFLOPs. Benchmarks were performed on a single NVIDIA A100 (80 GB) GPU using inference-only forward passes. “X” denotes out-of-memory failures.

### Supplementary tables

**Table S1**: SHHS2 baseline characteristics. Continuous variables are presented as median [interquartile range], while categorical variables are reported as percentages. Statistical significance was assessed using the Mann-Whitney test for continuous variables and the $\chi^2$-test for categorical data. The strength of associations (effect size) was quantified using Spearman's rank correlation for continuous variables and Cramer's V for categorical variables.

| | **N** | **Age (y)** | | **Gender-M (%)** | | **BMI ($kg/m^2$)** | | **CVD Meds (%)** | | **Sleep Meds (%)** | | **AHI (ev/h)** | | **Prev. CVD (%)** | |
|---|---|---|---|---|---|---|---|---|---|---|---|---|---|---|---|
| **Overall** | **2618** | **68 [60-76]** | | **46.3** | | **27.7 [24.9-31.1]** | | **61.7** | | **30.1** | | **13.5 [6.7-24.4]** | | **23.2** | |
| **Outcome** | | **Ev / No** | **Eff[a]** | **Ev / No** | **Eff[b]** | **Ev / No** | **Eff[a]** | **Ev / No** | **Eff[b]** | **Ev / No** | **Eff[b]** | **Ev / No** | **Eff[a]** | **Ev / No** | **Eff[b]** |
| Angina | 1636 | 80 [79-84] / 67 [56-78] | .26** | 54.5 / 45.1 | .04$^{ns}$ | 26.8 [23.9-29.3] / 27.4 [24.6-30.8] | .06 | 78.8 / 55.8 | .11** | 32.3 / 30.4 | .01$^{ns}$ | 18.7 [9.6-31.1] / 13.3 [6.5-23.7] | .09* | 59.6 / 25.5 | .18** |
| CVD Death | 2618 | 81 [77-84] / 67 [60-75] | .24** | 57.1 / 45.8 | .04 | 26.1 [23.9-29.2] / 27.8 [25-31.2] | .07** | 78.6 / 60.9 | .07* | 36.6 / 29.8 | .03$^{ns}$ | 21.1 [11.7-36.3] / 13.2 [6.7-23.7] | .10* | 69.6 / 21.1 | .23** |
| CHF | 2618 | 78 [73-83] / 67 [60-75] | .29** | 53.5 / 45.7 | .04 | 27.1 [24.6-30.2] / 27.8 [24.9-31.2] | .04$^{ns}$ | 82.2 / 59.8 | .12** | 40.8 / 29.1 | .07* | 18.4 [10.5-30.9] / 13.2 [6.6-23.6] | .10* | 69.5 / 19.1 | .32** |
| CHD Death | 2618 | 81 [78-85] / 67 [60-75] | .20** | 61.8 / 45.8 | .05* | 26.1 [24.3-29.3] / 27.7 [24.9-31.2] | .05* | 82.9 / 61 | .07** | 36.8 / 29.9 | .02$^{ns}$ | 22.3 [12.1-37.4] / 13.3 [6.7-23.9] | .09* | 69.7 / 21.8 | .19** |
| MI | 2618 | 78 [70-82] / 67 [60-75] | .16** | 53.7 / 45.9 | .03$^{ns}$ | 27.1 [24.1-30.2] / 27.7 [24.9-31.2] | .04 | 73.6 / 61.1 | .05** | 33.1 / 30 | .01$^{ns}$ | 16 [9.1-26.9] / 13.5 [6.7-24.2] | .04* | 46.3 / 22.1 | .12*** |
| Stroke | 2618 | 76 [69-81] / 67 [60-75] | .11** | 49.3 / 46.2 | .01$^{ns}$ | 28 [24.5-31] / 27.7 [24.9-31.1] | .00$^{ns}$ | 78.1 / 61.2 | .06* | 27.4 / 30.2 | .01$^{ns}$ | 15.9 [8.9-28.9] / 13.4 [6.7-24.2] | .03$^{ns}$ | 75.3 / 21.7 | .21*** |

**Ev / No**: Event / No Event; **Eff**: Effect size (absolute value); **BMI**: Body Mass Index; **Sleep Meds**: medications known or suspected to alter sleep [23]; **Hispanic**: Hispanic/Latino ethnicity; **AHI**: Apnea-Hypopnea Index (events/hour); **Prev. CVD**: cardiovascular disease at baseline.
$^{ns}$ $p > 0.05$; * $p < 0.05$; ** $p < 0.01$; *** $p < 0.001$.
[a] Spearman $\rho$; [b] Cramer's V.

**Table S2**: Baseline characteristics of the SHHS1 training set. Continuous variables are presented as median [interquartile range], while categorical variables are reported as percentages. Statistical significance was assessed using the Mann-Whitney test for continuous variables and the $\chi^2$-test for categorical data. The strength of associations (effect size) was quantified using Spearman's rank correlation for continuous variables and Cramer's V for categorical variables.

| | N | Age (y) | | Gender-M (%) | | BMI ($kg/m^2$) | | CVD Meds (%) | | Sleep Meds (%) | | Hispanic (%) | | AHI (ev/h) | | Prev. CVD (%) | |
|---|---|---|---|---|---|---|---|---|---|---|---|---|---|---|---|---|---|
| **Overall** | **3026** | **63 [55-72]** | | **48.1** | | **27.5 [24.7-30.8]** | | **49.6** | | **22.3** | | **4.6** | | **13.2 [6.7-23.9]** | | **12.1** | |
| **Outcome** | | **Ev / No** | **Eff[a]** | **Ev / No** | **Eff[b]** | **Ev / No** | **Eff[a]** | **Ev / No** | **Eff[b]** | **Ev / No** | **Eff[b]** | **Ev / No** | **Eff[b]** | **Ev / No** | **Eff[a]** | **Ev / No** | **Eff[b]** |
| Angina | 2047 | 76 [74-80] / 62 [51-74] | .31** | 47.9 / 47.6 | .00[ns] | 27.6 [25.2-30.6] / 27.2 [24.4-30.2] | .03[ns] | 63.7 / 47 | .10* | 28.9 / 22.4 | .04[ns] | 1.6 / 7.3 | .06* | 17 [8.9-27.5] / 12.4 [6.2-23.1] | .07** | 30 / 12.2 | .15** |
| CVD Death | 3025 | 76 [73-81] / 62 [55-71] | .28** | 53.3 / 47.7 | .03[ns] | 26.9 [24.3-29.9] / 27.5 [24.8-30.9] | .03[ns] | 71.4 / 48.2 | .11* | 29.1 / 21.8 | .04 | 0 / 4.9 | .05** | 17.3 [8.8-27.7] / 13 [6.6-23.7] | .06** | 35.7 / 10.6 | .18** |
| CHF | 3026 | 74 [68-79] / 62 [55-70] | .31** | 53.4 / 47.5 | .04 | 28 [25.2-31.2] / 27.5 [24.7-30.7] | .04* | 68.4 / 47.4 | .13** | 32.2 / 21.1 | .08* | 0.3 / 5.1 | .07* | 18 [9.9-29.2] / 12.8 [6.4-23.3] | .11* | 35.6 / 9.3 | .25** |
| CHD Death | 3025 | 76 [73-80.8] / 62 [55-71] | .22** | 59 / 47.6 | .04 | 27 [24.4-30.2] / 27.5 [24.8-30.9] | .02[ns] | 73 / 48.6 | .09** | 26.2 / 22.1 | .02[ns] | 0 / 4.8 | .04 | 18.4 [9.5-28.5] / 13 [6.6-23.7] | .06** | 38.5 / 11 | .16*** |
| MI | 3026 | 74 [66.2-78] / 62 [55-71] | .19** | 58.6 / 47.4 | .05* | 28.2 [25.4-31] / 27.5 [24.7-30.8] | .02[ns] | 58.6 / 49 | .04* | 25.3 / 22.1 | .02[ns] | 3.8 / 4.6 | .01[ns] | 16.9 [8.4-27.9] / 13 [6.5-23.7] | .05** | 23.7 / 11.4 | .09*** |
| Stroke | 3026 | 75 [69-79] / 62 [55-71] | .20** | 38.1 / 48.5 | .04 | 27.5 [24.6-30.5] / 27.5 [24.8-30.8] | .01[ns] | 64.9 / 48.9 | .06** | 33.6 / 21.7 | .06* | 0.7 / 4.8 | .04* | 15.3 [8.2-24.1] / 13.1 [6.6-23.9] | .03[ns] | 33.6 / 11.1 | .14*** |

**Ev / No**: Event / No Event; **Eff**: Effect size (absolute value); **BMI**: Body Mass Index; **Sleep Meds**: medications known or suspected to alter sleep [23]; **Hispanic**: Hispanic/Latino ethnicity; **AHI**: Apnea-Hypopnea Index (events/hour); **Prev. CVD**: cardiovascular disease at baseline.
[ns] $p > 0.05$; * $p < 0.05$; ** $p < 0.01$; *** $p < 0.001$.
[a] Spearman $\rho$; [b] Cramer's V.

**Table S3**: Baseline characteristics of the SHHS1 validation set. Continuous variables are presented as median [interquartile range], while categorical variables are reported as percentages. Statistical significance was assessed using the Mann-Whitney test for continuous variables and the $\chi^2$-test for categorical data. The strength of associations (effect size) was quantified using Spearman's rank correlation for continuous variables and Cramer's V for categorical variables.

| | N | Age (y) | | Gender-M (%) | | BMI ($kg/m^2$) | | CVD Meds (%) | | Sleep Meds (%) | | Hispanic (%) | | AHI (ev/h) | | Prev. CVD (%) | |
|---|---|---|---|---|---|---|---|---|---|---|---|---|---|---|---|---|---|
| **Overall** | **757** | **62 [55-72]** | | **46.4** | | **27.5 [24.7-30.8]** | | **49.1** | | **22.9** | | **5.2** | | **13.7 [6.9-24.8]** | | **12** | |
| **Outcome** | | **Ev / No** | **Eff$^a$** | **Ev / No** | **Eff$^b$** | **Ev / No** | **Eff$^a$** | **Ev / No** | **Eff$^b$** | **Ev / No** | **Eff$^b$** | **Ev / No** | **Eff$^b$** | **Ev / No** | **Eff$^a$** | **Ev / No** | **Eff$^b$** |
| Angina | 505 | 77 [74-81] / 60 [51-74] | .35** | 47.9 / 44.4 | .01$^{ns}$ | 27.3 [24.5-29.6] / 27.5 [24.5-30.7] | .04$^{ns}$ | 68.8 / 45.1 | .13* | 31.2 / 21.2 | .06$^{ns}$ | 0 / 8.5 | .08$^{ns}$ | 17.8 [9.2-24.3] / 13.5 [6.2-24.5] | .05$^{ns}$ | 47.9 / 9.4 | .33*** |
| CVD Death | 756 | 77 [72-82] / 61 [54-70] | .29** | 50 / 46.1 | .01$^{ns}$ | 27.3 [24.9-29.7] / 27.6 [24.7-30.9] | .04$^{ns}$ | 78.3 / 47.2 | .14* | 30.4 / 22.4 | .04$^{ns}$ | 0 / 5.5 | .05$^{ns}$ | 16.9 [9.4-26.7] / 13.6 [6.8-24.7] | .04$^{ns}$ | 54.3 / 9.2 | .33** |
| CHF | 757 | 76 [68.8-80] / 60 [54-70] | .34** | 55 / 45.3 | .06$^{ns}$ | 28.4 [25.7-32.1] / 27.5 [24.6-30.7] | .07 | 71.2 / 46.5 | .15** | 31.2 / 21.9 | .06$^{ns}$ | 0 / 5.8 | .07$^{ns}$ | 18.8 [10.8-31.4] / 13.1 [6.7-24.2] | .11* | 43.8 / 8.3 | .33*** |
| CHD Death | 756 | 77.5 [72.2-81.8] / 61 [54-70.8] | .24** | 56.7 / 45.9 | .04$^{ns}$ | 26.8 [25-29.6] / 27.6 [24.7-30.9] | .04$^{ns}$ | 70 / 48.2 | .08 | 30 / 22.6 | .03$^{ns}$ | 0 / 5.4 | .03$^{ns}$ | 12.5 [8.5-22.4] / 13.8 [6.9-25.2] | .01$^{ns}$ | 60 / 9.9 | .29*** |
| MI | 757 | 76 [69.5-80] / 61 [54-70] | .25** | 63.8 / 45.2 | .09 | 27.4 [25.1-29.6] / 27.5 [24.7-30.8] | .00$^{ns}$ | 63.8 / 48.2 | .07$^{ns}$ | 23.4 / 22.8 | .00$^{ns}$ | 2.1 / 5.4 | .02$^{ns}$ | 17.6 [10.7-23.2] / 13.4 [6.9-25.2] | .05$^{ns}$ | 40.4 / 10.1 | .22*** |
| Stroke | 757 | 74 [65.2-78.5] / 61 [54.5-71] | .18** | 50 / 46.2 | .01$^{ns}$ | 28.7 [25.3-30.2] / 27.5 [24.7-30.8] | .02$^{ns}$ | 70.6 / 48.1 | .09 | 26.5 / 22.7 | .01$^{ns}$ | 5.9 / 5.1 | .00$^{ns}$ | 20.8 [10-39.6] / 13.5 [6.9-24.4] | .09* | 41.2 / 10.7 | .18*** |

**Ev / No**: Event / No Event; **Eff**: Effect size (absolute value); **BMI**: Body Mass Index; **Sleep Meds**: medications known or suspected to alter sleep [23]; **Hispanic**: Hispanic/Latino ethnicity; **AHI**: Apnea-Hypopnea Index (events/hour). **Prev. CVD**: cardiovascular disease at baseline.

$^{ns}$ $p > 0.05$; * $p < 0.05$; ** $p < 0.01$; *** $p < 0.001$.

$^a$ Spearman $\rho$; $^b$ Cramer's V.

**Table S4**: Baseline characteristics of the SHHS1 test set. Continuous variables are presented as median [interquartile range], while categorical variables are reported as percentages. Statistical significance was assessed using the Mann-Whitney test for continuous variables and the $\chi^2$-test for categorical data. The strength of associations (effect size) was quantified using Spearman's rank correlation for continuous variables and Cramer's V for categorical variables.

| | N | Age (y) | | Gender-M (%) | | BMI ($kg/m^2$) | | CVD Meds (%) | | Sleep Meds (%) | | Hispanic (%) | | AHI (ev/h) | | Prev. CVD (%) | |
|---|---|---|---|---|---|---|---|---|---|---|---|---|---|---|---|---|---|
| **Overall** | **1991** | **64 [55-72]** | | **47.4** | | **27.4 [24.6-30.6]** | | **48.4** | | **21.9** | | **4.3** | | **13.1 [6.7-23.1]** | | **12.4** | |
| **Outcome** | | **Ev / No** | **Eff**$^a$ | **Ev / No** | **Eff**$^b$ | **Ev / No** | **Eff**$^a$ | **Ev / No** | **Eff**$^b$ | **Ev / No** | **Eff**$^b$ | **Ev / No** | **Eff**$^b$ | **Ev / No** | **Eff**$^a$ | **Ev / No** | **Eff**$^b$ |
| Angina | 1384 | 76 [74-80] / 63 [51-74] | .29** | 54.9 / 46.4 | .05$^{ns}$ | 27.7 [25-30.8] / 26.9 [24.4-30.2] | .04$^{ns}$ | 71.3 / 43.8 | .15* | 30.3 / 21.2 | .06 | 0 / 6.8 | .07** | 17.5 [11.3-28.8] / 12.6 [6.2-22.6] | .12** | 34.4 / 11.8 | .18** |
| CVD Death | 1990 | 76.5 [74-80] / 63 [54-71] | .30** | 57.9 / 46.7 | .05 | 27.3 [24-30.5] / 27.4 [24.7-30.6] | .01$^{ns}$ | 69.8 / 46.9 | .11** | 32.5 / 21.2 | .06* | 0 / 4.6 | .05* | 17 [10.7-29.8] / 12.7 [6.4-22.8] | .10** | 44.4 / 10.2 | .25** |
| CHF | 1991 | 75 [70-79] / 62 [54-70] | .34** | 54.4 / 46.6 | .05 | 28.3 [25.1-32.3] / 27.3 [24.6-30.4] | .06** | 70 / 45.8 | .15** | 32.7 / 20.6 | .09* | 0.5 / 4.8 | .06* | 16 [9.4-29.5] / 12.7 [6.4-22.6] | .09** | 39.6 / 9.1 | .29** |
| CHD Death | 1990 | 76 [72-80] / 63 [54-71] | .23** | 64.6 / 46.7 | .07* | 27.7 [25-30.6] / 27.4 [24.6-30.6] | .01$^{ns}$ | 74.7 / 47.3 | .10** | 29.1 / 21.6 | .03$^{ns}$ | 0 / 4.5 | .04$^{ns}$ | 16.7 [11.5-31.2] / 12.8 [6.5-22.9] | .09* | 45.6 / 11 | .20** |
| MI | 1991 | 74 [67-78] / 63 [54-71] | .19** | 65.6 / 46.2 | .09* | 27.5 [25-30.5] / 27.4 [24.6-30.6] | .00$^{ns}$ | 69.6 / 47 | .11* | 28 / 21.5 | .04$^{ns}$ | 1.6 / 4.5 | .03$^{ns}$ | 15.5 [9.8-28.8] / 12.8 [6.5-22.9] | .06* | 29.6 / 11.3 | .13*** |
| Stroke | 1991 | 74 [70-78] / 63 [54-71] | .23** | 44.1 / 47.6 | .01$^{ns}$ | 27.2 [23.7-30.8] / 27.4 [24.7-30.6] | .02$^{ns}$ | 63.6 / 47.5 | .07* | 23.7 / 21.8 | .01$^{ns}$ | 0.8 / 4.5 | .04$^{ns}$ | 16.1 [8-25.8] / 12.8 [6.5-23] | .05* | 38.1 / 10.8 | .19*** |

**Ev / No**: Event / No Event; **Eff**: Effect size (absolute value); **BMI**: Body Mass Index; **Sleep Meds**: medications known or suspected to alter sleep [23]; **Hispanic**: Hispanic/Latino ethnicity; **AHI**: Apnea-Hypopnea Index (events/hour); **Prev. CVD**: cardiovascular disease at baseline.
$^{ns}$ $p > 0.05$; * $p < 0.05$; ** $p < 0.01$; *** $p < 0.001$.
$^a$ Spearman $\rho$; $^b$ Cramer's V.

**Table S5**: Comparison with the only demographics model. Comparison of macro-average and outcome-specific C-index scores between SleepFM replica and demographic (age and gender only) MLP baseline on the SHHS1 test set.

| Outcome | SleepFM replica | MLP (age+gender) |
|---|---|---|
| Angina | 0.82 [0.79-0.84] | 0.81 [0.78-0.84] |
| CVD Death | 0.87 [0.84-0.89] | 0.87 [0.84-0.89] |
| CHF | 0.82 [0.8-0.84] | 0.82 [0.8-0.84] |
| CHD Death | 0.86 [0.82-0.89] | 0.87 [0.83-0.9] |
| MI | 0.77 [0.73-0.81] | 0.77 [0.73-0.8] |
| Stroke | 0.79 [0.75-0.84] | 0.79 [0.75-0.82] |
| Macro AVG | 0.82 (0.8-0.84) | 0.82 (0.81 - 0.84) |

**Table S6**: Multivariable Cox regression results across model configurations. Hazard Ratios (HR) and 95% Confidence Intervals (CI) are reported per 1 standard deviation increase in the PSG-derived risk score. $p$-values were adjusted using the Benjamini–Hochberg procedure across the six outcomes within each configuration: $^{*}p < 0.05$, $^{**}p < 0.01$, $^{***}p < 0.001$.

| Outcome | U-Sleep | Self-DANA | Raw |
|---|---|---|---|
| Angina | 0.85 (0.67–1.07) | 0.88 (0.71–1.08) | 1.17 (0.96–1.42) |
| CVD Death | 1.52 (1.21–1.90)$^{**}$ | 1.48 (1.23–1.77)$^{***}$ | 1.42 (1.23–1.64)$^{***}$ |
| CHF | 1.25 (1.05–1.49)$^{*}$ | 1.36 (1.18–1.57)$^{***}$ | 1.43 (1.27–1.63)$^{***}$ |
| CHD Death | 1.61 (1.22–2.13)$^{**}$ | 1.61 (1.26–2.05)$^{***}$ | 1.53 (1.28–1.83)$^{***}$ |
| MI | 1.24 (1.00–1.55) | 0.94 (0.79–1.13) | 0.95 (0.78–1.14) |
| Stroke | 1.15 (0.91–1.46) | 1.24 (1.03–1.50)$^{*}$ | 1.10 (0.92–1.32) |

**Table S7**: Pairwise C-index differences ($\Delta$ = Multilabel − Single-label) across SHHS1 evaluation subsets. Cells show $\Delta$C-index: positive values indicate superior performance of the multilabel architecture. Statistical significance: * $p \leq 0.1$.

| Outcome | All test | Primary Prev | Secondary Prev |
|---|---|---|---|
| Angina | -0.012 | -0.003 | -0.047 |
| CVD Death | +0.046* | +0.032 | +0.073 |
| CHF | -0.014 | -0.027 | +0.003 |
| CHD Death | +0.046 | +0.072 | +0.007 |
| MI | +0.048 | +0.061 | +0.025 |
| Stroke | -0.037 | -0.045 | -0.037 |
| Macro Avg | +0.013 | +0.015 | +0.004 |

**Table S8**: Pairwise C-index differences ($\Delta$ = Multilabel − Single-label) across SHHS2 evaluation subsets. Cells show $\Delta$C-index: positive values indicate superior performance of the multilabel architecture. Statistical significance: * $p \leq 0.1$.

| Outcome | All | Primary Prev | Secondary Prev | Training-overlap | Training-disjoint |
|---|---|---|---|---|---|
| Angina | -0.034 | -0.026 | -0.045 | -0.066 | +0.017 |
| CVD Death | +0.042 | +0.068 | +0.020 | +0.026 | +0.054 |
| CHF | -0.018 | -0.032 | -0.027 | -0.017 | -0.019 |
| CHD Death | +0.012 | +0.014 | +0.014 | +0.012 | +0.012 |
| MI | +0.020 | +0.024 | -0.019 | +0.011 | +0.034 |
| Stroke | -0.010 | -0.000 | -0.023 | -0.022 | -0.006 |
| Macro Avg | +0.002 | +0.008 | -0.013 | -0.009 | +0.017 |

**Table S9**: Pairwise C-index differences ($\Delta$ = Multilabel − No angina) across SHHS1 evaluation subsets. Cells show $\Delta$C-index: positive values indicate superior performance of the multilabel architecture. Statistical significance: * $p \leq 0.1$.

| Outcome | All test | Primary Prev | Secondary Prev |
|---|---|---|---|
| CVD Death | -0.001 | -0.010 | +0.004 |
| CHF | -0.004 | +0.005 | -0.017 |
| CHD Death | -0.026 | -0.017 | -0.038 |
| MI | +0.018 | +0.029 | -0.013 |
| Stroke | +0.011 | -0.011 | +0.010 |
| Macro Avg | -0.000 | -0.001 | -0.011 |

**Table S10**: Pairwise C-index differences ($\Delta$ = Multilabel − No angina) across SHHS2 evaluation subsets. Cells show $\Delta$C-index: positive values indicate superior performance of the multilabel architecture. Statistical significance: * $p \leq 0.1$.

| Outcome | All | Primary Prev | Secondary Prev | Training-overlap | Training-disjoint |
|---|---|---|---|---|---|
| CVD Death | -0.006 | +0.006 | -0.010 | -0.012 | +0.004 |
| CHF | +0.003 | -0.037 | +0.010 | +0.019 | -0.010 |
| CHD Death | -0.052* | -0.063 | -0.053 | -0.066* | -0.037 |
| MI | +0.007 | +0.012 | -0.006 | +0.007 | +0.021 |
| Stroke | +0.068* | +0.068 | +0.029 | +0.072 | +0.074 |
| Macro Avg | +0.004 | -0.003 | -0.006 | +0.004 | +0.010 |

**Table S11**: Outcome-specific C-index performance on the SHHS1 test set. Detailed breakdown of Harrell's C-index (with 95% confidence intervals) for each of the six cardiovascular labels: Angina, CVD Death, CHF, CHD Death, MI, and Stroke. The table summarizes performance across all experimental configurations, including variations in demographic features (with demographics) and physiological signal-only combinations (without demographics).

| Configuration | Model | Angina | CVD Death | CHF | CHD Death | MI | Stroke |
|---|---|---|---|---|---|---|---|
| **With demographics** | | | | | | | |
| All Demo | Raw | 0.792 (0.753-0.818) | 0.863 (0.833-0.889) | 0.820 (0.800-0.847) | 0.839 (0.808-0.878) | 0.740 (0.703-0.776) | 0.754 (0.702-0.796) |
| | USleep | 0.808 (0.784-0.839) | 0.866 (0.844-0.891) | 0.821 (0.791-0.844) | **0.865** (0.823-0.904) | **0.764** (0.719-0.803) | 0.784 (0.749-0.816) |
| | DANA | **0.810** (0.782-0.839) | **0.868** (0.839-0.894) | **0.825** (0.800-0.847) | 0.862 (0.817-0.902) | 0.756 (0.718-0.793) | **0.788** (0.751-0.821) |
| Without Age | Raw | 0.601 (0.552-0.649) | 0.748 (0.702-0.799) | 0.716 (0.682-0.754) | 0.684 (0.615-0.748) | 0.608 (0.559-0.650) | 0.666 (0.609-0.711) |
| | USleep | 0.701 (0.662-0.734) | 0.779 (0.739-0.819) | 0.741 (0.715-0.774) | 0.777 (0.735-0.826) | 0.658 (0.603-0.708) | **0.707** (0.665-0.746) |
| | DANA | **0.711** (0.668-0.750) | **0.798** (0.765-0.837) | **0.780** (0.746-0.809) | **0.813** (0.762-0.856) | **0.680** (0.637-0.722) | 0.686 (0.631-0.734) |
| Only Age and Gender | Raw | **0.813** (0.780-0.850) | 0.860 (0.832-0.880) | 0.831 (0.807-0.852) | 0.852 (0.822-0.881) | 0.756 (0.716-0.792) | 0.782 (0.743-0.821) |
| | USleep | 0.805 (0.769-0.839) | 0.864 (0.837-0.891) | 0.816 (0.792-0.837) | 0.865 (0.836-0.899) | 0.748 (0.710-0.786) | 0.787 (0.746-0.828) |
| | DANA | 0.783 (0.754-0.817) | 0.860 (0.832-0.888) | **0.834** (0.802-0.854) | 0.848 (0.805-0.879) | 0.697 (0.638-0.745) | 0.764 (0.724-0.807) |
| | SleepFM | 0.812 (0.782-0.842) | **0.872** (0.841-0.894) | 0.818 (0.797-0.843) | **0.868** (0.832-0.899) | **0.768** (0.731-0.802) | **0.791** (0.752-0.827) |
| Only Gender | Raw | 0.691 (0.647-0.733) | 0.771 (0.724-0.804) | 0.762 (0.732-0.798) | 0.779 (0.726-0.818) | 0.612 (0.569-0.666) | 0.661 (0.605-0.708) |
| | USleep | 0.686 (0.647-0.722) | 0.771 (0.732-0.811) | 0.739 (0.709-0.775) | 0.774 (0.723-0.825) | 0.646 (0.609-0.693) | 0.711 (0.673-0.755) |
| | DANA | 0.718 (0.679-0.752) | 0.789 (0.744-0.823) | 0.785 (0.758-0.808) | 0.796 (0.747-0.832) | 0.663 (0.610-0.715) | **0.712** (0.670-0.760) |
| | SleepFM | **0.724** (0.680-0.761) | **0.795** (0.753-0.828) | **0.790** (0.759-0.815) | **0.810** (0.764-0.852) | **0.704** (0.661-0.738) | 0.710 (0.657-0.754) |
| **Without demographics** | | | | | | | |
| Only Signals | Raw | 0.645 (0.594-0.687) | 0.753 (0.714-0.794) | 0.708 (0.680-0.739) | 0.787 (0.736-0.831) | 0.617 (0.566-0.676) | 0.656 (0.596-0.706) |
| | USleep | **0.685** (0.637-0.728) | **0.803** (0.765-0.834) | 0.758 (0.733-0.784) | **0.789** (0.742-0.850) | **0.676** (0.629-0.720) | 0.684 (0.644-0.725) |
| | DANA | 0.683 (0.640-0.724) | 0.775 (0.727-0.813) | **0.761** (0.736-0.789) | 0.770 (0.715-0.828) | 0.632 (0.587-0.686) | 0.676 (0.626-0.728) |
| | SleepFM | 0.677 (0.628-0.722) | 0.770 (0.732-0.805) | 0.721 (0.686-0.753) | **0.789** (0.743-0.847) | 0.637 (0.589-0.685) | **0.708** (0.661-0.745) |
| ECG+EEG | Raw | 0.641 (0.582-0.694) | 0.747 (0.696-0.784) | 0.726 (0.687-0.760) | 0.769 (0.721-0.821) | 0.603 (0.551-0.650) | 0.645 (0.596-0.693) |
| | USleep | 0.720 (0.676-0.768) | 0.772 (0.734-0.805) | 0.761 (0.733-0.786) | 0.771 (0.721-0.817) | 0.639 (0.598-0.688) | **0.740** (0.691-0.777) |
| | DANA | **0.727** (0.685-0.770) | **0.791** (0.753-0.825) | **0.765** (0.736-0.799) | 0.773 (0.723-0.825) | 0.675 (0.620-0.720) | 0.719 (0.683-0.750) |
| | SleepFM | 0.693 (0.640-0.737) | 0.782 (0.740-0.815) | 0.759 (0.724-0.789) | **0.800** (0.754-0.845) | **0.680** (0.633-0.723) | 0.698 (0.647-0.744) |
| ECG+EOG | Raw | 0.660 (0.609-0.705) | 0.675 (0.626-0.721) | 0.679 (0.643-0.713) | 0.670 (0.590-0.729) | 0.589 (0.538-0.634) | 0.619 (0.558-0.684) |
| | USleep | **0.715** (0.680-0.752) | 0.748 (0.710-0.789) | 0.750 (0.720-0.776) | 0.737 (0.668-0.801) | 0.630 (0.579-0.670) | 0.689 (0.640-0.725) |
| | DANA | 0.705 (0.665-0.748) | 0.745 (0.706-0.790) | **0.765** (0.739-0.792) | 0.748 (0.695-0.799) | 0.620 (0.564-0.682) | **0.691** (0.638-0.736) |
| | SleepFM | 0.678 (0.628-0.724) | **0.764** (0.726-0.795) | 0.715 (0.676-0.750) | **0.781** (0.721-0.822) | **0.641** (0.586-0.680) | 0.680 (0.637-0.723) |
| EEG+EOG | Raw | 0.615 (0.573-0.655) | 0.663 (0.618-0.715) | 0.640 (0.603-0.679) | 0.589 (0.542-0.651) | 0.604 (0.554-0.657) | 0.641 (0.599-0.697) |
| | USleep | **0.688** (0.654-0.730) | 0.747 (0.708-0.783) | **0.730** (0.702-0.764) | 0.768 (0.720-0.806) | 0.636 (0.593-0.675) | 0.688 (0.636-0.737) |
| | SleepFM | 0.674 (0.631-0.718) | **0.771** (0.732-0.801) | 0.728 (0.688-0.758) | **0.792** (0.750-0.836) | **0.647** (0.595-0.699) | **0.716** (0.668-0.754) |
| ECG | Raw | 0.494 (0.433-0.554) | 0.648 (0.595-0.704) | 0.694 (0.657-0.729) | 0.646 (0.581-0.717) | 0.593 (0.540-0.650) | 0.603 (0.547-0.641) |
| | USleep | **0.726** (0.686-0.767) | **0.788** (0.747-0.822) | 0.737 (0.706-0.769) | 0.773 (0.724-0.823) | **0.655** (0.607-0.698) | **0.708** (0.654-0.757) |
| | DANA | **0.726** (0.683-0.772) | **0.788** (0.750-0.825) | **0.786** (0.756-0.812) | **0.795** (0.740-0.847) | 0.649 (0.598-0.700) | 0.687 (0.643-0.733) |

**Table S11 – Continued from previous page**

| Configuration | Model | Angina | CVD Death | CHF | CHD Death | MI | Stroke |
|---|---|---|---|---|---|---|---|
| | SleepFM | 0.651 (0.594-0.700) | 0.703 (0.663-0.741) | 0.715 (0.689-0.746) | 0.721 (0.681-0.762) | 0.623 (0.572-0.676) | 0.625 (0.568-0.684) |
| EEG | Raw | 0.661 (0.613-0.717) | 0.737 (0.698-0.775) | 0.713 (0.681-0.748) | 0.778 (0.743-0.818) | 0.604 (0.554-0.651) | 0.674 (0.621-0.728) |
| | USleep | **0.685** (0.639-0.733) | 0.756 (0.720-0.788) | 0.715 (0.686-0.748) | 0.742 (0.698-0.796) | **0.661** (0.599-0.707) | 0.661 (0.623-0.704) |
| | SleepFM | 0.647 (0.598-0.688) | **0.778** (0.740-0.814) | **0.727** (0.693-0.758) | **0.786** (0.735-0.840) | 0.655 (0.611-0.691) | **0.689** (0.629-0.732) |
| EOG | Raw | 0.597 (0.546-0.648) | 0.624 (0.578-0.666) | 0.631 (0.589-0.668) | 0.606 (0.551-0.658) | 0.566 (0.510-0.629) | 0.566 (0.515-0.615) |
| | USleep | **0.710** (0.665-0.759) | **0.759** (0.725-0.797) | **0.706** (0.670-0.742) | **0.739** (0.687-0.799) | **0.617** (0.579-0.660) | 0.624 (0.583-0.665) |
| | SleepFM | 0.630 (0.579-0.676) | 0.737 (0.696-0.783) | 0.680 (0.646-0.712) | 0.721 (0.663-0.785) | 0.602 (0.552-0.652) | **0.676** (0.620-0.722) |

**Table S12**: Outcome-specific C-index performance on the SHHS2 follow-up cohort. Detailed breakdown of Harrell's C-index (with 95% confidence intervals) for each of the six cardiovascular labels: Angina, CVD Death, CHF, CHD Death, MI, and Stroke. The table summarizes performance across all experimental configurations, including variations in demographic features (with demographics) and physiological signal-only combinations (without demographics).

| Configuration | Model | Angina | CVD Death | CHF | CHD Death | MI | Stroke |
|---|---|---|---|---|---|---|---|
| **With demographics** | | | | | | | |
| All Demo | Raw | 0.735 (0.697-0.773) | 0.820 (0.782-0.855) | 0.785 (0.765-0.810) | 0.821 (0.784-0.851) | 0.702 (0.654-0.749) | **0.758** (0.702-0.819) |
| | USleep | 0.778 (0.741-0.812) | 0.838 (0.805-0.868) | 0.796 (0.770-0.817) | **0.850** (0.816-0.889) | 0.701 (0.644-0.747) | 0.707 (0.652-0.755) |
| | DANA | **0.799** (0.770-0.831) | **0.845** (0.815-0.872) | **0.807** (0.783-0.833) | 0.835 (0.799-0.872) | **0.727** (0.679-0.778) | 0.716 (0.669-0.774) |
| Without Age | Raw | 0.656 (0.601-0.706) | 0.679 (0.618-0.719) | 0.680 (0.656-0.709) | 0.620 (0.554-0.681) | 0.603 (0.559-0.654) | 0.654 (0.587-0.720) |
| | USleep | **0.738** (0.688-0.783) | **0.822** (0.786-0.859) | 0.771 (0.746-0.798) | **0.817** (0.778-0.859) | 0.652 (0.604-0.696) | 0.682 (0.629-0.731) |
| | DANA | 0.714 (0.673-0.764) | 0.799 (0.751-0.835) | **0.779** (0.747-0.806) | 0.799 (0.750-0.852) | **0.658** (0.614-0.706) | **0.695** (0.633-0.755) |
| Only Age and Gender | Raw | 0.788 (0.750-0.826) | 0.818 (0.789-0.846) | 0.800 (0.773-0.827) | 0.822 (0.785-0.851) | 0.721 (0.667-0.759) | 0.738 (0.672-0.798) |
| | USleep | **0.814** (0.781-0.848) | 0.843 (0.808-0.877) | 0.798 (0.771-0.820) | 0.856 (0.820-0.887) | 0.730 (0.686-0.772) | 0.722 (0.678-0.762) |
| | DANA | 0.796 (0.763-0.820) | 0.833 (0.804-0.863) | **0.830** (0.806-0.847) | 0.796 (0.750-0.834) | 0.715 (0.663-0.760) | 0.734 (0.674-0.787) |
| | SleepFM | 0.808 (0.764-0.838) | **0.853** (0.824-0.883) | 0.809 (0.783-0.835) | **0.862** (0.827-0.893) | **0.733** (0.691-0.771) | **0.760** (0.704-0.818) |
| Only Gender | Raw | 0.682 (0.637-0.726) | 0.698 (0.632-0.746) | 0.700 (0.666-0.736) | 0.712 (0.649-0.767) | 0.627 (0.588-0.675) | 0.666 (0.609-0.728) |
| | USleep | **0.732** (0.683-0.776) | **0.822** (0.793-0.851) | 0.771 (0.748-0.795) | 0.824 (0.788-0.855) | 0.659 (0.609-0.704) | 0.681 (0.615-0.730) |
| | DANA | 0.725 (0.688-0.761) | 0.801 (0.757-0.844) | **0.801** (0.775-0.824) | 0.788 (0.733-0.840) | 0.671 (0.635-0.716) | **0.710** (0.640-0.765) |
| | SleepFM | 0.724 (0.677-0.772) | 0.816 (0.788-0.851) | 0.790 (0.762-0.817) | **0.829** (0.787-0.875) | **0.681** (0.631-0.719) | 0.697 (0.638-0.743) |
| **Without demographics** | | | | | | | |
| Only Signals | Raw | 0.601 (0.553-0.648) | 0.697 (0.654-0.737) | 0.661 (0.621-0.697) | 0.725 (0.664-0.781) | 0.574 (0.524-0.622) | 0.655 (0.587-0.730) |
| | USleep | **0.713** (0.666-0.766) | **0.823** (0.793-0.845) | 0.764 (0.735-0.792) | **0.808** (0.769-0.841) | **0.678** (0.632-0.722) | 0.652 (0.607-0.710) |
| | DANA | 0.708 (0.648-0.768) | 0.809 (0.777-0.843) | **0.783** (0.754-0.811) | 0.750 (0.700-0.806) | 0.643 (0.591-0.682) | 0.674 (0.599-0.727) |
| | SleepFM | 0.684 (0.633-0.723) | 0.787 (0.755-0.822) | 0.746 (0.712-0.777) | 0.802 (0.767-0.842) | 0.648 (0.604-0.694) | **0.695** (0.627-0.757) |
| ECG+EEG | Raw | 0.652 (0.601-0.699) | 0.666 (0.619-0.709) | 0.650 (0.606-0.699) | 0.690 (0.637-0.738) | 0.581 (0.526-0.634) | **0.717** (0.638-0.783) |
| | USleep | **0.743** (0.708-0.793) | **0.818** (0.786-0.846) | 0.772 (0.747-0.797) | **0.841** (0.802-0.876) | **0.707** (0.666-0.747) | 0.712 (0.654-0.761) |
| | DANA | 0.701 (0.653-0.752) | 0.812 (0.775-0.849) | **0.803** (0.773-0.831) | 0.795 (0.734-0.837) | 0.649 (0.607-0.684) | 0.689 (0.627-0.745) |

**Table S12 – Continued from previous page**

| Configuration | Model | Angina | CVD Death | CHF | CHD Death | MI | Stroke |
|---|---|---|---|---|---|---|---|
| | SleepFM | 0.660 (0.600-0.713) | 0.790 (0.755-0.820) | 0.764 (0.726-0.798) | 0.805 (0.762-0.843) | 0.655 (0.600-0.705) | 0.696 (0.624-0.761) |
| ECG+EOG | Raw | 0.601 (0.526-0.662) | 0.648 (0.595-0.704) | 0.640 (0.603-0.683) | 0.661 (0.596-0.730) | 0.556 (0.505-0.601) | 0.627 (0.559-0.687) |
| | USleep | **0.760** (0.717-0.800) | **0.786** (0.748-0.827) | **0.767** (0.738-0.792) | **0.756** (0.686-0.808) | **0.680** (0.637-0.734) | 0.669 (0.621-0.725) |
| | DANA | 0.722 (0.672-0.762) | 0.769 (0.731-0.809) | 0.775 (0.746-0.802) | 0.739 (0.677-0.796) | 0.641 (0.589-0.681) | **0.707** (0.651-0.766) |
| | SleepFM | 0.678 (0.633-0.721) | 0.774 (0.733-0.818) | 0.749 (0.709-0.781) | 0.800 (0.753-0.842) | 0.630 (0.585-0.679) | 0.670 (0.592-0.731) |
| EEG+EOG | Raw | 0.626 (0.572-0.672) | 0.611 (0.554-0.666) | 0.602 (0.557-0.637) | 0.592 (0.532-0.651) | 0.558 (0.504-0.607) | 0.630 (0.566-0.693) |
| | USleep | **0.712** (0.671-0.751) | 0.786 (0.753-0.821) | 0.746 (0.716-0.773) | **0.805** (0.753-0.842) | 0.632 (0.585-0.688) | 0.683 (0.625-0.732) |
| | SleepFM | 0.688 (0.633-0.729) | **0.797** (0.762-0.831) | **0.748** (0.712-0.778) | 0.801 (0.761-0.847) | **0.656** (0.614-0.705) | **0.705** (0.638-0.771) |
| ECG | Raw | 0.466 (0.392-0.540) | 0.606 (0.550-0.668) | 0.619 (0.575-0.658) | 0.589 (0.521-0.649) | 0.577 (0.534-0.623) | 0.564 (0.490-0.630) |
| | USleep | **0.751** (0.696-0.793) | 0.787 (0.748-0.826) | 0.750 (0.721-0.780) | **0.793** (0.747-0.848) | **0.680** (0.626-0.730) | 0.670 (0.622-0.720) |
| | DANA | 0.676 (0.632-0.720) | **0.801** (0.763-0.834) | **0.758** (0.720-0.790) | 0.781 (0.731-0.827) | 0.629 (0.574-0.674) | **0.711** (0.648-0.784) |
| | SleepFM | 0.622 (0.561-0.675) | 0.671 (0.616-0.723) | 0.654 (0.623-0.694) | 0.675 (0.615-0.738) | 0.615 (0.561-0.675) | 0.558 (0.469-0.626) |
| EEG | Raw | 0.673 (0.614-0.730) | 0.657 (0.595-0.707) | 0.619 (0.582-0.661) | 0.673 (0.610-0.736) | 0.591 (0.540-0.647) | 0.663 (0.583-0.714) |
| | USleep | **0.686** (0.647-0.739) | **0.813** (0.779-0.845) | 0.737 (0.708-0.767) | **0.818** (0.769-0.857) | 0.594 (0.551-0.643) | 0.664 (0.609-0.725) |
| | SleepFM | 0.683 (0.613-0.735) | 0.796 (0.766-0.831) | **0.747** (0.713-0.774) | 0.805 (0.767-0.837) | **0.648** (0.595-0.700) | **0.699** (0.638-0.757) |
| EOG | Raw | 0.608 (0.558-0.665) | 0.599 (0.537-0.646) | 0.603 (0.562-0.637) | 0.587 (0.507-0.654) | 0.543 (0.485-0.586) | 0.618 (0.536-0.688) |
| | USleep | 0.711 (0.666-0.757) | **0.762** (0.715-0.803) | 0.701 (0.671-0.742) | 0.750 (0.704-0.800) | 0.619 (0.572-0.659) | 0.575 (0.507-0.631) |
| | SleepFM | **0.727** (0.673-0.772) | 0.742 (0.706-0.786) | **0.716** (0.677-0.755) | **0.755** (0.709-0.805) | **0.647** (0.602-0.692) | **0.655** (0.577-0.720) |

**Table S13**: Outcome-specific AUROC performance on the SHHS1 test cohort. Detailed breakdown of the area under the ROC curve (with 95% confidence intervals) at a 5-year horizon for each of the six cardiovascular labels: Angina, CVD Death, CHF, CHD Death, MI, and Stroke. The table summarizes performance across all experimental configurations, including variations in demographic features (with demographics) and physiological signal-only combinations (without demographics).

| Configuration | Model | Angina | CVD Death | CHF | CHD Death | MI | Stroke |
|---|---|---|---|---|---|---|---|
| **With demographics** | | | | | | | |
| All Demo | Raw | 0.815 (0.776-0.850) | 0.850 (0.798-0.892) | **0.833** (0.803-0.871) | 0.857 (0.789-0.913) | 0.767 (0.715-0.810) | 0.759 (0.715-0.812) |
| | U-Sleep | **0.834** (0.804-0.861) | 0.869 (0.837-0.900) | 0.831 (0.802-0.866) | 0.879 (0.828-0.927) | **0.784** (0.736-0.836) | 0.787 (0.737-0.839) |
| | DANA | 0.833 (0.801-0.859) | **0.875** (0.842-0.909) | 0.828 (0.793-0.865) | **0.883** (0.824-0.936) | 0.772 (0.711-0.821) | **0.788** (0.741-0.843) |
| Without Age | Raw | 0.643 (0.588-0.705) | 0.735 (0.662-0.810) | 0.738 (0.692-0.802) | 0.746 (0.635-0.862) | 0.662 (0.598-0.740) | 0.669 (0.604-0.732) |
| | U-Sleep | 0.741 (0.697-0.789) | 0.764 (0.712-0.831) | 0.794 (0.757-0.829) | 0.755 (0.654-0.861) | 0.694 (0.638-0.757) | 0.715 (0.658-0.777) |
| | DANA | **0.757** (0.711-0.810) | **0.858** (0.807-0.898) | **0.818** (0.778-0.863) | **0.878** (0.802-0.937) | **0.703** (0.628-0.781) | **0.717** (0.669-0.777) |
| Only Age and Gender | Raw | **0.838** (0.803-0.868) | 0.853 (0.816-0.888) | 0.843 (0.814-0.876) | 0.870 (0.818-0.920) | 0.784 (0.731-0.833) | 0.782 (0.738-0.838) |
| | U-Sleep | 0.834 (0.800-0.866) | 0.862 (0.827-0.897) | 0.831 (0.800-0.863) | 0.867 (0.821-0.916) | 0.761 (0.701-0.815) | 0.785 (0.739-0.840) |
| | DANA | 0.808 (0.771-0.840) | **0.891** (0.857-0.919) | **0.859** (0.825-0.894) | **0.876** (0.815-0.932) | 0.708 (0.622-0.783) | 0.772 (0.728-0.819) |
| | SleepFM | 0.836 (0.793-0.865) | 0.858 (0.821-0.899) | 0.822 (0.794-0.857) | 0.860 (0.816-0.906) | **0.794** (0.741-0.837) | **0.788** (0.742-0.843) |

**Table S13 – Continued from previous page**

| Configuration | Model | Angina | CVD Death | CHF | CHD Death | MI | Stroke |
|---|---|---|---|---|---|---|---|
| Only Gender | Raw | 0.743 (0.685-0.790) | 0.753 (0.677-0.824) | 0.785 (0.747-0.830) | 0.786 (0.680-0.886) | 0.613 (0.535-0.682) | 0.669 (0.605-0.731) |
| | U-Sleep | 0.732 (0.689-0.781) | 0.750 (0.692-0.822) | 0.789 (0.750-0.825) | 0.757 (0.654-0.862) | 0.686 (0.627-0.751) | 0.716 (0.658-0.776) |
| | DANA | **0.763** (0.720-0.808) | **0.839** (0.789-0.893) | 0.821 (0.780-0.862) | **0.854** (0.766-0.928) | 0.701 (0.622-0.772) | **0.736** (0.679-0.794) |
| | SleepFM | 0.751 (0.697-0.799) | 0.820 (0.771-0.863) | **0.825** (0.785-0.855) | 0.819 (0.719-0.890) | **0.731** (0.671-0.791) | 0.733 (0.661-0.798) |
| **Without demographics** | | | | | | | |
| Only Signals | Raw | 0.665 (0.606-0.728) | 0.732 (0.636-0.802) | 0.725 (0.673-0.780) | 0.737 (0.621-0.837) | 0.592 (0.525-0.673) | 0.675 (0.605-0.738) |
| | U-Sleep | 0.704 (0.645-0.747) | 0.817 (0.742-0.878) | 0.799 (0.753-0.841) | 0.803 (0.713-0.882) | **0.714** (0.657-0.777) | 0.694 (0.638-0.740) |
| | DANA | **0.719** (0.673-0.766) | **0.820** (0.768-0.872) | **0.811** (0.766-0.846) | **0.816** (0.738-0.894) | 0.653 (0.558-0.732) | 0.702 (0.620-0.772) |
| | SleepFM | 0.700 (0.644-0.753) | 0.781 (0.736-0.841) | 0.755 (0.706-0.792) | 0.807 (0.737-0.885) | 0.649 (0.566-0.714) | **0.722** (0.660-0.779) |
| ECG+EEG | Raw | 0.689 (0.633-0.748) | 0.741 (0.671-0.817) | 0.756 (0.708-0.813) | 0.779 (0.662-0.882) | 0.635 (0.565-0.731) | 0.652 (0.578-0.717) |
| | U-Sleep | 0.756 (0.710-0.806) | 0.769 (0.712-0.850) | 0.797 (0.761-0.835) | 0.758 (0.681-0.872) | 0.678 (0.599-0.745) | **0.757** (0.709-0.817) |
| | DANA | **0.765** (0.722-0.813) | **0.823** (0.775-0.868) | **0.825** (0.779-0.861) | 0.801 (0.699-0.880) | **0.733** (0.667-0.790) | 0.714 (0.640-0.781) |
| | SleepFM | 0.730 (0.673-0.773) | 0.810 (0.766-0.867) | 0.813 (0.779-0.854) | **0.837** (0.756-0.906) | 0.692 (0.619-0.762) | 0.737 (0.676-0.791) |
| ECG+EOG | Raw | 0.694 (0.635-0.754) | 0.696 (0.635-0.779) | 0.706 (0.657-0.749) | 0.743 (0.638-0.857) | 0.590 (0.531-0.673) | 0.635 (0.579-0.709) |
| | U-Sleep | **0.763** (0.722-0.800) | 0.759 (0.691-0.839) | 0.796 (0.747-0.836) | 0.737 (0.616-0.837) | 0.666 (0.589-0.737) | 0.683 (0.630-0.745) |
| | DANA | 0.755 (0.710-0.799) | **0.812** (0.763-0.862) | **0.815** (0.777-0.855) | **0.833** (0.754-0.894) | 0.662 (0.577-0.736) | **0.713** (0.663-0.769) |
| | SleepFM | 0.718 (0.656-0.767) | 0.789 (0.732-0.857) | 0.755 (0.707-0.802) | 0.801 (0.717-0.890) | **0.678** (0.603-0.754) | 0.699 (0.640-0.768) |
| EEG+EOG | Raw | 0.641 (0.577-0.691) | 0.659 (0.577-0.725) | 0.672 (0.625-0.718) | 0.649 (0.542-0.754) | 0.644 (0.570-0.730) | 0.642 (0.574-0.716) |
| | U-Sleep | **0.712** (0.653-0.755) | 0.776 (0.676-0.853) | **0.764** (0.709-0.812) | 0.791 (0.681-0.875) | 0.653 (0.571-0.713) | 0.730 (0.682-0.785) |
| | SleepFM | 0.706 (0.655-0.764) | **0.780** (0.714-0.842) | 0.763 (0.713-0.804) | **0.800** (0.725-0.884) | **0.663** (0.583-0.744) | **0.736** (0.671-0.792) |
| ECG | Raw | 0.468 (0.402-0.543) | 0.666 (0.584-0.747) | 0.715 (0.663-0.778) | 0.681 (0.561-0.817) | 0.580 (0.503-0.649) | 0.634 (0.574-0.703) |
| | U-Sleep | 0.765 (0.723-0.808) | 0.780 (0.718-0.844) | 0.783 (0.737-0.820) | 0.763 (0.648-0.875) | **0.678** (0.610-0.742) | **0.698** (0.645-0.748) |
| | DANA | **0.773** (0.723-0.817) | **0.822** (0.776-0.869) | **0.842** (0.814-0.876) | **0.826** (0.748-0.903) | 0.658 (0.564-0.742) | 0.681 (0.638-0.748) |
| | SleepFM | 0.688 (0.608-0.751) | 0.711 (0.641-0.788) | 0.760 (0.713-0.815) | 0.718 (0.578-0.823) | 0.634 (0.556-0.711) | 0.649 (0.570-0.728) |
| EEG | Raw | 0.702 (0.647-0.749) | 0.715 (0.643-0.778) | 0.741 (0.696-0.794) | **0.821** (0.741-0.888) | 0.628 (0.555-0.698) | 0.671 (0.602-0.731) |
| | U-Sleep | **0.706** (0.650-0.752) | **0.798** (0.727-0.861) | 0.751 (0.697-0.804) | 0.781 (0.695-0.875) | **0.718** (0.644-0.778) | 0.692 (0.636-0.763) |
| | SleepFM | 0.672 (0.613-0.721) | 0.776 (0.719-0.845) | **0.764** (0.715-0.799) | 0.784 (0.701-0.877) | 0.656 (0.589-0.722) | **0.697** (0.625-0.758) |
| EOG | Raw | 0.647 (0.596-0.699) | 0.666 (0.589-0.726) | 0.654 (0.593-0.710) | 0.688 (0.606-0.771) | 0.580 (0.534-0.653) | 0.593 (0.521-0.644) |
| | U-Sleep | **0.753** (0.703-0.795) | **0.800** (0.748-0.858) | **0.717** (0.669-0.771) | **0.803** (0.732-0.863) | **0.647** (0.563-0.709) | 0.603 (0.515-0.663) |
| | SleepFM | 0.666 (0.617-0.714) | 0.773 (0.714-0.848) | 0.703 (0.645-0.743) | 0.729 (0.600-0.847) | 0.632 (0.534-0.704) | **0.700** (0.637-0.772) |

**Table S14**: Outcome-specific AUROC performance on the SHHS2 follow-up cohort. Detailed breakdown of the area under the ROC curve (with 95% confidence intervals) at a 5-year horizon for each of the six cardiovascular labels: Angina, CVD Death, CHF, CHD Death, MI, and Stroke. The table summarizes performance across all experimental configurations, including variations in demographic features (with demographics) and physiological signal-only combinations (without demographics).

| Configuration | Model | Angina | CVD Death | CHF | CHD Death | MI | Stroke |
|---|---|---|---|---|---|---|---|
| **With demographics** | | | | | | | |
| All Demo | Raw | 0.799 (0.740-0.852) | 0.849 (0.807-0.892) | 0.796 (0.758-0.829) | **0.864** (0.823-0.903) | 0.728 (0.655-0.780) | 0.711 (0.630-0.783) |
| | U-Sleep | 0.803 (0.760-0.839) | 0.848 (0.806-0.880) | 0.817 (0.787-0.850) | 0.854 (0.798-0.898) | 0.719 (0.673-0.776) | 0.720 (0.659-0.775) |
| | DANA | **0.823** (0.784-0.854) | **0.851** (0.813-0.881) | **0.825** (0.794-0.853) | 0.835 (0.791-0.871) | **0.747** (0.697-0.805) | **0.736** (0.673-0.794) |
| Without Age | Raw | 0.665 (0.589-0.736) | 0.709 (0.651-0.774) | 0.696 (0.644-0.745) | 0.620 (0.547-0.699) | 0.615 (0.547-0.691) | 0.652 (0.572-0.732) |
| | U-Sleep | **0.770** (0.720-0.817) | **0.855** (0.817-0.883) | 0.786 (0.748-0.815) | **0.853** (0.804-0.889) | **0.667** (0.610-0.723) | 0.684 (0.622-0.744) |
| | DANA | 0.741 (0.679-0.791) | 0.810 (0.756-0.852) | **0.787** (0.752-0.815) | 0.797 (0.725-0.853) | 0.656 (0.603-0.714) | **0.721** (0.636-0.788) |
| Only Age and Gender | Raw | 0.820 (0.786-0.851) | 0.849 (0.816-0.880) | 0.812 (0.782-0.846) | 0.839 (0.806-0.870) | 0.751 (0.698-0.811) | 0.751 (0.693-0.817) |
| | U-Sleep | 0.845 (0.810-0.877) | 0.856 (0.816-0.885) | 0.813 (0.783-0.840) | 0.864 (0.819-0.897) | 0.748 (0.694-0.804) | 0.733 (0.679-0.788) |
| | DANA | 0.820 (0.783-0.856) | 0.831 (0.785-0.872) | **0.842** (0.811-0.868) | 0.787 (0.722-0.842) | 0.725 (0.670-0.783) | **0.756** (0.701-0.818) |
| | SleepFM | **0.853** (0.824-0.879) | **0.873** (0.834-0.900) | 0.819 (0.791-0.846) | **0.874** (0.834-0.906) | **0.758** (0.705-0.807) | 0.730 (0.672-0.793) |
| Only Gender | Raw | 0.700 (0.647-0.748) | 0.711 (0.657-0.758) | 0.707 (0.648-0.749) | 0.710 (0.637-0.769) | 0.649 (0.600-0.701) | 0.684 (0.632-0.748) |
| | U-Sleep | **0.764** (0.713-0.811) | **0.848** (0.809-0.878) | 0.782 (0.743-0.812) | **0.854** (0.806-0.891) | 0.670 (0.611-0.724) | 0.685 (0.624-0.746) |
| | DANA | 0.736 (0.680-0.784) | 0.804 (0.751-0.848) | 0.805 (0.769-0.830) | 0.784 (0.712-0.843) | 0.662 (0.608-0.716) | **0.731** (0.651-0.795) |
| | SleepFM | 0.752 (0.709-0.801) | 0.824 (0.785-0.868) | **0.806** (0.775-0.838) | 0.832 (0.781-0.884) | **0.704** (0.651-0.753) | 0.701 (0.634-0.763) |
| **Without demographics** | | | | | | | |
| Only Signals | Raw | 0.658 (0.549-0.735) | 0.723 (0.628-0.814) | 0.720 (0.669-0.781) | 0.739 (0.662-0.835) | 0.606 (0.528-0.677) | **0.731** (0.635-0.822) |
| | U-Sleep | 0.743 (0.685-0.794) | **0.842** (0.801-0.884) | 0.775 (0.733-0.803) | **0.822** (0.783-0.865) | 0.679 (0.628-0.723) | 0.653 (0.601-0.701) |
| | DANA | **0.746** (0.686-0.803) | 0.805 (0.742-0.851) | **0.792** (0.757-0.821) | 0.728 (0.654-0.788) | 0.660 (0.606-0.712) | 0.687 (0.615-0.763) |
| | SleepFM | 0.728 (0.677-0.768) | 0.806 (0.763-0.843) | 0.774 (0.739-0.809) | 0.821 (0.778-0.867) | **0.688** (0.627-0.735) | 0.698 (0.633-0.757) |
| ECG+EEG | Raw | 0.687 (0.632-0.754) | 0.680 (0.627-0.731) | 0.681 (0.642-0.721) | 0.682 (0.617-0.738) | 0.616 (0.565-0.691) | 0.674 (0.619-0.749) |
| | U-Sleep | **0.778** (0.723-0.825) | **0.848** (0.809-0.886) | 0.782 (0.742-0.819) | **0.874** (0.829-0.915) | **0.724** (0.674-0.776) | **0.718** (0.667-0.779) |
| | DANA | 0.738 (0.685-0.798) | 0.812 (0.766-0.855) | **0.815** (0.778-0.839) | 0.795 (0.731-0.859) | 0.653 (0.601-0.706) | 0.701 (0.613-0.766) |
| | SleepFM | 0.704 (0.654-0.749) | 0.787 (0.742-0.828) | 0.793 (0.758-0.822) | 0.804 (0.747-0.845) | 0.677 (0.620-0.732) | 0.692 (0.625-0.760) |
| ECG+EOG | Raw | 0.619 (0.563-0.691) | 0.654 (0.595-0.706) | 0.690 (0.654-0.732) | 0.654 (0.575-0.716) | 0.579 (0.514-0.644) | 0.607 (0.544-0.698) |
| | U-Sleep | **0.791** (0.739-0.839) | **0.814** (0.778-0.860) | 0.775 (0.728-0.804) | 0.779 (0.721-0.839) | **0.683** (0.635-0.735) | 0.672 (0.614-0.733) |
| | DANA | 0.750 (0.690-0.808) | 0.773 (0.720-0.811) | **0.777** (0.737-0.808) | 0.734 (0.665-0.803) | 0.642 (0.586-0.696) | **0.739** (0.666-0.802) |
| | SleepFM | 0.733 (0.682-0.772) | 0.800 (0.750-0.844) | 0.775 (0.745-0.807) | **0.811** (0.755-0.858) | 0.670 (0.613-0.728) | 0.688 (0.626-0.754) |
| EEG+EOG | Raw | 0.646 (0.585-0.708) | 0.626 (0.573-0.688) | 0.629 (0.589-0.670) | 0.589 (0.517-0.659) | 0.589 (0.532-0.653) | 0.628 (0.562-0.706) |
| | U-Sleep | **0.747** (0.688-0.799) | **0.812** (0.757-0.853) | 0.762 (0.723-0.791) | 0.815 (0.752-0.864) | 0.649 (0.581-0.698) | 0.689 (0.634-0.743) |
| | SleepFM | 0.733 (0.672-0.772) | 0.812 (0.764-0.850) | **0.777** (0.748-0.811) | **0.818** (0.765-0.866) | **0.686** (0.631-0.736) | **0.698** (0.639-0.761) |

**Table S14 – Continued from previous page**

| Configuration | Model | Angina | CVD Death | CHF | CHD Death | MI | Stroke |
|---|---|---|---|---|---|---|---|
| ECG | Raw | 0.471 (0.392-0.544) | 0.609 (0.543-0.672) | 0.632 (0.588-0.675) | 0.573 (0.493-0.664) | 0.588 (0.540-0.642) | 0.586 (0.524-0.661) |
| | U-Sleep | **0.783** (0.729-0.836) | **0.807** (0.754-0.862) | 0.756 (0.710-0.787) | **0.820** (0.774-0.873) | **0.681** (0.635-0.731) | 0.661 (0.600-0.731) |
| | DANA | 0.724 (0.669-0.784) | 0.789 (0.739-0.831) | **0.791** (0.757-0.825) | 0.765 (0.694-0.813) | 0.640 (0.584-0.687) | **0.716** (0.638-0.785) |
| | SleepFM | 0.634 (0.572-0.693) | 0.664 (0.616-0.726) | 0.690 (0.639-0.727) | 0.666 (0.596-0.731) | 0.635 (0.577-0.697) | 0.557 (0.483-0.629) |
| EEG | Raw | 0.704 (0.654-0.754) | 0.673 (0.620-0.715) | 0.652 (0.608-0.688) | 0.684 (0.609-0.736) | 0.612 (0.547-0.677) | 0.697 (0.639-0.756) |
| | U-Sleep | 0.719 (0.666-0.768) | **0.834** (0.783-0.875) | 0.755 (0.713-0.786) | 0.825 (0.773-0.865) | 0.582 (0.512-0.631) | 0.660 (0.607-0.715) |
| | SleepFM | **0.725** (0.655-0.778) | 0.815 (0.780-0.854) | **0.780** (0.752-0.811) | **0.827** (0.786-0.860) | **0.684** (0.627-0.735) | **0.705** (0.640-0.752) |
| EOG | Raw | 0.640 (0.587-0.705) | 0.625 (0.556-0.686) | 0.620 (0.584-0.655) | 0.615 (0.548-0.691) | 0.577 (0.515-0.640) | 0.590 (0.506-0.659) |
| | U-Sleep | 0.735 (0.674-0.785) | **0.776** (0.714-0.820) | 0.707 (0.670-0.737) | 0.742 (0.688-0.792) | 0.622 (0.564-0.698) | 0.589 (0.506-0.673) |
| | SleepFM | **0.771** (0.713-0.811) | 0.756 (0.708-0.807) | **0.733** (0.696-0.768) | **0.765** (0.706-0.827) | **0.677** (0.625-0.727) | **0.678** (0.612-0.737) |

**Table S15**: Calibration and overall predicted accuracy at a 5-year horizon, on the SHHS1 test cohort and the SHHS2 follow-up cohort. Values are macro-averages across the six cardiovascular outcomes.

| | | **SHHS1 test** | | | **SHHS2** | | |
|---|---|---|---|---|---|---|---|
| **Configuration** | **Model** | **Slope** | **CITL** | **IPA** | **Slope** | **CITL** | **IPA** |
| | | **With demographics** | | | | | |
| All Demo | Raw | 1.18 | **0.01** | 0.04 | **1.01** | **0.05** | +0.01 |
| | U-Sleep | 1.08 | 0.04 | **+0.05** | 1.02 | -0.15 | +0.04 |
| | DANA | **0.98** | 0.04 | **+0.05** | 0.99 | -0.15 | **+0.05** |
| Without Age | Raw | **0.90** | 0.10 | **+0.03** | 0.60 | 0.54 | +0.00 |
| | U-Sleep | 1.12 | **0.04** | +0.02 | **1.34** | 0.37 | **+0.04** |
| | DANA | 0.67 | 0.16 | +0.01 | 0.54 | **0.29** | +0.01 |
| Only Age and Gender | Raw | **0.92** | 0.21 | +0.04 | 0.90 | **0.05** | +0.04 |
| | U-Sleep | **0.92** | **0.03** | +0.04 | **1.00** | -0.14 | +0.05 |
| | DANA | 0.81 | 0.10 | +0.02 | 0.75 | -0.20 | +0.00 |
| | SleepFM | 1.29 | **0.03** | **+0.05** | 1.48 | -0.12 | **+0.06** |
| Only Gender | Raw | 0.68 | 0.14 | +0.02 | 0.51 | 0.47 | +0.01 |
| | U-Sleep | **1.10** | **0.04** | +0.02 | 1.33 | 0.39 | **+0.04** |
| | DANA | 0.75 | 0.15 | +0.02 | 0.65 | **0.22** | +0.02 |
| | SleepFM | 1.11 | **0.04** | **+0.05** | **1.01** | 0.49 | **+0.04** |
| | | **Without demographics** | | | | | |
| Only Signals | Raw | 1.93 | -0.20 | -0.00 | 2.08 | -0.21 | -0.00 |
| | U-Sleep | 1.23 | 1.41 | +0.00 | 1.22 | 1.74 | -0.00 |
| | DANA | **0.97** | 3.05 | -0.03 | **0.91** | 3.23 | -0.03 |
| | SleepFM | 1.56 | **0.03** | **+0.02** | 1.57 | **0.4** | **+0.02** |
| ECG+EEG | Raw | 0.81 | 0.13 | **+0.03** | 0.54 | 0.81 | +0.00 |
| | U-Sleep | **0.99** | -0.87 | -0.21 | **1.07** | -0.51 | -0.09 |
| | DANA | 0.59 | 0.28 | -0.03 | 0.56 | **0.33** | -0.01 |
| | SleepFM | 2.25 | **0.03** | **+0.03** | 1.84 | 0.44 | **+0.02** |
| ECG+EOG | Raw | 0.51 | 0.26 | -0.01 | 0.38 | 0.56 | -0.03 |
| | U-Sleep | **1.02** | 1.53 | -0.00 | **1.09** | 1.90 | -0.01 |
| | DANA | **1.02** | 0.06 | **+0.03** | 0.89 | **0.27** | **+0.03** |
| | SleepFM | 1.18 | **0.03** | +0.02 | 1.23 | 0.46 | +0.02 |
| EEG+EOG | Raw | 0.65 | 0.08 | -0.01 | 0.46 | 0.54 | -0.02 |
| | U-Sleep | **0.98** | 2.82 | -0.02 | **1.10** | 2.97 | -0.02 |
| | SleepFM | 1.18 | **0.03** | **+0.02** | 1.23 | **0.46** | **+0.02** |
| ECG | Raw | 0.63 | **0.04** | +0.01 | 0.49 | **0.30** | -0.00 |
| | U-Sleep | **1.24** | 0.48 | **+0.02** | 1.25 | 0.77 | **+0.02** |
| | DANA | 0.62 | 0.17 | -0.01 | 0.57 | 0.34 | -0.01 |
| | SleepFM | 1.37 | **0.04** | **+0.02** | **1.07** | 0.48 | +0.01 |
| EEG | Raw | 0.79 | 0.17 | -0.02 | 0.76 | 0.42 | -0.02 |
| | U-Sleep | 1.22 | 2.06 | -0.01 | 1.31 | 2.14 | -0.02 |
| | SleepFM | **1.06** | **0.04** | **+0.02** | **1.14** | **0.29** | **+0.03** |
| EOG | Raw | 0.85 | 0.04 | -0.01 | 0.69 | 0.46 | -0.02 |
| | U-Sleep | **1.01** | 2.5 | -0.02 | **0.95** | 2.74 | -0.02 |
| | SleepFM | 2.17 | **0.02** | **+0.01** | 2.63 | **0.3** | **+0.01** |

**CITL**: Calibration-In-The-Large; **IPA**: Index of Prediction Accuracy.

**Table S16**: Pairwise C-index differences: U-Sleep ECG+EEG vs ECG-only models on SHHS1 test and SHHS2. Cells show ΔC-index. Statistical significance: * $p \leq 0.1$.

| | **SHHS1 Test** | | **SHHS2 All** | |
|---|---|---|---|---|
| **Outcome** | U-Sleep ECG+EEG *vs* U-Sleep ECG | U-Sleep ECG+EEG *vs* Self-DANA ECG | U-Sleep ECG+EEG *vs* U-Sleep ECG | U-Sleep ECG+EEG *vs* Self-DANA ECG |
| Angina | −0.006 | −0.006 | −0.008 | +0.067* |
| CVD Death | −0.016 | −0.016 | +0.030 | +0.016 |
| CHF | +0.025 | −0.025 | +0.023 | +0.014 |
| CHD Death | −0.002 | −0.024 | +0.047 | +0.060* |
| MI | −0.016 | −0.009 | +0.027 | +0.078* |
| Stroke | +0.032 | +0.053* | +0.042 | +0.001 |
| Macro Average | +0.003 | −0.005 | +0.027* | +0.039* |

**Table S17**: Pairwise C-index differences: U-Sleep ECG+EEG vs ECG-only on SHHS2 patients that were present in the SHHS1 train cohort and patients completely out from the training set. Cells show ΔC-index. Statistical significance: * $p \leq 0.1$

| | **SHHS2 training-overlap** | | **SHHS2 training-disjoint** | |
|---|---|---|---|---|
| **Outcome** | U-Sleep ECG+EEG *vs* U-Sleep ECG | U-Sleep ECG+EEG *vs* Self-DANA ECG | U-Sleep ECG+EEG *vs* U-Sleep ECG | U-Sleep ECG+EEG *vs* Self-DANA ECG |
| Angina | −0.019 | +0.038 | +0.007 | +0.052 |
| CVD Death | +0.029 | +0.018 | +0.012 | +0.025 |
| CHF | +0.013 | +0.002 | +0.037 | +0.006 |
| CHD Death | +0.034 | +0.095* | +0.043 | +0.037 |
| MI | +0.006 | +0.039 | +0.037 | +0.096 |
| Stroke | +0.093 | −0.009 | +0.019 | +0.033 |
| Macro Avg | +0.026 | +0.031 | +0.026 | +0.042 |

**Table S18**: Pairwise C-index differences (Δ = Coupled Mamba − LSTM) across SHHS1 evaluation subsets. Cells show ΔC-index: positive values indicate superior performance of the Coupled Mamba fusion method with respect to LSTM with the raw signals as input. Statistical significance: * $p \leq 0.1$.

| **Outcome** | **All test** | **Primary Prev** | **Secondary Prev** |
|---|---|---|---|
| Angina | +0.008 | +0.015 | -0.064 |
| CVD Death | +0.116* | +0.122* | +0.095* |
| CHF | +0.054* | +0.060* | +0.038 |
| CHD Death | +0.152* | +0.130* | +0.162* |
| MI | +0.026 | +0.003 | +0.035 |
| Stroke | +0.113* | +0.145* | +0.075 |
| Macro Avg | +0.078* | +0.079* | +0.057* |

**Table S19**: Pairwise C-index differences ($\Delta$ = Coupled Mamba – LSTM) across SHHS2 evaluation subsets. Cells show $\Delta$C-index: positive values indicate superior performance of the Coupled Mamba fusion method with respect to LSTM with the raw signals as input. Statistical significance: * $p \leq 0.1$.

| **Outcome** | **All** | **Primary Prev** | **Secondary Prev** | **Train** | **No train** |
|---|---|---|---|---|---|
| Angina | +0.124* | +0.211* | +0.041 | +0.088 | +0.168* |
| CVD Death | +0.162* | +0.045 | +0.149* | +0.188* | +0.140* |
| CHF | +0.171* | +0.160* | +0.108* | +0.245* | +0.107* |
| CHD Death | +0.161* | +0.175* | +0.104* | +0.187* | +0.148* |
| MI | +0.077* | +0.033 | +0.126* | +0.170* | -0.022 |
| Stroke | +0.162* | +0.058 | +0.085 | +0.196* | +0.111* |
| Macro Avg | +0.143* | +0.114* | +0.102* | +0.179* | +0.109* |

**Table S20**: For each endpoint, we report the number of subjects (N) and the corresponding event/non-event counts (Ev/No). Results are shown for the SHHS1 test set and for SHHS2, each stratified into primary and secondary prevention. SHHS2 is additionally stratified by overlap with the SHHS1 training cohort (overlapping vs training-disjoint). Subjects categorized as "uncertain" were handled according to our masking strategy.

| | **SHHS1 Test Set** | | | | **SHHS2** | | | | | | | |
|---|---|---|---|---|---|---|---|---|---|---|---|---|
| | Primary | | Secondary | | Primary | | Secondary | | Training-overlap | | Training-disjoint | |
| **Outcome** | **N** | **Ev/No** | **N** | **Ev/No** | **N** | **Ev/No** | **N** | **Ev/No** | **N** | **Ev/No** | **N** | **Ev/No** |
| **Total Population** | **1744** | | **247** | | **2010** | | **608** | | **1353** | | **1265** | |
| Angina | | 80/1113 | | 42/149 | | 40/1145 | | 59/392 | | 58/785 | | 41/752 |
| CVD Death | | 70/1674 | | 56/190 | | 34/1976 | | 78/530 | | 61/1292 | | 51/1214 |
| CHF | | 131/1613 | | 86/161 | | 65/1945 | | 148/460 | | 105/1248 | | 108/1157 |
| CHD Death | | 43/1701 | | 36/210 | | 23/1987 | | 53/555 | | 39/1314 | | 37/1228 |
| MI | | 88/1656 | | 37/210 | | 65/1945 | | 56/552 | | 64/1289 | | 57/1208 |
| Stroke | | 73/1671 | | 45/202 | | 18/1992 | | 55/553 | | 34/1319 | | 39/1226 |

**Table S21**: PSG preprocessing, tokenization, and representation-extraction settings for U-Sleep, Self-DANA, SleepFM, and raw end-to-end encoders.

| **Setting** | **U-Sleep** | **Self-DANA** | **SleepFM** | **Raw CNN** |
|---|---|---|---|---|
| Resampling | 128 Hz (polyphase) | 500 Hz (cubic-spline) | 128 Hz (linear) | 128 Hz (linear) |
| Normalization | Median 0, IQR 1 (per channel/subject) | Mean removal + MA(5) + BP 0.5–40 Hz | Z-score | Z-score |
| Frozen encoder | True | True | True | False |
| Token resolution | 1 s | 5 s | 5 s | 5 s |
| Token dimension | 6 | 768 | 128 | $d_{\text{model}}$ |
| Trainable pre-fusion layers | Linear projection to $d_{\text{model}}$ | Linear projection to $d_{\text{model}}$ | Linear projection to $d_{\text{model}}$ | – |

**IQR**: interquartile range; **MA(5)**: 5th-order moving-average filter; **BP**: band pass filter

**Table S22**: Hyperparameter search space and optimal configuration. Details of the explored ranges and selected values for the Coupled Mamba framework architecture, pooling strategy, and optimization parameters.

| **Category** | **Hyperparameter** | **Search Space** | **Selected Value** |
|---|---|---|---|
| **Architecture** | Model Dimension ($d_{model}$) | 32, 64, 128 | **64** |
| | Number of Layers ($N$) | 2, 4, 6 | **2** |
| | State Dimension ($d_{state}$) | 16, 32, 64 | **16** |
| **Pooling** | Global Pooling Strategy | Average, Last Token | **Average** |
| **Optimization** | Learning Rate | [1e-4 - 1e-1][a] | **0.002** |
| | Weight Decay | [1e-4 - 1e-1][b] | **0.1** |
| | Dropout Rate | 0.1, 0.2, 0.3 | **0.2** |
| | Batch Size | 16, 32, 64 | **32** |

a: Log-uniform distribution; b: Continuous uniform distribution
